# Taxation of foods high in fat, sugar, and sodium in India: A modelling study of health and economic impacts

**DOI:** 10.1101/2025.03.06.25323478

**Authors:** Maxime Roche, Jingmin Zhu, Jack Olney, Daniel J Laydon, William Joe, Manika Sharma, Lindsay Steele, Franco Sassi

## Abstract

**Background:** Consumption of foods high in fat, sugar, and sodium (HFSS) and obesity are rapidly increasing in India. Taxing HFSS foods has been proposed as one of the policy interventions to promote healthier diets globally. This study estimates the effect of this approach on nutrient intake, diet-related disease, and associated health and economic burdens in India.

**Methods and findings:** We use nationally representative household expenditure survey data, dietary requirements, and food composition tables to model individual nutrient intake. Consumer responsiveness to food price changes for three income terciles, captured in price elasticities, is estimated using an Almost Ideal Demand System model. Longer-term policy impacts are estimated through a novel dynamic microsimulation model, Health-GPS. Modelled policy outcomes include changes in risk exposures, disease incidence and burden, and total health expenditure. On average, 9.9% of total energy intake comes from HFSS items, based on the definition by the Food Safety and Standards Authority of India’s Labelling & Display Amendment Draft Regulations 2022. Applying the highest Goods and Services Tax (GST) rate of 40% on HFSS items is associated with a persistent average per capita decrease of 0.17kg/m^2^ (95% CI: −0.17, −0.17) in body mass index and 45.8mg (95% CI: −45.9, −45.7) in daily sodium intake. Over 30 years, this could reduce annual disease incidence by up to 1.72% (95% CI: −1.78%, −1.66%) on average and prevent 0.63 million (95% CI: −0.71, −0.55) disability-adjusted life years per year from ischemic heart disease, chronic kidney disease, stroke, diabetes, and asthma, reducing total health expenditure by US$601 million (95% CI: −624, −578) per year. Further work is needed to incorporate potential benefits or harms associated with changes in other foods and nutrients that are not currently modelled, such as fruits, whole grains, fibre, and red and processed meats. Larger absolute health gains accrue to higher-income individuals, reflecting higher baseline HFSS food intake. Given substitution patterns and a price-inelastic demand, the tax change is expected to generate a 92.0% (95% CI: 88.2%, 95.7%) increase in tax revenue from foods and beverages with only a minor effect on household spending (+1.0%, 95% CI: +0.0%, +1.9%).

**Conclusions:** Higher taxation of HFSS foods could help mitigate rising incidence of diet-related diseases and morbidity in India, reduce healthcare costs, and serve as an additional source of revenue for the government.

## Introduction

Unhealthy diets and obesity are major risk factors for non-communicable diseases, such as cardiovascular disease, cancer, and diabetes. These diseases account for substantial healthcare costs, as well as wider economic costs for governments and individuals [1]. In India, adult overweight and obesity levels have increased fourfold since 1975 [2]. Of particular concern is the increasing consumption of highly industrially processed foods rich in energy, saturated fat, sugar, and sodium [3].

Governments can promote healthy diets through fiscal policies, including the taxation of foods high in fat, sugar, and sodium (HFSS), as well as subsidies on fruits and vegetables, alongside food marketing regulation and front-of-pack nutrition labelling. There are strong health and economic rationales for using fiscal policies to address the external costs of unhealthy diets and obesity (i.e. ‘externalities’, such as collectively borne health care costs linked with obesity) while “encouraging people to avoid acting against their own self-interest” (i.e., ‘internalities’) [4,5].

India applies a Goods and Services Tax (GST), introduced in July 2017, at five different rates: 0%, 5%, 12%, 18%, and 28%. Among food and beverage items, the 28% rate only applies to caffeinated and sweetened aerated beverages, which include sugar-sweetened beverages (SSBs), along with an additional 12% compensatory Cess. This cess was part of a nationwide mechanism intended to compensate states for any shortfall in their tax revenue for five years following the introduction of the GST system, relative to a baseline assuming 14% annual growth in their 2015–16 tax revenue from taxes subsumed under GST. Adjusting existing GST rates to align with nutrition and health objectives may improve diets by shifting food preferences.

Only a few countries have implemented taxes on HFSS foods, with limited scope, targeting either specific food categories (e.g., confectionery and ice cream in Finland or French Polynesia), non-essential energy-dense foods (e.g., in Mexico), or selected nutrients (e.g., sugar and salt in Hungary) [6]. Identifying HFSS foods for taxation based on nutrient content, for example, using a nutrient profile model (NPM), may capture unhealthy foods more broadly, and is less likely to apply high rates to healthier foods or incentivise negative substitutions from ‘healthy’ to ‘less healthy’ foods [7]. NPMs are increasingly used in nutrition policies, such as front-of-pack nutrition labelling and marketing restrictions. In 2023, Colombia became the first country to introduce a broad excise tax on HFSS foods based on an NPM. The government subsequently increased the excise tax rate in 2024 and 2025, offering a promising example for other countries [8].

This study estimates the potential impacts on nutrient intakes and diet-related diseases of aligning India’s GST rate differentiation on foods and beverages to nutritional quality, placing a higher tax burden on HFSS foods, in line with suggested policy intervention globally [6]. We first estimate baseline energy and nutrient intakes, as well as consumers’ responsiveness to food price changes. We then calibrate a microsimulation model to measure the effects of alternative taxation scenarios on various health and economic outcomes, as well as their distribution across income groups, over the next 30 years.

Simulation analyses in Costa Rica, the Philippines, and the UK have suggested that NPM-based taxes can significantly improve diet [9–11]. The long-term population health and economic impacts of such a comprehensive nutrient-based approach remain understudied, especially in low- and middle-income countries. A recent study investigated the potential impact of an excise tax on HFSS foods in India on consumption and tax revenue, without considering nutrition or health outcomes [12]. While others have simulated SSB taxes and their impact on non-communicable diseases in middle-income countries [13], including in India [14], our study fills this gap for broader HFSS food taxation.

We contribute to the literature by applying a novel microsimulation model, Health-GPS, to evaluate the impacts of HFSS food taxation in India. The model’s architecture involves a hierarchy of demographic and socio-economic characteristics, risk factors, and diseases [15]. We simulate 0.5% of the Indian population to estimate policy impacts on risk factors, disease incidence, and disability-adjusted life years (DALYs) as well as associated health expenditure. We provide the most recent estimates of macronutrient intakes in the Indian population using the latest National Sample Survey Office (NSSO) Household Consumption Expenditure survey 2022-23. Lastly, we present updated estimates of the price and income elasticity of demand for foods and beverages by income groups. These represent essential metrics to estimate the potential impact of alternative fiscal reforms.

## Methods

### Estimating energy and nutrient intake

Our primary data source is the 2022-23 Household Consumption Expenditure survey from the NSSO. This survey collected food and beverage consumption, and expenditure from a nationally representative sample of 261,746 households between August 2022 and July 2023 (**Table A1** in **S1 Supporting Information** provides household characteristics) [16]. Food consumption is recorded over two recall periods: 30 days for cereals, pulses, sugar, and salt, and 7 days for all other items. We harmonised these data to obtain daily household consumption for each item and matched them to their respective nutrient content. We extract nutrient information (energy, sugar, sodium, saturated fat, total fat, carbohydrates, and protein) from the National Institute of Nutrition (NIN) Indian Food Composition Tables 2017 [17], complemented by the United States Department of Agriculture (USDA) Food and Nutrient Database for Dietary Studies 2017-2018 [18]. As NSSO items do not differentiate between processing levels for meat and fish, we assume that 20% of purchased quantities are processed for these groups, based on expert opinion. We individualise household-level intake based on household members’ age and sex using the daily NIN Dietary Guidelines for Indians, 2024 [19]. Lastly, we assume that 55kg of food is wasted yearly by the average Indian to better proxy dietary intake based on data from the UN Environmental Programme [20]. The final sample size is 256,464 households after dropping households with missing food consumption information and 1% lowest outliers (i.e., daily per capita energy intake lower than 933 kcal) and 1% highest outliers (i.e., daily per capita energy intake higher than 4,673 kcal).

### Estimating response to food price changes

We estimate consumer responses to food price changes, accounting for food substitutions (own- and cross-price elasticities of demand) using the 2022-23 Household Consumption Expenditure survey data and an Almost Ideal Demand System (AIDS) model. In this utility-based structural model, goods are heterogeneous, and households choose the quantity and quality of a given good as a function of its price, the price of other goods, household income, and household sociodemographic characteristics. Following Deaton (1988), we adjust for quality shading and measurement error stemming from the use of unit values as a proxy for unobservable prices (i.e., expenditure divided by quantity) [21]. This model is extensively used to estimate the demand for food in low- and middle-income countries [22]. It assumes spatially varying prices, where all households within a near geographical area, here a cluster, defined by NSSO as a village or urban block, face the same price, regardless of their income group. Any within-cluster variation in unit values is due to differences in the quality of the purchased items. Spatial variations in unit values between clusters provide an identification strategy to avoid the endogeneity of prices and address quality shading.

This paper uses uncompensated price elasticities, which capture the effect of a change in the price of a good on demand, holding income and other prices constant. These elasticities reflect both the substitution effect and the income effect of the price change. Details of the microeconomic foundations of Deaton’s (1988) AIDS model, as well as its derivation and the steps involved in its estimation, have been described elsewhere [23]. More information is provided in **Appendix C** in **S3 Supporting Information**.

We estimate the model parameters for eleven groups: cereals, dairy, pulses, edible oils and spices (including salt and raw sugar), fruits and vegetables (including nuts, hereafter referred to as F&V), animal meats (fresh), packaged processed foods, sweets, SSBs, non-SSBs, and a *numeraire* group containing food-away-from-home and non-food expenditure. Adding the latter allows reallocation between food and non-food when prices or real expenditure change. We normalise the *numeraire* group’s unit value to one. After dropping clusters that do not have at least two households consuming at least one item for each group [24], we are left with a total of 5,432 clusters and 212,797 households. We fit this model for three income groups, defined by terciles of monthly per capita consumption expenditure.

### Fiscal policy scenarios

The estimated income-group-specific own- and cross-price elasticities are used to simulate the impacts of two main scenarios consisting of increasing GST to the highest rate of 28% on foods and beverages classified as HFSS based on their nutrient composition, namely the proposed HFSS food definition from the Food Safety and Standards Authority of India (FSSAI) Labelling & Display Amendment Draft Regulations 2022 [25] (scenario 1) as well as an additional 12% (equivalent in effect to a 40% tax rate) (scenario 2). This latter scenario aligns with the “sin” goods GST rate (40%), which is applied to caffeinated and sweetened aerated beverages (including SSBs) and some tobacco products.

The proposed FSSAI definition applies uniform sodium, sugar, and saturated fat thresholds across all processed food and beverage products. We assume processed products to be items not included in Schedule IV, Category III Solid Foods/Liquid Foods, which are exempt from front-of-pack nutrition labelling under the Indian Nutrition Rating, as per the FSSAI draft regulation [25]. The impact of using an alternative NPM to define HFSS foods, namely the one developed for the South East Asian Region by the World Health Organisation (WHO SEARO) [26], based on category-specific nutrient thresholds for energy, sodium, sugar, saturated fat, and total fat content (**Table A2** in **S1 Supporting Information**), is tested in further simulations using the same tax rates as in scenarios 1 and 2 (scenarios 3 and 4, respectively). **Figure A1** and **Figure A2** in **S1 Supporting Information** present the baseline distribution of total energy intake by GST rate and food group and by GST rate and HFSS status (based on the FSSAI definition), respectively. They show that most HFSS items are currently taxed at 12% or 18%.

The immediate impact of each policy scenario on individual energy and nutrient intake, total household expenditure on foods and beverages, and government tax revenue from foods and beverages is calculated based on the estimated price elasticities. Key assumptions, which are subsequently relaxed in sensitivity analyses (**Table B1** in **S2 Supporting Information**), include a 100% tax passthrough to retail prices and no supply-side tax-induced product reformulation. Finally, we also test the impact of a 0% GST on F&V and pulses (equivalent to a subsidy). Results sensitivity analyses are presented in **Appendix B** in **S2 Supporting Information**.

### Health-GPS microsimulation model

Long-term nutrient intake projections and policy impacts are estimated using Health-GPS, a dynamic policy microsimulation model developed by the Centre for Health Economics & Policy Innovation [27]. Health-GPS creates a synthetic population of individuals that broadly reproduces demographic and socioeconomic characteristics of the population of a given country, or sub-national jurisdiction. It simulates individual life histories from birth to death, including age, gender, socioeconomic characteristics, risk factors, and disease profiles. Health-GPS updates annually with statistical and probabilistic models, which are calibrated with real-world data to capture relationships between variables. Changes in disease risks are assumed to result solely from changes in demographic composition and risk factor distributions, with no underlying time trend in diseases.

A baseline scenario is simulated reflecting expected demographic and epidemiological changes over a time horizon in a given population. Intervention scenarios are then simulated, reflecting the introduction of policy interventions applied to targeted risk factors. Changes in risk factor exposures will, in turn, impact other risk factors, risk of diseases, and disease burden. Disease outcomes typically include incidence, prevalence, and mortality of each disease in the population. Disease burden is measured by disability-adjusted life years (DALYs), a sum of years of life lost (YLLs) and years lived with disability (YLDs). To calculate YLLs and YLDs, the model iterates over each individual in the simulated population, summing the YLLs due to premature mortality and the YLDs, weighted by the corresponding disability weights. Policy impacts are estimated as the difference in these outcomes between the baseline and intervention scenarios.

In this study, 0.5% of the Indian population is simulated at the individual level, repeated 20 times for each scenario, between 2022 and 2053, with policy introduced in 2024. The synthetic population is modelled to reflect demographic estimates and projections from the United Nations World Population Prospects [28], estimated distribution of individual energy and nutrient intakes, weight and height distributions by age and gender from NCD-RisC [29], as well as prevalence, incidence and excess mortality of diseases from the Institute for Health Metrics and Evaluation Global Burden of Disease [30]. Specifically, for each individual, we simulate two demographic characteristics: age, gender; two socio-economic characteristics: income tertile, sector (urban/rural); six risk factors including four nutrients (carbohydrate, protein, fat, and sodium), physical activity, and body mass index (BMI); disease status of five key diseases: ischemic heart disease (IHD), chronic kidney disease (CKD), stroke (including ischemic stroke, intracerebral haemorrhage and subarachnoid haemorrhage), diabetes, and asthma. A complete list of model parameters and variables definition and sources is presented in **Table D1** in **S4 Supporting Information**.

We assume two main diet-related risk-factor-to-disease pathways in this study: the first reflects the impact of diet (and physical activity) on energy balance and thereafter BMI; the second accounts for the impact of dietary sodium. Diet composition-related benefits (e.g., increased consumption of whole grains, F&V, pulses) are not modelled. With data on two risk factors and relative risk ratios linking risk factors to diseases from the literature, changes in incidence and disease burden of five key diseases are estimated. In addition, changes in hypertension prevalence are estimated independently with projected risk factor data from Health-GPS.

We apply two alternative assumptions on the time trend of nutrient intake. First, we assume no time trends in age-, sex- and income-group-specific nutrient intakes, which means that socio-demographic shifts solely drive population-wide changes. Microsimulation outcomes under this assumption thus represent a lower bound of estimates and policy impacts (hereafter referred to as the ‘lower bound’). Our alternative assumption is an income trend which accounts for a constant 6% income growth based on IMF projections for real GDP growth [31]. It is scaled by the estimated income group- and food group-specific income elasticities (**Table C1** and **Figure C1** in **S3 Supporting Information**) and an exponential decay over time, based on Engel’s Law, which posits that, as household income increases, the proportion spent on food decreases [32]. The decay rate is nutrient-specific and estimated by regressing the logarithm of the predicted growth rate in individual nutrient intake on a time trend. The former is estimated based on a regression of individual nutrient intake on monthly per capita total expenditure and a set of household- and individual-level controls among the NSS Household Consumption Expenditure 2022-23 sample. **Figure E1** in **S5 Supporting Information** cross-validates this approach by comparing the estimated yearly growth rate in the consumption of packaged processed foods and sweets - two food groups with a high proportion of HFSS foods (**Table A6** in **S1 Supporting Information**) - with historical trends based on Euromonitor International Passport data and Tak et al (2022) for selected highly processed food items [3,33]. This alternative assumption gives an upper-bound estimate of policy impacts (the ‘upper bound’). More details on how we derive the parameters for the income trend assumption can be found in **Appendix E** in **S5 Supporting Information**.

Policy scenarios are applied at the beginning of 2024 (two years after the initial year) and their impact on health is estimated over 30 years until 2053. We report microsimulations outcomes including the reduction in major risk factors: sodium intake and BMI, cumulative reduction in disease incidence and DALYs, in the general population and by income group. Economic benefits are calculated as averted total health expenditures in US dollar 2024, including both government and household expenditure. Household health expenditures per disease per year are derived from the literature and converted to total health expenditure based on data from the literature and National Health Accounts Estimates for India 2021-22 [34]. Confidence intervals are constructed with results from 20 repeated simulation runs to capture stochastic variation.

A simplified model diagram describing the Health-GPS model can also be found in **Figure D1** in **S4 Supporting Information**. More details on the methods related to its use are listed in **Appendix D** in **S4 Supporting Information**. Lastly, **Figure 1** offers a simplified flow chart of the data sources and methodological steps taken in this analysis.

**Figure 1.**
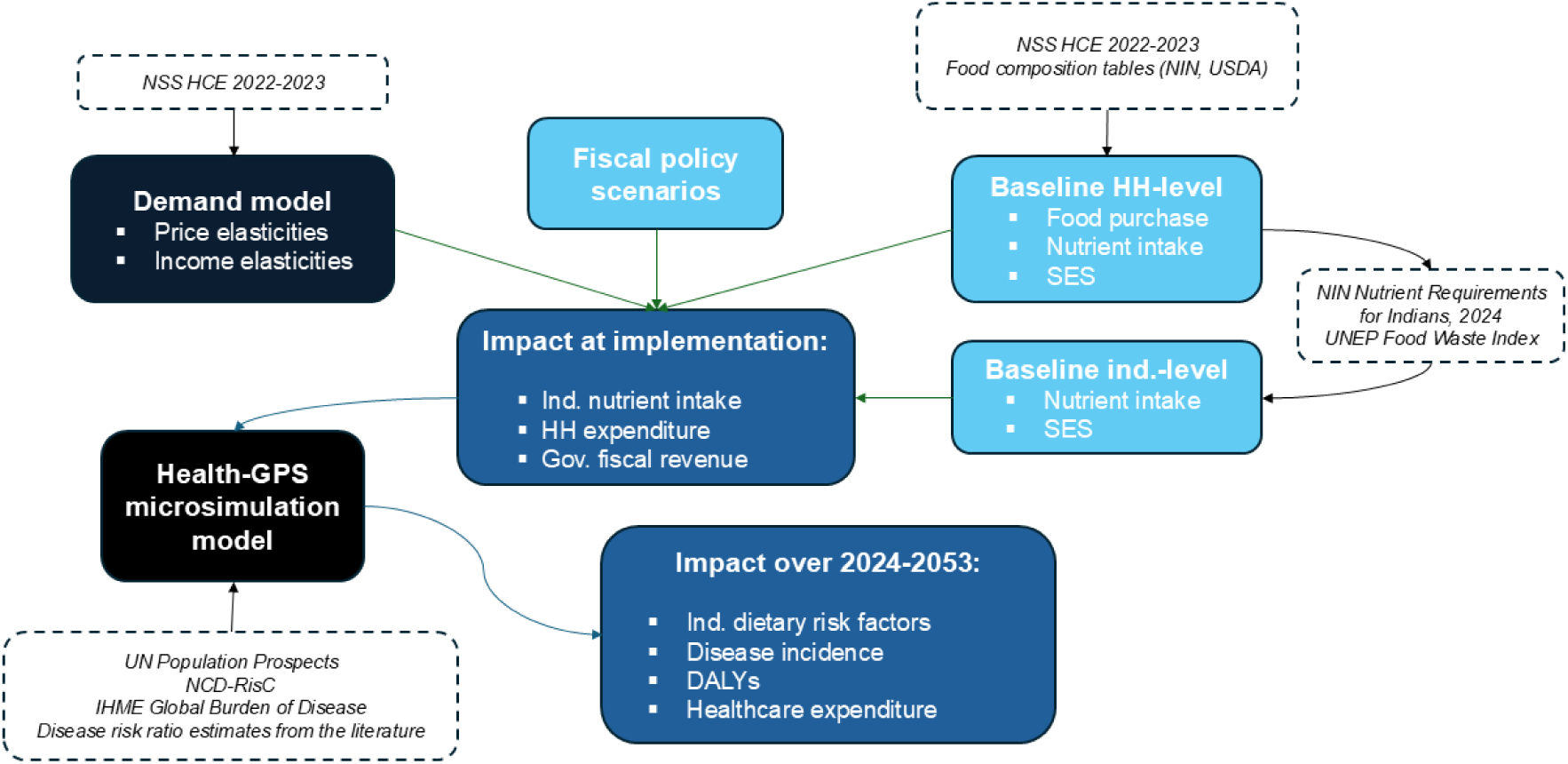
Analysis steps flow chart. Notes: DALYs: Disability-adjusted life years; Gov: Government; HH: Household; IHME: Institute for Health Metrics and Evaluation; Ind: Individual; NIN: National Institute of Nutrition; NSS HCE: National Sample Survey Household Consumption Expenditure; SES: Socio-economics characteristics; UN: United Nations; UNEP: United Nations Environmental Programme; USDA: United States Department of Agriculture.

## Results

### Energy and nutrient intakes

**Table 1** presents the estimated baseline average daily nutrient intake per capita. **Tables A3-A5** in **S1 Supporting Information** show the results for low-, middle-, and high-income groups, respectively. The average daily energy intake is lower than previous NSS-based estimates, which did not account for waste [35]. Cereals represent approximately half of the total calorie intake. Total daily energy intake increases with income. While our estimated average daily sodium intake is slightly more conservative than subnational dietary recall estimates [36], we find that 80.7% of adult males, 73.0% of adult females, and 80.8% of adolescents have daily sodium intakes above the WHO-recommended level of 2,000mg/day. Three in ten adults also consume more saturated fat daily than the WHO-recommended levels (no more than 10% of total daily energy intake). For total sugars, 10.0% of adult males and 6.5% of adult females have a daily total sugar intake above the National Health Service of the United Kingdom (NHS) recommended levels (no more than 90g per day) (**Figure A3** in **S1 Supporting Information**).

**Table 1.** Estimated average daily per capita energy and nutrient intake, full sample.

|  | Energy (kcal) | Sugar (g) | Sat. fat (g) | Sodium (mg) | Carbs (g) | Total fat (g) | Protein (g) |
| --- | --- | --- | --- | --- | --- | --- | --- |
| Served proc. food | 103.6 | 0.7 | 1.0 | 134.8 | 14.1 | 4.1 | 2.7 |
| Cereals | 900.2 | 3.1 | 0.5 | 6.6 | 194.1 | 2.9 | 24.6 |
| Dairy | 161.8 | 8.5 | 6.9 | 58.5 | 10.9 | 10.5 | 5.9 |
| Pulses | 71.2 | 0.3 | 0.1 | 4.5 | 11.7 | 0.4 | 5.2 |
| Oils & spices | 361.9 | 20.4 | 3.8 | 2,650.9 | 23.3 | 29.3 | 1.3 |
| F&V | 193.9 | 9.9 | 5.2 | 20.9 | 22.2 | 9.5 | 4.8 |
| Animal meats, fresh | 27.2 | 0.0 | 0.4 | 17.4 | 0.1 | 1.5 | 3.3 |
| Packaged proc. foods | 89.1 | 0.6 | 1.4 | 185.4 | 9.8 | 4.2 | 3.0 |
| Sweets | 91.6 | 8.1 | 1.7 | 40.3 | 14.3 | 3.3 | 1.2 |
| SSBs | 12.0 | 2.2 | 0.0 | 2.7 | 2.7 | 0.1 | 0.2 |
| Non-SSBs | 2.6 | 0.0 | 0.0 | 6.5 | 0.6 | 0.0 | 0.0 |
| **Total** | 2,015.2 | 53.7 | 21.0 | 3,128.6 | 303.7 | 65.7 | 52.2 |

Notes: Survey weighted. Sat.: saturated; carbs: carbohydrates; kcal: kilo calorie; g: gram; mg: milligram. proc.: processed; F&V: fruits & vegetables; SSBs: sugar-sweetened beverages.

### Price elasticities of demand

**Table 2** shows the estimated average own- and cross-price elasticities of demand. Demand for most foods is inelastic (proportionate change in purchases smaller than proportionate change in price), except for dairy, SSBs, and non-SSBs. The demand for oils & spices is the least price-sensitive, with consumption decreasing by 3.7% for each 10% rise in oils & spices price (95% CI: 3.5%, 3.9% decrease). The demand for packaged processed foods, which mainly contain HFSS items (**Table A6** in **S1 Supporting Information**), is close to unit-elastic (−0.92, 95% CI: −0.95, −0.89). Results indicate significant substitution and complementarity patterns between groups. F&V and fresh animal meats are complements (negative cross-price elasticities, p<0.01). Packaged processed foods and fresh animal meats are substitutes, as well as SSBs and non-SSBs. **Figure 2** provides the estimated average own-price elasticity results by income group. **Tables A7-A9** in **S1 Supporting Information** additionally display the estimated cross-price elasticities. In line with the international literature and previous studies in India [22,37], the demand from low-income households is more price-sensitive, except for dairy.

**Figure 2.**
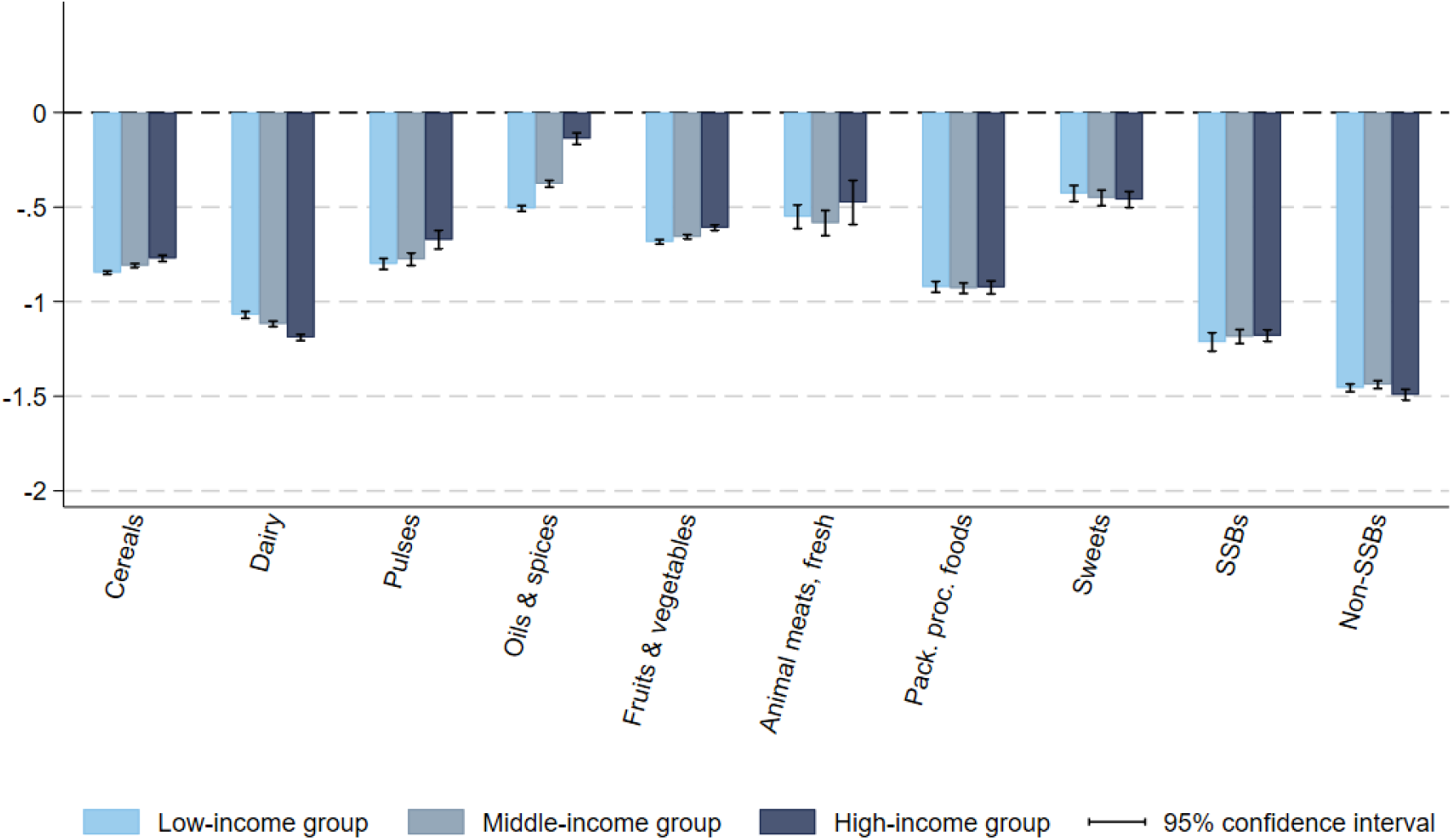
Estimated average own-price elasticity estimates, by income group. Notes: Estimated based on NSS Household Consumption Expenditure survey 2022-23 [16] and using Deaton (1988)’s quality-adjusted Almost Ideal Demand System model [21]. Bootstrapped standard errors after 300 replications. Vertical segments represent the 95% confidence intervals. Pack.: Packaged; proc.: processed; SSBs: sugar-sweetened beverages.

**Table 2.** Estimated average price elasticity estimates, full sample.

|  | Cereals | Dairy | Pulses | Oils & spices | F&V | Animal meats, fresh | Packaged proc. foods | Sweets | SSBs | Non-SSBs |
| --- | --- | --- | --- | --- | --- | --- | --- | --- | --- | --- |
| Cereals | **-0.794\*\*\*** | -0.089*** | 0.009*** | -0.023*** | 0.051*** | 0.026*** | -0.025*** | 0.025*** | 0.013** | 0.004 |
|  | **(0.004)** | (0.007) | (0.003) | (0.005) | (0.005) | (0.008) | (0.004) | (0.005) | (0.006) | (0.004) |
| Dairy | -0.039*** | **-1.128\*\*\*** | -0.012*** | -0.006 | -0.024*** | 0.072*** | 0.048*** | 0.055*** | 0.013* | 0.071*** |
|  | (0.002) | **(0.005)** | (0.002) | (0.004) | (0.003) | (0.006) | (0.004) | (0.005) | (0.008) | (0.004) |
| Pulses | 0.015*** | -0.025*** | **-0.744\*\*\*** | -0.142*** | -0.169*** | 0.298*** | -0.070*** | -0.027*** | -0.002 | 0.037*** |
|  | (0.004) | (0.008) | **(0.013)** | (0.017) | (0.009) | (0.020) | (0.009) | (0.010) | (0.008) | (0.004) |
| Oils & spices | -0.005** | 0.019*** | -0.041*** | **-0.372\*\*\*** | 0.060*** | 0.070*** | 0.004 | -0.031*** | 0.016** | 0.075*** |
|  | (0.002) | (0.004) | (0.005) | **(0.008)** | (0.004) | (0.008) | (0.007) | (0.007) | (0.006) | (0.003) |
| F&V | 0.015*** | -0.018*** | -0.051*** | 0.036*** | **-0.646\*\*\*** | -0.126*** | -0.015*** | 0.004 | 0.006 | 0.006** |
|  | (0.002) | (0.004) | (0.003) | (0.004) | **(0.005)** | (0.006) | (0.004) | (0.004) | (0.005) | (0.003) |
| Animal meats, fresh | 0.008 | 0.131*** | 0.135*** | 0.085*** | -0.234*** | **-0.624\*\*\*** | 0.266*** | -0.008 | -0.019 | 0.025*** |
|  | (0.006) | (0.011) | (0.010) | (0.013) | (0.010) | **(0.033)** | (0.013) | (0.014) | (0.013) | (0.007) |
| Packaged proc. foods | -0.039*** | 0.133*** | -0.062*** | -0.024 | -0.060*** | 0.416*** | **-0.917\*\*\*** | 0.005 | 0.034*** | -0.011** |
|  | (0.004) | (0.012) | (0.007) | (0.017) | (0.011) | (0.021) | **(0.011)** | (0.009) | (0.007) | (0.005) |
| Sweets | 0.017** | 0.184*** | -0.036*** | -0.141*** | -0.014 | -0.022 | 0.004 | **-0.437\*\*\*** | 0.031*** | -0.073*** |
|  | (0.007) | (0.019) | (0.009) | (0.021) | (0.015) | (0.027) | (0.011) | **(0.018)** | (0.010) | (0.008) |
| SSBs | 0.014 | 0.067 | -0.020 | 0.050 | -0.005 | -0.105* | 0.083*** | 0.061*** | **-1.214\*\*\*** | 0.093*** |
|  | (0.018) | (0.062) | (0.018) | (0.045) | (0.035) | (0.056) | (0.020) | (0.022) | **(0.018)** | (0.018) |
| Non-SSBs | -0.000 | 0.196*** | 0.020*** | 0.152*** | 0.017** | 0.043*** | -0.002 | -0.045*** | 0.037*** | **-1.476\*\*\*** |
|  | (0.004) | (0.011) | (0.003) | (0.007) | (0.007) | (0.010) | (0.005) | (0.006) | (0.006) | **(0.009)** |

Notes: Estimated based on NSS Household Consumption Expenditure survey 2022-23 [16] and using Deaton (1988)'s quality-adjusted Almost Ideal Demand System model [21]. Own-price elasticities and their standard errors are denoted in bold. Bootstrapped standard errors after 300 replications. Pack.: Packaged; proc.: processed; SSBs: sugar-sweetened beverages.

We provide sensitivity analyses estimating the model using a broader beverage category, i.e., combining SSBs and non-SSBs, to reduce the number of clusters dropped due to non-purchases (**Table C2** in **S3 Supporting Information**). We also perform a sensitivity analysis in which we relax our assumption of unitary unit values for the *numeraire* group and instead assign the non-food consumer price index by state, quarter, and sector (i.e., rural or urban) (**Table C3** in **S3 Supporting Information**) [38]. Elasticity estimates from these alternative specifications are consistent.

### Immediate policy impacts

Depending on the NPM used, HFSS foods account for 9.9% (FSSAI) or 11.4% (WHO SEARO) of total baseline energy intakes by Indians (**Table A6** in **S1 Supporting Information**). Most of the items in the sweets and packaged processed food groups are HFSS. The difference between the two approaches is primarily driven by the extensive list of items excluded from the proposed FOPNL by FSSAI [25]. Immediate policy impacts on nutrient intake are presented in **Figure A4** in **S1 Supporting Information**. **Figures B1**, **B4**, and **B7** in **S2 Supporting Information** show the results for the sensitivity scenarios.

All scenarios would lead to a minor increase in expenditure for all income groups, from 0.6% (95% CI: 0.1%,1.1%) for low-income households for scenario 1 to 1.3% (95% CI: 0.1%, 2.5%) for high-income households for scenario 2 (**Figure A5** in **S1 Supporting Information**). These results are mainly driven by a significant decrease in HFSS food purchases – particularly, packaged processed foods (high absolute price elasticity of −0.92) – and substitutions from HFSS foods to dairy and fresh animal meats. Results are similar for sensitivity scenarios (passthrough, reformulation, and subsidy) (**Figures B2**, **B5**, and **B8** in **S2 Supporting Information**).

All scenarios would significantly increase tax revenue from foods and beverages, from +51.3% for scenario 1 (95% CI: 49.8%, 52.8%) to +110.0% for scenario 4 (95% CI: 105.3%, 114.7%) (**Figure A6** in **S1 Supporting Information**). Results are similar for sensitivity passthrough and reformulation scenarios (**Figures B3** and **B6** in **S2 Supporting Information**). Tax revenue is predictably lower for the sensitivity scenario with GST 0% on F&V and pulses (**Figure B9** in **S2 Supporting Information**). Behaviourally, these results are driven by a quantity (substitution) effect, with consumers shifting away from HFSS items toward dairy and fresh animal meats, and a tax increase effect, as most HFSS items belong to the packaged processed food and sweets groups, whose demand is price inelastic. The latter implies that the percentage fall in quantity demanded is smaller than the tax-induced percentage increase in price, thus increasing tax revenue. Regarding the magnitude of these results, most HFSS items are taxed at 12% or 18% at baseline; therefore, our scenarios imply a 10- to 28-percentage-point increase, which represents a significant 43% to 122% relative rise in rates for these items. The tax base is also concentrated in these tax brackets: HFSS items account for 85.3%, 93.1%, and 100% of baseline energy intake taxed at 12%, 18%, and 28%, respectively, but only 1.6% and 1.1% at the 0% and 5% rates, respectively. Most non-HFSS items are zero-rated or taxed at 5%. (**Figure A2** in **S1 Supporting Information**).

### Long-term policy impacts

Figure 3 presents the upper bounds of reduction in sodium intake and BMI under scenarios 1 and 2 compared to no policy change over time, by income group (lower bound results available in **Figure A7** in **S1 Supporting Information**). The reduction in daily sodium intake is persistent and stable over the 30 years, while the decrease in BMI tends to be stable after 4 years of policy implementation. Scenario 2 has greater impact on sodium intake and BMI than scenario 1. Under scenario 2, the population average reduction in daily sodium intake is approximately 45.8mg (95% CI: −45.9, −45.7) (−1.62% of baseline sodium intake; 95% CI: −1.623%, −1.615%), ranging from 31.8mg (95% CI: −32.1, −31.6) in the low-income group (−1.24%; 95% CI: −1.25%, −1.23%) to 66.3mg (95% CI: −66.7, −66.0) in the high-income group (−2.07%; 95% CI: −2.08%, −2.06%). After 30 years of policy implementation, the average reduction in BMI of the entire population is 0.1705kg/m^2^ (95% CI: −0.1709, −0.1700) (−0.796% of baseline BMI; 95% CI: −0.798%, −0.793%) under scenario 2, ranging from 0.137kg/m^2^ (95% CI: −0.138, −0.136) in the low-income group (−0.663%; 95% CI: −0.668%, −0.659%) to 0.219 kg/m^2^ (95% CI: −0.220, −0.218) in the high-income group (−0.972%; 95% CI: −0.978%, −0.967%). Sodium and BMI reductions are consistently larger for the high-income group compared to the lower-income group. Results under scenarios 3 and 4 are reported in **Figures A8** and **A9** in **S1 Supporting Information**.

**Figure 3.**
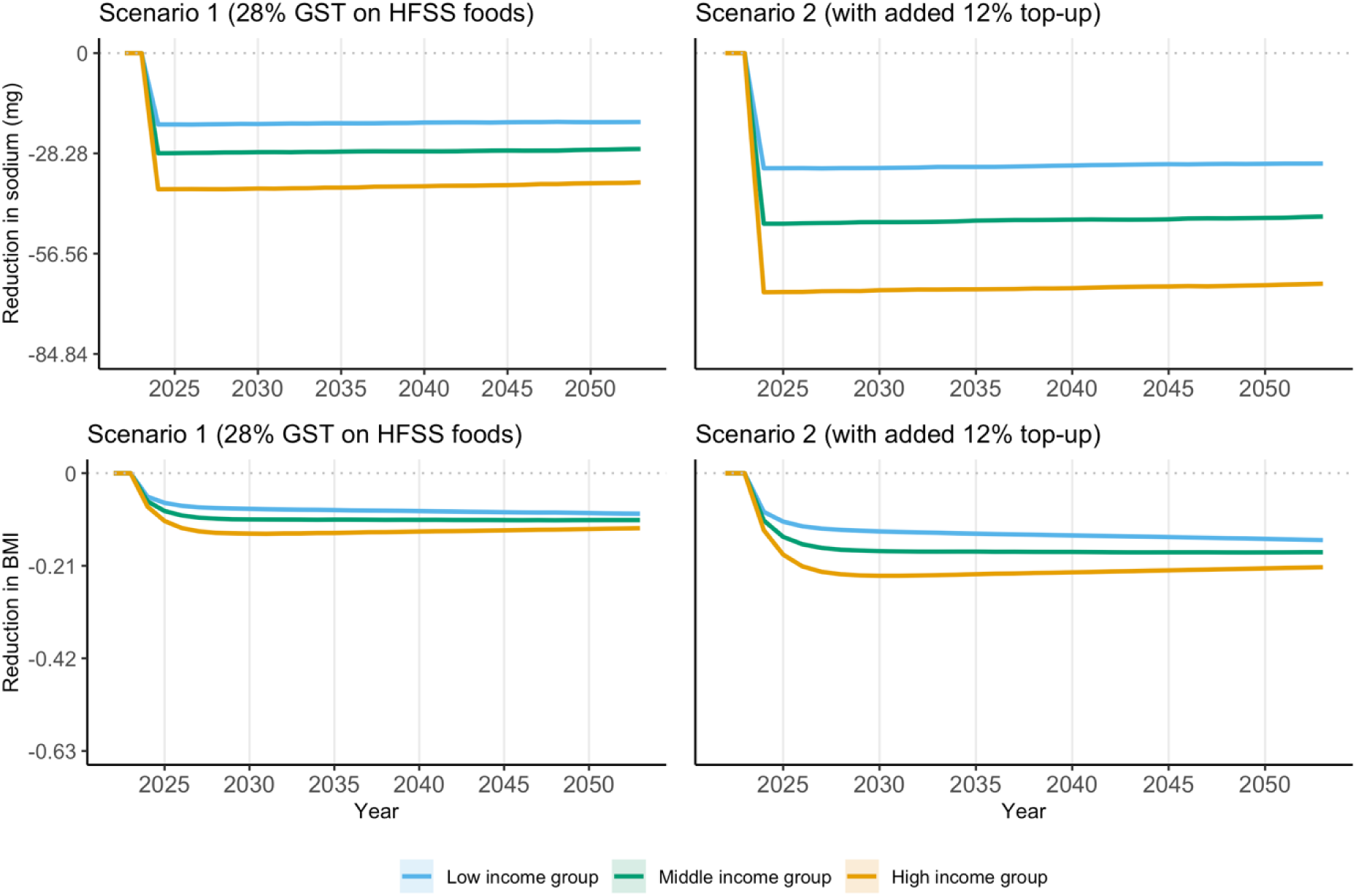
Reduction in sodium and body mass index compared to no policy change, by income group, scenarios 1 and 2. Notes: Upper bound estimates (lower bound estimates can be found in **Figure A7** in **S1 Supporting Information**). 95% confidence interval reported as shaded area. In the top two plots on sodium, a unit of 28.28 in y-axis corresponds to 1% of adjusted baseline sodium intake (2,828mg after adjustment according to height and weight distributions). In the bottom two plots on BMI, a unit of 0.21 in y-axis corresponds to 1% of baseline average BMI (21.43kg/m^2^). Policy is introduced in 2024. Scenario 1: GST rate is increased to 28% for foods and beverages high in fat, sugar, and sodium (HFSS) based on the definition by the Food Safety and Standards Authority of India [25]; Scenario 2: adding a 12% top-up to the tax rate applied on HFSS foods and beverages in Scenario 1. BMI: body mass index; GST: Goods and Services Tax. HFSS: High in fat, sodium, and sugar.

A reduction in sodium intake and a decrease in BMI reduce the risk of five major diseases: IHD, CKD, stroke, diabetes, and asthma. Over 30 years, the cumulative reduction in the incidence of the five diseases in the entire population is 5.44 million (95% CI: −5.71, −5.17) cases and 9.23 million (95% CI: −9.54, −8.91) cases (equivalent to 0.18 million (95% CI: −0.17, −0.19) and 0.31 million (95% CI: −0.32, −0.30) cases per year on average) under scenarios 1 and 2, respectively (**Table A10** in **S1 Supporting Information**, corresponding lower bound estimates in **Table A11** in **S1 Supporting Information**). This corresponds to an average yearly reduction of 1.01% (95% CI: −1.06%, −0.96%) and 1.72% (95% CI: −1.78%, −1.66%) in the baseline incidence number of the five diseases in India (2019). As reported in **Table 3**, the high-income group has the largest cumulative reduction in incidence rate of the five diseases, with 915 cases per 100,000 population (95% CI: −972, −858) over 30 years under scenario 2. Among diseases, the largest reductions are observed in IHD and diabetes, which made up 7.8% and 2.5% of DALYs in India in 2019, respectively [30]. Corresponding lower bound estimates and results under scenarios 3 and 4 are reported in **Tables A12-A14** in **S1 Supporting Information**. In addition, a lower prevalence of hypertension is observed under all scenarios. In the 30^th^ year after policy implementation, the number of hypertension cases in India decrease by 7.07 million (95% CI: −7.66, −6.48) and 11.87 million (95% CI: −12.46, −11.28) under scenarios 1 and 2, respectively (**Figure A10** in **S1 Supporting Information**, −3.10% (95% CI: −3.36%, −2.84%) and −5.20% (95% CI: −5.46%, −4.94%) compared to the national prevalence number in India from National Family Health Survey, 2019-21; lower bound estimates in **Figure A11** in **S1 Supporting Information**).

**Table 3.**
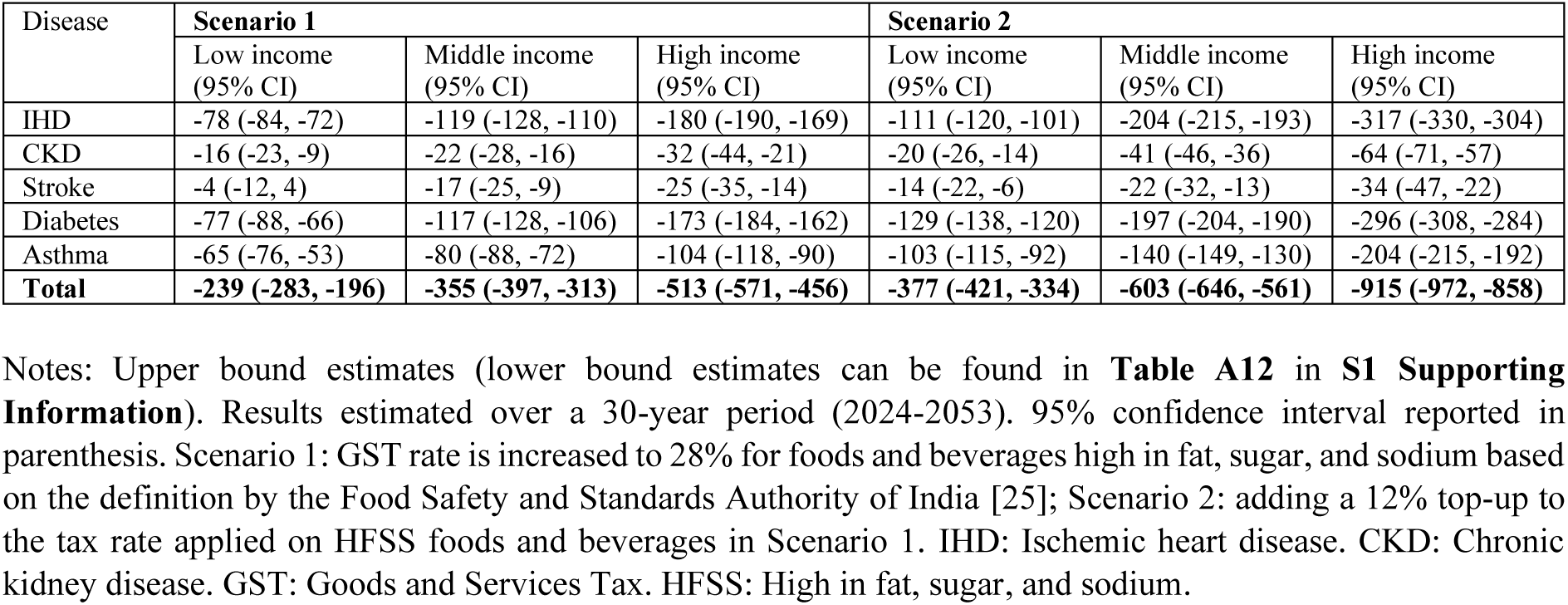
Cumulative reduction in disease incidence rate (cases per 100,000 population) compared to no policy change, by income group, scenarios 1 and 2.

| Disease | Scenario 1 |  |  | Scenario 2 |  |  |
| --- | --- | --- | --- | --- | --- | --- |
|  | Low income<br>(95% CI) | Middle income<br>(95% CI) | High income<br>(95% CI) | Low income<br>(95% CI) | Middle income<br>(95% CI) | High income<br>(95% CI) |
| IHD | -78 (-84, -72) | -119 (-128, -110) | -180 (-190, -169) | -111 (-120, -101) | -204 (-215, -193) | -317 (-330, -304) |
| CKD | -16 (-23, -9) | -22 (-28, -16) | -32 (-44, -21) | -20 (-26, -14) | -41 (-46, -36) | -64 (-71, -57) |
| Stroke | -4 (-12, 4) | -17 (-25, -9) | -25 (-35, -14) | -14 (-22, -6) | -22 (-32, -13) | -34 (-47, -22) |
| Diabetes | -77 (-88, -66) | -117 (-128, -106) | -173 (-184, -162) | -129 (-138, -120) | -197 (-204, -190) | -296 (-308, -284) |
| Asthma | -65 (-76, -53) | -80 (-88, -72) | -104 (-118, -90) | -103 (-115, -92) | -140 (-149, -130) | -204 (-215, -192) |
| **Total** | **-239 (-283, -196)** | **-355 (-397, -313)** | **-513 (-571, -456)** | **-377 (-421, -334)** | **-603 (-646, -561)** | **-915 (-972, -858)** |

Notes: Upper bound estimates (lower bound estimates can be found in **Table A12 in S1 Supporting Information**). Results estimated over a 30-year period (2024-2053). 95% confidence interval reported in parenthesis. Scenario 1: GST rate is increased to 28% for foods and beverages high in fat, sugar, and sodium based on the definition by the Food Safety and Standards Authority of India [25]; Scenario 2: adding a 12% top-up to the tax rate applied on HFSS foods and beverages in Scenario 1. IHD: Ischemic heart disease. CKD: Chronic kidney disease. GST: Goods and Services Tax. HFSS: High in fat, sugar, and sodium.

As a result, associated DALYs of IHD, CKD, stroke, diabetes, and asthma are reduced. Over 30 years, in the entire population, the cumulative reduction in DALYs is 10.36 million years (95% CI: −12.47, −8.25) and 18.98 million years (95% CI: −21.37, −16.58) under scenarios 1 and 2, respectively (**Table A10** in **S1 Supporting Information**, equivalent to 0.35 million years (95% CI: −0.42, −0.27) and 0.63 million years (95% CI: −0.71, −0.55) per year on average). The high-income group has the largest cumulative reduction in DALYs rate, up to 2,059 DALYs per 100,000 population (95% CI: −2553, −1564) under scenario 2 (Figure 4). Meanwhile, the policy scenarios are associated with a decrease in total health expenditure of US$10.7 billion (95% CI: −11.3, −10.1) and US$18.0 billion (95% CI: −18.7, −17.3) over 30 years under scenarios 1 and 2, respectively (**Table 4**, equivalent to US$357 million (95% CI: −376, −337) and US$601 million (95% CI: −624, −578) per year on average). Lower bound estimates and results under scenarios 3 and 4 are reported in **Tables A15-A17** in **S1 Supporting Information**, as well as in **Figures A12-A14** in **S1 Supporting Information**.

**Figure 4.**
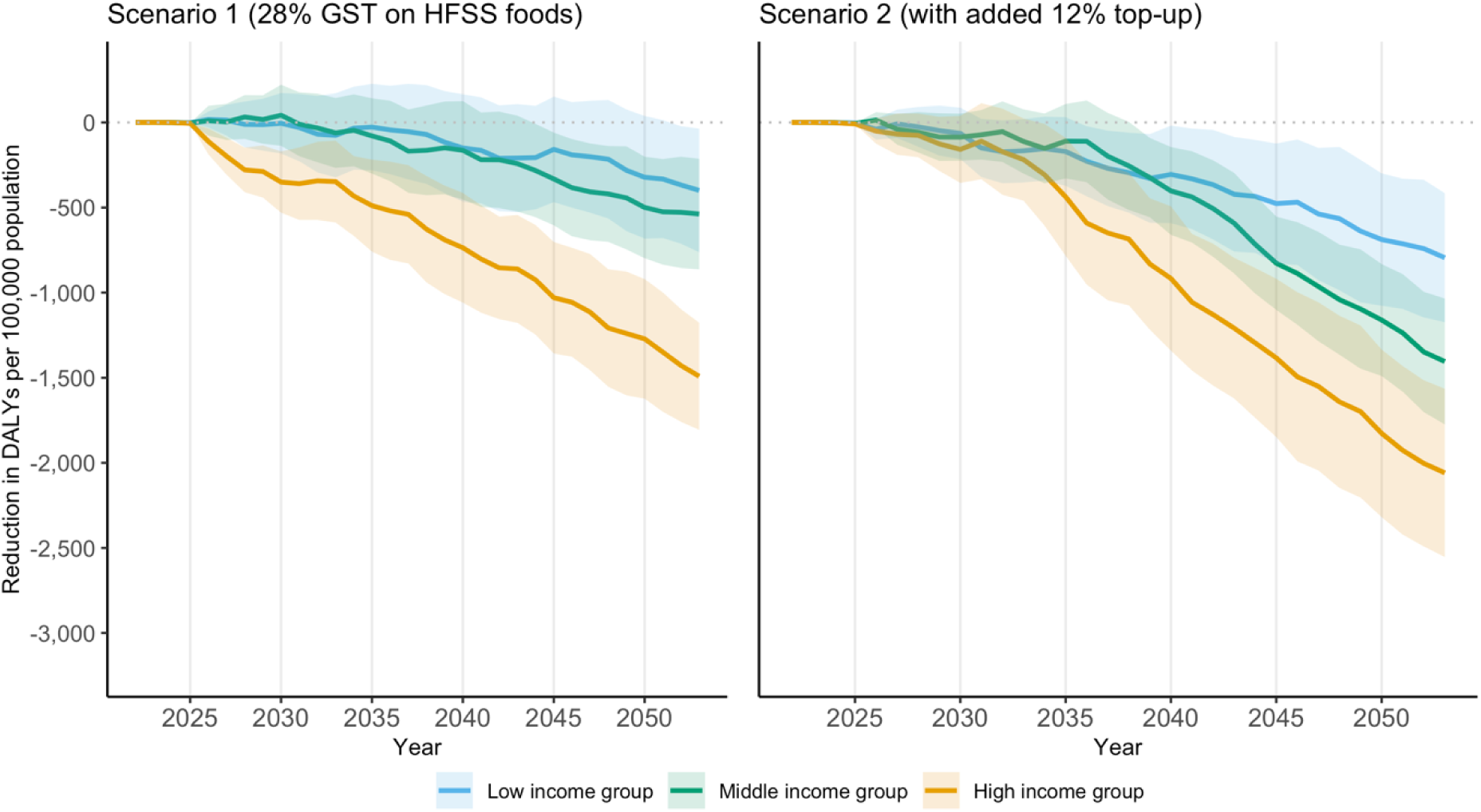
Cumulative reduction in DALYs per 100,000 population compared to no policy change, by income group, scenarios 1 and 2. Notes: Cumulative reduction in DALYs per 100,000 population of five key diseases including ischemic heart disease, chronic kidney disease, stroke, diabetes and asthma over 2022-2053. Upper bound estimates (lower bound estimates can be found in **Figure A12** in **S1 Supporting Information**). 95% confidence interval reported as shaded area. Policy is introduced in 2024. Scenario 1: GST rate is increased to 28% for foods and beverages high in fat, sugar, and sodium based on the definition by the Food Safety and Standards Authority of India [25]; Scenario 2: adding a 12% top-up to the tax rate applied on HFSS foods and beverages in Scenario 1. DALYs: Disability-Adjusted Life Years. GST: Goods and Services Tax. HFSS: High in fat, sugar, and sodium.

**Table 4.** Summary of main results, scenarios 1 and 2.

|  |  | Immediate effect at implementation |  | Cumulative results for the five key diseases over 2024-2053 |  |  |
| --- | --- | --- | --- | --- | --- | --- |
|  |  | Change in HH expenditure on foods and beverages (%) | Change in tax revenue from foods and beverages (%) | Change in incidence rate per 100,000 population | Change in DALYs per 100,000 population | Change in total health expenditure (USD billion) |
| Scenario 1 | Low income | +0.6% (+0.1%, +1.1%) | +51.3% (+49.8%, +52.8%) | -239 (-283, -196) | -400 (-763, -36) | -10.7 (-11.3, -10.1) |
|  | Middle income | +0.6% (+0.1%, +1.1%) |  | -355 (-397, -313) | -538 (-863, -213) |  |
|  | High income | +0.8% (+0.2%, +1.4%) |  | -513 (-571, -456) | -1492 (-1806, -1177) |  |
| Scenario 2 | Low income | +0.9% (+0.0%, +1.8%) | +92.0% (+88.2%, +95.7%) | -377 (-421, -334) | -795 (-1174, -416) | -18.0 (-18.7, -17.3) |
|  | Middle income | +1.0% (+0.0%, +1.9%) |  | -603 (-646, -561) | -1405 (-1775, -1035) |  |
|  | High income | +1.3% (+0.1%, +2.5%) |  | -915 (-972, -858) | -2059 (-2553, -1564) |  |

Notes: Upper bound estimates (lower bound estimates can be found in **Table A15 in S1 Supporting Information**). Results estimated over a 30-year period (2024-2053). 95% confidence interval reported in parenthesis. Scenario 1: GST rate is increased to 28% for foods and beverages high in fat, sugar, and sodium based on the definition by the Food Safety and Standards Authority of India [25]; Scenario 2: adding a 12% top-up to the tax rate applied on HFSS foods and beverages in Scenario 1. DALYs: Disability-adjusted life years. GST: Goods and Services Tax. HFSS: High in fat, sugar, and sodium. HH: household. USD: US dollar 2024.

## Discussion

Applying a GST rate of 40% to HFSS items, based on the draft FSSAI HFSS definition, could reduce annual average disease incidence by up to 1.72% (95% CI: −1.78%, −1.66%). Over 30 years, it could prevent as many as 18.98 million (95% CI: −21.37, −16.58) DALYs and avert up to US$18.0 billion (95% CI: −18.7, −17.3) in total health expenditure on IHD, CKD, stroke, diabetes and asthma (upper bound estimates). This represents an average reduction of approximately 0.6% of total health expenditure, equivalent to US$0.44 per capita and 0.02% of GDP in India per year. A 40% GST rate on HFSS items is also associated with an immediate increase in tax revenue from foods and beverages of 92.0% (95% CI: 88.2%, 95.7%) and a minor increase in total household expenditure on foods and beverages for all income groups (ranging from +0.9% (95% CI: 0.0%, 1.8%) for low-income to +1.3% for high-income (95% CI: 0.1%, 2.5%)).

The largest absolute health gains accrue to higher-income individuals because their baseline intake of HFSS calories and sodium is greater (**Tables A3, A4,** and **A5** in **S1 Supporting Information**). Nevertheless, health benefits remain significant for low-income individuals, who face higher odds of incidence and intensity of catastrophic out-of-pocket health expenditure [39]. The modelled reductions in morbidity and service use lead to a decline in total health expenditure and thus out-of-pocket expenditure - representing over 40% of total health spending in the country [34]-, which could offset the rise in food and beverage expenditure, particularly for the poorest, eliminating or mitigating the net financial burden.

Slightly higher health and economic benefits are obtained under a scenario using the WHO SEARO NPM to define HFSS foods (scenarios 3 and 4, **Appendix A** in **S1 Supporting Information**), as fewer items are exempted (**Table A2** and **Table A6** in **S1 Supporting Information**). Under both the draft FSSAI and the WHO SEARO NPM HFSS food definition, significantly stronger health and economic benefits are associated with a 40% GST rate (scenarios 2 and 4, i.e., including an additional 12% tax on HFSS food) than with a 28% rate. The difference between lower- and upper-bound estimates is small, suggesting the results are robust to a plausible income trend (**Appendix A** in **S1 Supporting Information**). Our results are robust to potential strategic industry tax avoidance, namely tax under-shifting to prices and reformulation (**Appendix B** in **S2 Supporting Information**).

This study is based on the latest available nationally representative household consumption survey data, with information for more than 250,000 households. By linking this data with nutritional information, it provides the most up-to-date energy and nutrient intake estimates across the Indian population. Additionally, it estimates the income and price elasticity of demand for ten food groups and three income strata, updating previous estimates based on earlier waves of this microdata from more than a decade ago. Up-to-date intake levels and elasticities are foundational inputs for projecting diet and nutrition outcomes and for simulating policy or market shocks (e.g., taxes, subsidies, income growth, inflation). This analysis utilises a novel microsimulation model, Health-GPS, to project health impacts. It simulates a large synthetic population of over 7 million individuals, representing 0.5% of the Indian population. The individual-level simulation allows us to capture heterogeneity in risk factor exposures and disease status. Health-GPS accounts for comorbidity, reflecting real-world disease patterns, and encompasses a wide range of diseases. Mortality from other causes is captured through residual mortality in Health-GPS.

Our analysis is not exempt from limitations. Our approach is a first-order attempt to individualise energy and nutrient intake from household-level data based on members’ age and sex and dietary guidelines. However, intra-household allocation of food and nutrients in India may be influenced by age and gender biases [40]. As our demand parameters are estimated at the household level, our analysis also assumes uniform relative consumption responses to the fiscal scenarios across household members, regardless of age or sex. Inherent to demand estimations based on cross-sectional data and rather aggregated food groups [9,37], we do not account for cross-price effects within such groups. Further disaggregation comes with a trade-off. It reduces the number of non-zero consumption observations per group, which undermines the reliability and identification of demand elasticities. Future research using product-level consumer panel or scanner data in India could overcome this issue [11]. Consistent with standard practice, we assume elasticity estimates are constant over time [13,14]. However, responsiveness can drift with income growth, urbanisation, demographics, and the food environment, such that holding elasticities fixed may bias long-run impacts [41].

Our microsimulation assumes no underlying time trend of diseases, which may be inconsistent with the observed increasing risks of non-communicable diseases associated with, for example, air pollution and climate change [42]. As a result, our absolute health and cost impacts estimates should be interpreted as conservative, because any unmodelled upward drift in baseline risk would scale up the number of cases and DALYs averted by a given proportional risk reduction. The analysis accounts for two pathways from diet to health, or two dietary risks: high sodium intake and high BMI (determined by energy imbalance). It does not consider the positive health impact of improving the source of energy (e.g., from F&V or pulses rather than HFSS foods) or increasing whole grain, F&V or legume intake [43]. Because these composition effects are not captured, our health impact estimates are likely conservative, particularly for policies that promote these foods (e.g., subsidy scenario zero-rating F&V and pulses) (**Figure B7** in **S2 Supporting Information**). A fuller treatment would require multi-risk modelling with joint intake distributions and covariance to avoid double-counting, which is not currently feasible with available data.

For ease of computability, we only account for the five diseases most attributable to excess sodium intake and high BMI in terms of DALYs in India. Our health benefit estimates therefore represent an underestimate of the potential effect. The same epidemiological data on disease incidence, prevalence, mortality, and remission are applied across income groups due to the dearth of granular data. Thus, differences in estimated absolute health benefits across income groups need to be interpreted with caution, as non-communicable diseases are not necessarily diseases of affluence in India [44]. In addition, the exposure-response functions (relative risks) used in this study are derived from global meta-analyses (e.g., Institute for Health Metrics and Evaluation Global Burden of Disease), whose underlying cohorts are disproportionately from high-income countries and typically assume generalizability across different settings. Restricted by data availability, total health expenditure is extrapolated from household expenditure on health, assuming an equal distribution of health expenditure between households and the government across diseases, following the methods adopted by WHO [45]. We did not fully propagate parameter and structural uncertainty through the model. Uncertainty was addressed by bootstrapping standard errors in our demand estimation, providing confidence intervals from 20 repeated simulation runs to capture stochastic variation, and reporting sensitivity scenario analyses. This is due to data limitations regarding the variance of risk ratio estimates and the underlying distribution of dietary risk factors. As such, our uncertainty intervals are likely conservative.

### Policy implications

The 2024 Economic Survey of India and the WHO take cognisance of the increasing consumption of HFSS foods in India and outline the need to promote healthy diets [46,47]. The introduction of the 28% GST and 12% Cess on caffeinated and sweetened aerated beverages (including SSBs) in 2017, which represent only a fraction of HFSS food intake, has shown limited impacts in reducing consumption [48]. However, the 2016-2017 Kerala state-wise indirect tax of 14.5% on burgers, pizzas, tacos, doughnuts, sandwiches, pasta and bread fillings sold in restaurants (repealed following the launch of the nationwide GST by the federal government to replace all state-level indirect taxes) was associated with reduced sales [49].

Adjusting taxes within the existing GST rate structure helps avoid multiple taxation schemes. This is particularly relevant given the latest policy debates around simplifying the Indian GST rate structure to reduce financial and administrative burdens and increase revenue collection efficiency. The most recent government proposal, enacted in September 2025, includes reducing the number of rates from five to three, removing the 12% and 28% GST rates. The 12% compensatory Cess is discontinued and a 40% GST rate on “sin” goods is introduced (equivalent to the previous 28% GST + 12% Cess), still covering only caffeinated and sweetened aerated beverages and some tobacco products [50]. Curbing HFSS food consumption may require a further expansion of the “sin” goods rate to all HFSS foods.

The tax administration challenges and companies’ compliance costs associated with taxing foods based on their nutritional content should be assessed, particularly in a setting with a significant informal market. While there is no robust independent evidence for negative macroeconomic impacts of diet-related fiscal policies [51], concerns may arise regarding the economic impact of the measure on the food industry and inflation. These potential challenges should be further studied.

Increasing GST rates on HFSS foods based on nutrient content may lead to significant dietary improvements and positive health effects, reducing health expenditure and increasing government tax revenue, with only a minor impact on household spending. The latter is important since it is estimated that at least two-thirds of the rural poor could not afford a recommended healthy diet in 2011 [52]. Our nutrient intake estimates based on the latest available microdata, robust demand elasticity estimates, and simulations of various GST scenarios contribute to informing future GST reforms to improve nutrition among the Indian population.

As an increasing number of countries contemplate taxing HFSS foods [6], the results of this study may also contribute to the broader global discussion on the use of this policy. Indeed, many countries share a similar differentiated value-added or sales tax system applied to foods and beverages, as seen in India, with an opportunity to align existing rates with nutritional objectives without adding new tiers. This approach has, for example, been supported by the European Parliament in its Farm to Fork strategy [53].

## Conclusions

India faces a rapid shift in diets towards HFSS foods. Tackling unhealthy diets is urgent. Other countries have highlighted the feasibility of regulating the marketing of unhealthy foods and beverages, the introduction of FOPNL, and shifts in relative prices to encourage significant changes towards healthier food purchases. Clear quantitative nutrient thresholds defining HFSS foods are crucial for the consistent development and implementation of these policies.

India introduced a higher 28% GST rate and a 12% Cess on caffeinated and sweetened aerated beverages, including SSBs, in July 2017 (equivalent to the “sin” goods 40% GST rate which will be applied from September 2025). Broader alignment of GST rates with nutrition and health objectives, targeting HFSS foods with higher rates, may represent the next critical step. This study has shown that it could result in meaningful reductions in the intake of nutrients of concern and significant long-run health benefits, yielding both lower health expenditure and higher tax revenue from foods and beverages while having a limited impact on household expenditure.

## Data Availability

Input data and statistical code as well as further information on the Health-GPS microsimulation model is available at https://imperialchepi.github.io/healthgps/. Data sources used in this analysis are available publicly: NSS Household Consumption Expenditure survey (2022-23) microdata at https://microdata.gov.in/NADA/index.php/catalog/224; IHME relative risks, disease prevalence, incidence and mortality data at https://ghdx.healthdata.org/ihme_data; UN World Population Prospects database at https://population.un.org/wpp/; and NCD-RisC data at https://www.ncdrisc.org/data-downloads.html.

## Acknowledgments

We thank Laura Cobb and the participants at two stakeholder meetings organized by the Institute for Economic Growth (IEG) in New Delhi, India, at the ‘Delivering for Nutrition in South Asia: Equity and Inclusion’ conference organized by the International Food Policy Research Institute (IFPRI) in Kathmandu, Nepal, and at the Indian Health Economics and Policy Association annual conference for their comments. We thank Belabess Alijadallah, Mahima Ghosh, and James Turner as well as his team members for their support with the calibration of the Health-GPS model. We thank Preeti Khanna and Abhishek Kumar for their assistance in matching food items with nutritional information and with GST rates, respectively. Finally, we thank Heather Lodge for administrative support.

## Authors’ contributions (based on CRediT taxonomy)

Conceptualization: FS, JO, MR, LS, MS, WJ. Data curation: MR, JZ. Formal Analysis: MR, JZ. Funding acquisition: FS, JO, LS, MS, WJ. Investigation: MR, JZ. Methodology: FS, MR, JZ, JO, DJL. Project administration: FS, JO, LS, MS, WJ. Resources: FS, JO, WJ. Software: JZ, JO, DJL. Supervision: FS, LS, MS, WJ. Validation: MR, JZ, JO. Visualization: MR, JZ, JO. Writing – original draft: MR, JZ. Writing – review & editing: MR, JZ, JO, DJL, MS, LS, WJ, FS.

## Supplementary information files

### Supporting information file 1

#### Appendix A. Additional tables and figures

Appendix A forms part of the revised submission.

Supplement to: Roche M, Zhu J, Olney J, Laydon DJ, Joe W, Sharma M, Steele L, Sassi F. Taxation of foods high in saturated fat, sugar, and sodium in India: A modelling study of health and economic impacts. Submitted after revision on 19 September 2025.

**Table A1.**
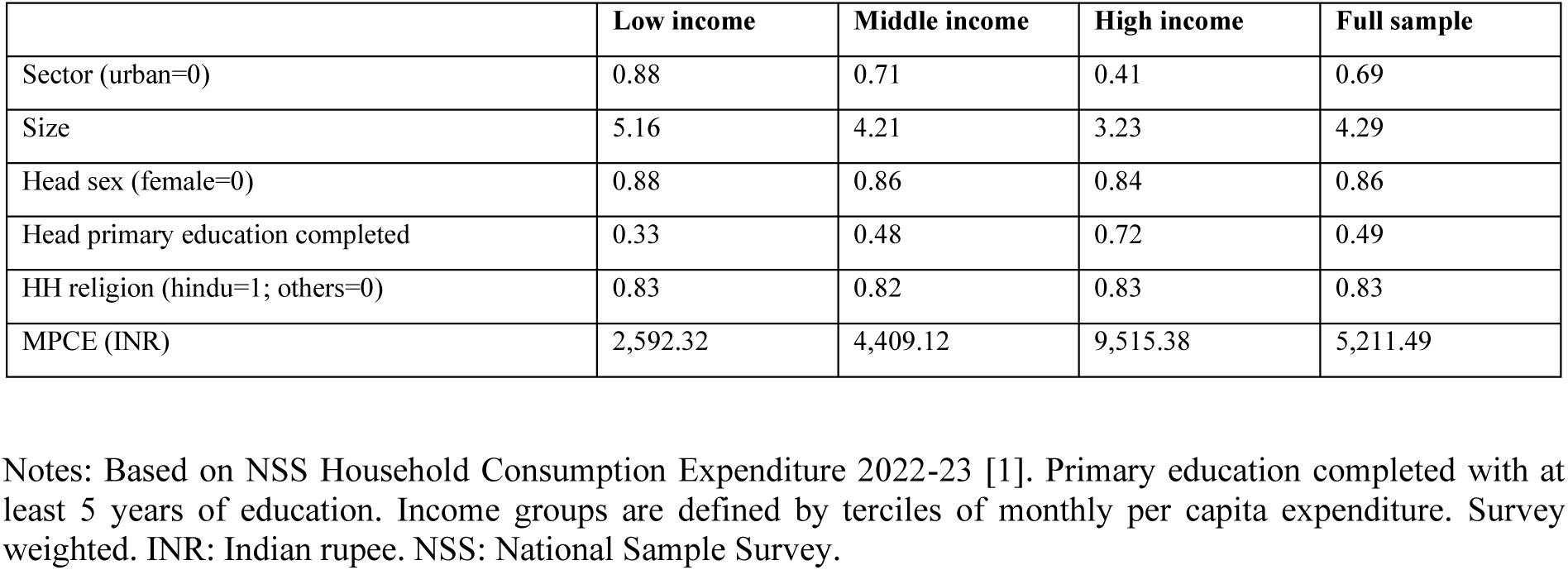
Household characteristics by income group.

|  | Low income | Middle income | High income | Full sample |
| --- | --- | --- | --- | --- |
| Sector (urban=0) | 0.88 | 0.71 | 0.41 | 0.69 |
| Size | 5.16 | 4.21 | 3.23 | 4.29 |
| Head sex (female=0) | 0.88 | 0.86 | 0.84 | 0.86 |
| Head primary education completed | 0.33 | 0.48 | 0.72 | 0.49 |
| HH religion (hindu=1; others=0) | 0.83 | 0.82 | 0.83 | 0.83 |
| MPCE (INR) | 2,592.32 | 4,409.12 | 9,515.38 | 5,211.49 |

Notes: Based on NSS Household Consumption Expenditure 2022-23 [1]. Primary education completed with at least 5 years of education. Income groups are defined by terciles of monthly per capita expenditure. Survey weighted. INR: Indian rupee. NSS: National Sample Survey.

**Table A2.** Draft Food Safety and Standards Authority of India definition of HFSS foods and WHO South East Asia Region nutrient profile model category-specific thresholds. <u>Food Safety and Standards Authority of India, Labelling & Display Amendment Draft Regulations 2022</u> “High fat, sugar, salt (HFSS) food means a processed food product which has high levels of saturated fat or total sugar or sodium. The declared values of these ingredients are such that the product; does not satisfy the value of energy (kcal) from total sugar less than 10 percent of total energy, or from saturated fat 10 percent of total energy, and sodium less than 1 mg/1 kcal.” In this analysis, we assume processed products to be items not included in Schedule IV Category III Solid Foods/Liquid Foods exempted from FOPNL under INR in the Labelling & Display Amendment Draft Regulations 2022 (44272/2022/REGULATION-FSSAI) [2]. <u>WHO South East Asia Region nutrient profile model</u> In this analysis, we define items as HFSS if nutrient content exceeds the values per 100g (or 100ml) listed below.

| Food category | Food category name | Total fat (g) | Saturated fat (g) | Total sugars (g) | Sodium (g) | Energy (kcal) |
| --- | --- | --- | --- | --- | --- | --- |
| 1 | Confectionery | 8.0 | NA | 6.0 | NA | 230 |
| 2 | Fine bakery wares | 8.0 | NA | 6.0 | 0.25 | 230 |
| 3 | Bread & ordinary bakery wares | 8.0 | NA | 6.0 | 0.25 | NA |
| 4 | Cereals | 12.0 | NA | 9.0 | 0.35 | NA |
| 5a | Ready-to-eat savouries: potato, cereal or starch-based | 8.0 | NA | NA | 0.25 | 230 |
| 5b | Ready-to-eat savouries: processed nuts | NA | NA | NA | 0.05 | NA |
| 5c | Ready-to-eat savouries: fish-based | NA | NA | 6.0 | 0.25 | 230 |
| 6a | Beverages: juices | NA | NA | 6.0 | NA | NA |
| 6b | Beverages: milk & dairy-based | 7.0 | NA | NA | NA | NA |
| 6c | Beverages: water-based flavoured drink | NA | NA | 2.0 | 0.3 | NA |
| 6d | Beverages: coffee, tea, herbal infusions | NA | NA | 2.0 | NA | NA |
| 6e | Beverages: cereal, grain, & tree nut-based | NA | NA | 6.0 | 0.2 | NA |
| 7 | Frozen dairy-based desserts & edible ices | 8.0 | NA | 12.0 | 0.1 | 230 |
| 8 | Curded dairy-based desserts | 7.0 | NA | 6.0 | 0.1 | 230 |
| 9 | Cheese & analogues | 20.0 | NA | NA | 0.6 | NA |
| 10 | Composite foods (prepared foods) | 8.0 | 3.5 | 9.0 | 0.35 | NA |
| 11 | Fats, oils, & fat emulsions | NA | 35.0 | NA | 0.1 | NA |
| 12 | Pasta, noodles, & like products | 3.0 | NA | NA | 0.25 | NA |
| 13 | Fresh & frozen meat, poultry, game, fish & seafood products | 15.0 | NA | NA | NA | NA |
| 14a | Processed meat, poultry, & game products | 8.0 | NA | NA | 0.4 | NA |
| 14b | Processed fish & sea food products | 8.0 | 3.0 | NA | 0.4 | NA |
| 15 | Fresh & frozen fruits, vegetables, & legumes | NA | NA | NA | NA | NA |
| 16 | Processed fruits & vegetables | NA | NA | NA | 0.4 | NA |
| 17 | Solid-form soybean products | 12.0 | NA | 5.0 | 0.1 | NA |
| 18 | Sauces, dips, & dressings | 12.0 | NA | 10.0 | 0.3 | NA |

Source: Food Safety and Standards Authority of India, Labelling & Display Amendment Draft Regulations 2022 (44272/2022/REGULATION-FSSAI) and WHO South East Asia Region nutrient profile model.
Notes: We omit the ‘Added sugars (g)’ column for the WHO South East Asia Region nutrient profile model as we lacked information on added sugars [3]. FOPNL: front-of-pack nutritional labelling; HFSS: high in fat, sugar, and sodium; INR: Indian Nutrition Rating; NA: not applicable; WHO: World Health Organization.

**Figure A1.**
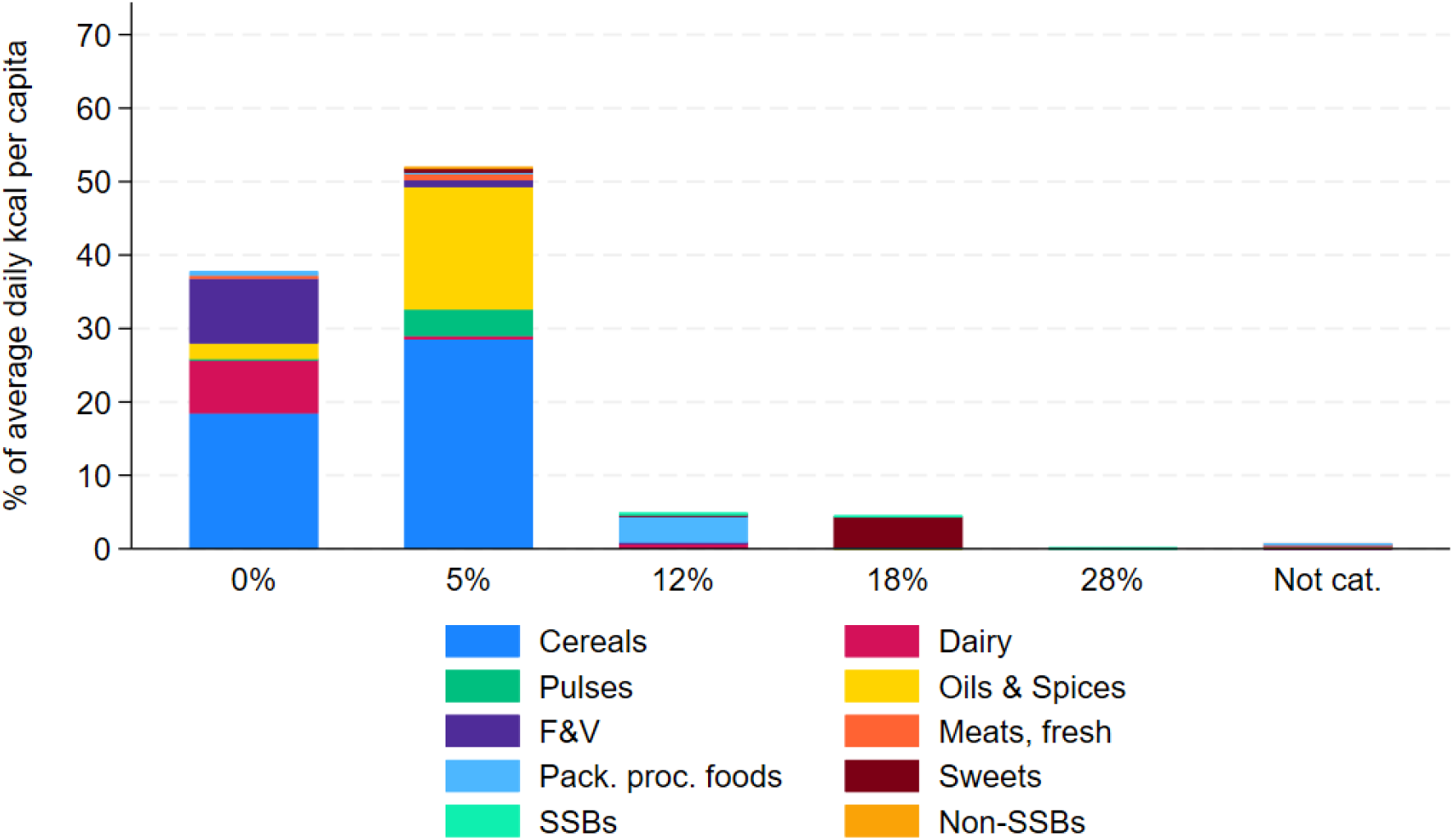
Baseline distribution of average daily per capita energy intake by GST rate, by food group, full sample. Notes: Based on NSS Household Consumption Expenditure 2022-23 [1]. On-trade food items are excluded from this analysis, i.e., NSS category ‘served processed food’. All non-PDS cereals, fresh meat/fish, arhar, khesari, curd, and honey are assumed pre-packaged (GST rates: not pre-packaged 0% / pre-packaged 5%). Tea and coffee cups assumed GST rate 5%, i.e., service GST in non-air-conditioned restaurants/bars. Survey-weighted. Pack.: packaged; proc.: processed; F&V: fruits & vegetables; NSS: National Sample Survey; SSBs: sugar-sweetened beverages. GST: Goods and Services Tax; PDS: Public Distribution System. Not. cat.: NSS items defined as ‘other’ in their respective category, thus their GST rate is assumed as the quantity-weighted average GST rate among items in the category and not categorized in this figure.

**Figure A2.**
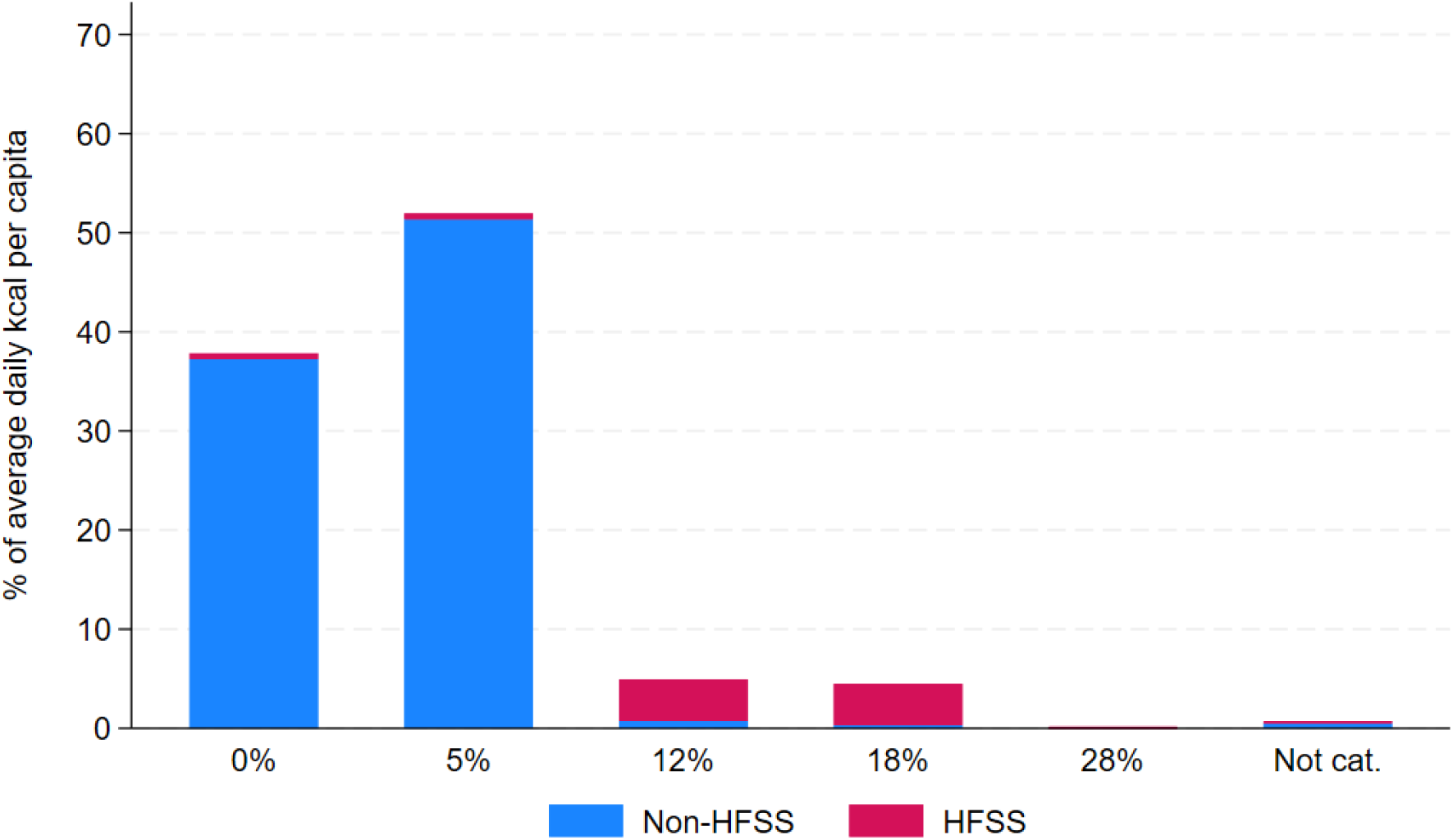
Baseline distribution of total energy intake by GST rate, by HFSS status, full sample. Notes: Based on NSS Household Consumption Expenditure 2022-23 [1]. HFSS definition based on the FSSAI Labelling & Display Amendment Draft Regulations, 2022 (44272/2022/REGULATION-FSSAI) [2]. On-trade food items are excluded from this analysis, i.e., NSS category ‘served processed food’. All non-PDS cereals, fresh meat/fish, arhar, khesari, curd, and honey are assumed pre-packaged (GST rates: not pre-packaged 0% / pre-packaged 5%). Tea and coffee cups assumed GST rate 5%, i.e., service GST in non-air-conditioned restaurants/bars. Survey-weighted. FSSAI: Food Safety and Standards Authority of India. GST: Goods and Services Tax; HFSS: High in fat, salt, and sugar; NSS: National Sample Survey; PDS: Public Distribution System. Not. cat.: NSS items defined as ‘other’ in their respective category, thus their GST rate is assumed as the quantity-weighted average GST rate among items in the category and not categorized in this figure.

**Table A3.**
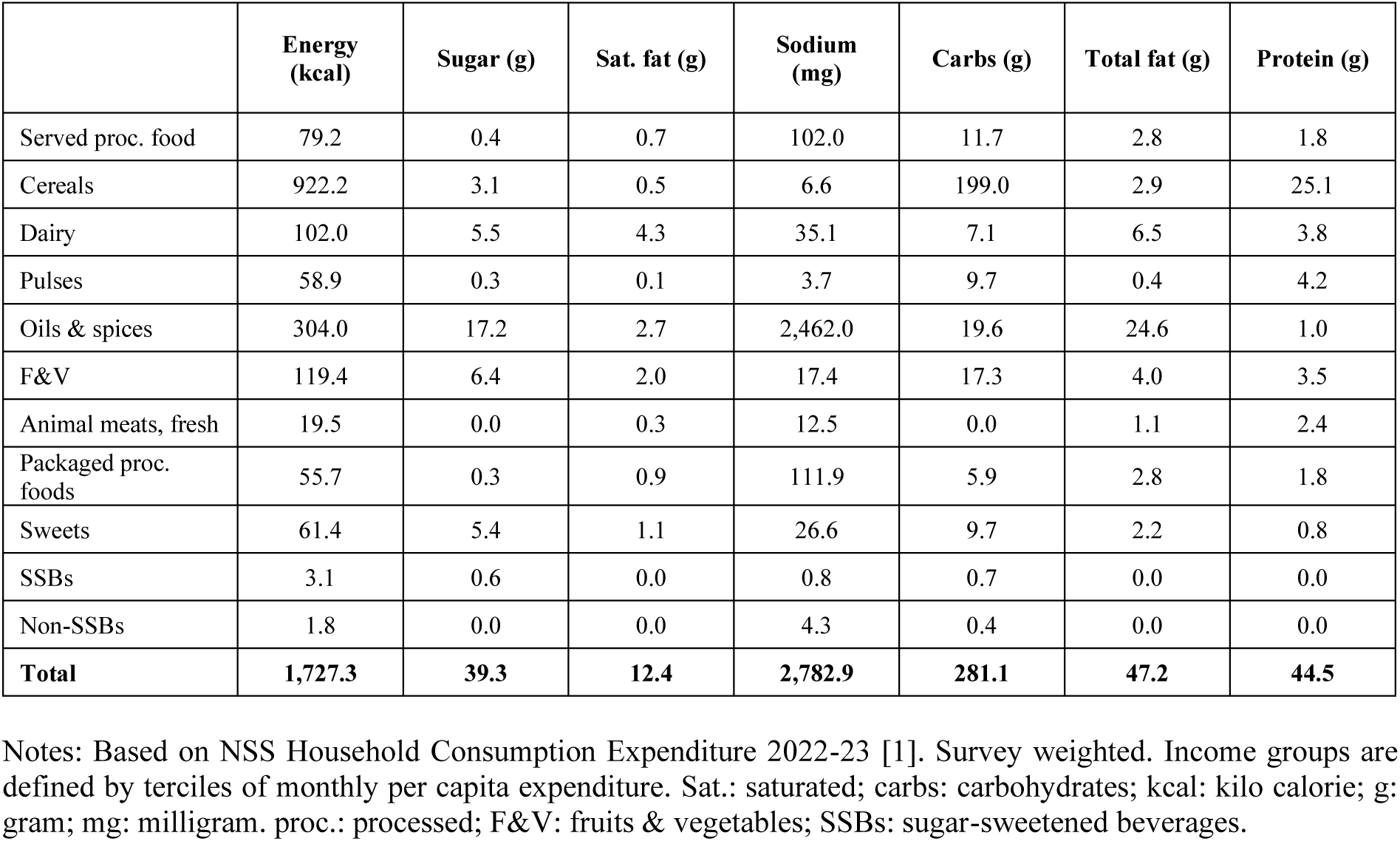
Estimated average daily per capita energy and nutrient intake, low-income group.

**Table A4.** Estimated average daily per capita energy and nutrient intake, middle-income group.

|  | Energy (kcal) | Sugar (g) | Sat. fat (g) | Sodium (mg) | Carbs (g) | Total fat (g) | Protein (g) |
| --- | --- | --- | --- | --- | --- | --- | --- |
| Served proc. food | 88.9 | 0.5 | 0.8 | 114.1 | 12.6 | 3.4 | 2.1 |
| Cereals | 926.2 | 3.2 | 0.5 | 6.8 | 199.5 | 3.0 | 25.3 |
| Dairy | 168.6 | 8.9 | 7.1 | 59.3 | 11.4 | 10.9 | 6.2 |
| Pulses | 72.9 | 0.3 | 0.1 | 4.6 | 11.9 | 0.5 | 5.3 |
| Oils & spices | 378.4 | 21.7 | 3.9 | 2,747.8 | 24.7 | 30.4 | 1.4 |
| F&V | 191.1 | 9.8 | 5.0 | 21.4 | 22.3 | 9.2 | 4.9 |
| Animal meats, fresh | 27.8 | 0.0 | 0.4 | 17.8 | 0.1 | 1.5 | 3.4 |
| Packaged proc. foods | 84.2 | 0.6 | 1.3 | 173.4 | 9.2 | 4.0 | 2.9 |
| Sweets | 85.2 | 7.7 | 1.5 | 37.1 | 13.5 | 3.0 | 1.1 |
| SSBs | 9.9 | 1.8 | 0.0 | 2.5 | 2.2 | 0.1 | 0.1 |
| Non-SSBs | 2.6 | 0.0 | 0.0 | 6.6 | 0.6 | 0.0 | 0.0 |
| **Total** | **2,035.9** | **54.5** | **20.6** | **3,191.3** | **308.1** | **65.9** | **52.7** |

Notes: Based on NSS Household Consumption Expenditure 2022-23 [1]. Survey weighted. Income groups are defined by terciles of monthly per capita expenditure. Sat.: saturated; carbs: carbohydrates; kcal: kilo calorie; g: gram; mg: milligram. proc.: processed; F&V: fruits & vegetables; SSBs: sugar-sweetened beverages.

**Table A5.** Estimated average daily per capita energy and nutrient intake, high-income group.

|  | Energy (kcal) | Sugar (g) | Sat. fat (g) | Sodium (mg) | Carbs (g) | Total fat (g) | Protein (g) |
| --- | --- | --- | --- | --- | --- | --- | --- |
| Served proc. food | 151.8 | 1.3 | 1.8 | 200.7 | 18.9 | 6.5 | 4.5 |
| Cereals | 842.2 | 2.8 | 0.4 | 6.3 | 181.6 | 2.7 | 23.0 |
| Dairy | 231.5 | 11.9 | 9.9 | 88.1 | 15.2 | 15.2 | 8.4 |
| Pulses | 85.2 | 0.4 | 0.1 | 5.5 | 13.9 | 0.5 | 6.2 |
| Oils & spices | 418.2 | 23.1 | 5.1 | 2,785.0 | 26.6 | 33.9 | 1.6 |
| F&V | 293.7 | 14.4 | 9.8 | 24.9 | 28.4 | 17.1 | 6.5 |
| Animal meats, fresh | 36.5 | 0.0 | 0.5 | 23.4 | 0.1 | 2.0 | 4.5 |
| Packaged proc. foods | 137.8 | 1.1 | 2.2 | 294.3 | 15.5 | 6.3 | 4.8 |
| Sweets | 137.9 | 11.9 | 2.6 | 61.8 | 21.2 | 5.1 | 1.8 |
| SSBs | 26.1 | 4.7 | 0.1 | 5.5 | 5.8 | 0.1 | 0.4 |
| Non-SSBs | 3.7 | 0.0 | 0.0 | 9.2 | 0.8 | 0.0 | 0.1 |
| **Total** | **2,364.5** | **71.5** | **32.5** | **3,504.7** | **328.0** | **89.5** | **61.7** |

Notes: Based on NSS Household Consumption Expenditure 2022-23 [1]. Survey weighted. Income groups are defined by terciles of monthly per capita expenditure. Sat.: saturated; carbs: carbohydrates; kcal: kilo calorie; g: gram; mg: milligram. proc.: processed; F&V: fruits & vegetables; SSBs: sugar-sweetened beverages.

**Figure A3.**
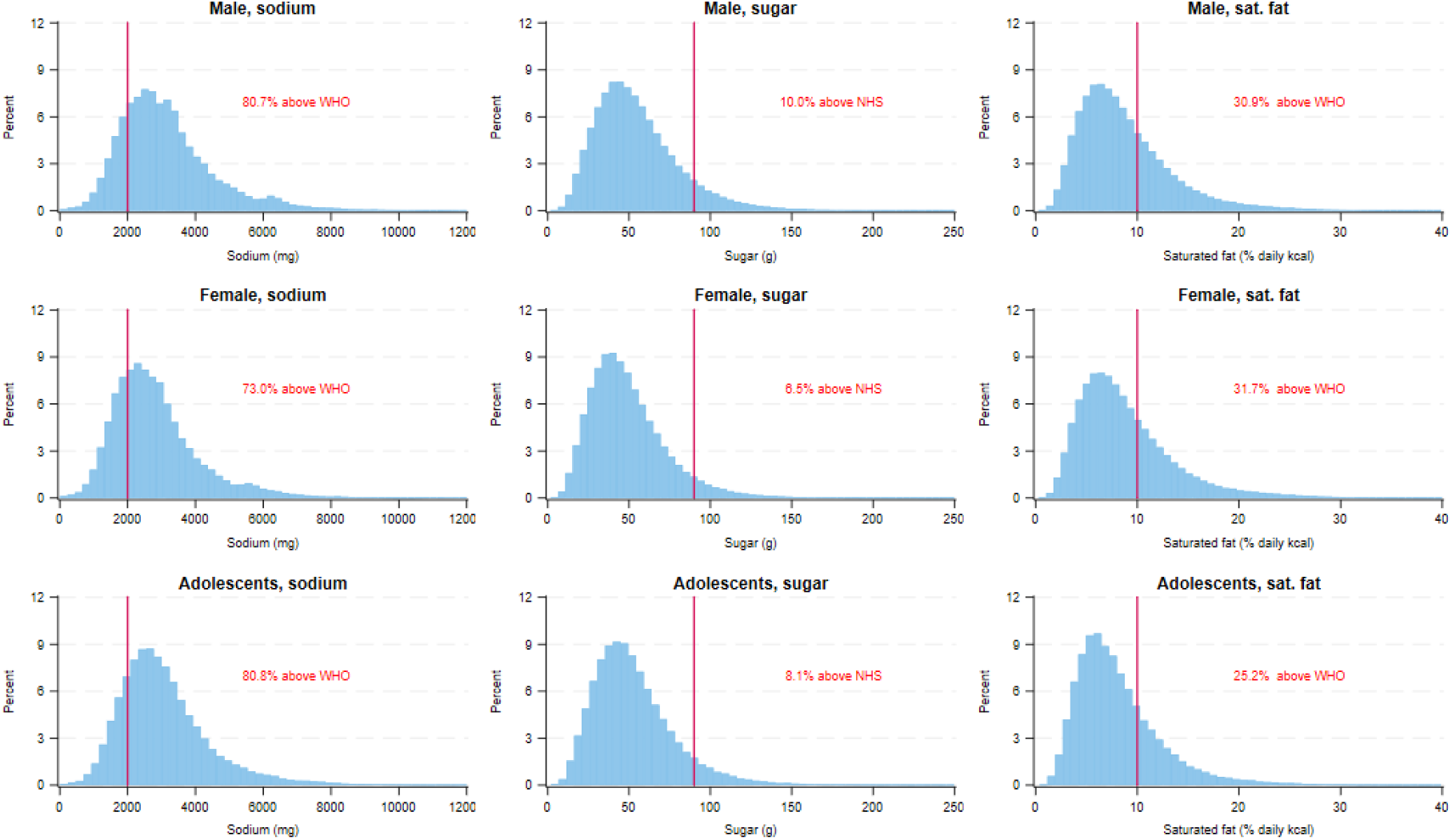
Estimated daily distribution of sodium, sugar, and saturated fat intake per capita compared to daily recommendations, for adult men, adult women, and adolescents (10-18 years old) Notes: Based on NSS Household Consumption Expenditure 2022-23 [1]. NHS: National Health Service, here referring to the NHS daily recommendation for total sugar intake in the United Kingdom (90g), which used instead of the WHO daily recommendation for free sugars as we missed information on free sugars; Sat.: saturated; WHO: World Health Organization, here referring to the WHO daily recommendations for the intake of sodium and saturated fat (no more than 2,000mg for sodium and 10% of total energy intake for saturated fat); kcal: kilo calorie; g: gram; mg: milligram.

**Table A6.** The proportion of total energy intake from the off-trade sector defined as high in fat, sugar, and sodium, full sample.

|  | **FSSAI HFSS** | **WHO SEARO NPM** |
| --- | --- | --- |
| Cereals | 0.000 | 0.000 |
| Dairy | 0.005 | 0.137 |
| Pulses | 0.000 | 0.000 |
| Oils & spices | 0.000 | 0.017 |
| F&V | 0.053 | 0.000 |
| Animal meats, fresh | 0.000 | 0.000 |
| Packaged proc. foods | 0.956 | 0.956 |
| Sweets | 0.911 | 0.998 |
| SSBs | 0.752 | 1.000 |
| Non-SSBs | 0.000 | 0.000 |
| **Total** | **0.099** | **0.114** |

Notes: On-trade food items are excluded from this analysis, i.e., NSS category ‘served processed food’. Survey-weighted. FSSAI HFSS: definition of foods and beverages high in fat, sugar, and sodium by the Food Safety and Standards Authority of India [2]; WHO SEARO NPM: items with nutrient content above at least one of the respective thresholds set by the World Health Organization South East Asia Region nutrient profile model [3]. proc.: processed; F&V: fruits & vegetables; HFSS: high in fat, sodium, and sugar; NSS: National Sample Survey; SSBs: sugar-sweetened beverages.

**Table A7.** Estimated average price elasticities, low-income group.

|  | Cereals | Dairy | Pulses | Oils & spices | F&V | Animal meats, fresh | Packaged proc. foods | Sweets | SSBs | Non-SSBs |
| --- | --- | --- | --- | --- | --- | --- | --- | --- | --- | --- |
| Cereals | **-0.847\*\*\*** | -0.052*** | 0.003 | -0.029*** | 0.050*** | 0.032*** | -0.005 | 0.017*** | 0.029*** | -0.001 |
|  | **(0.004)** | (0.007) | (0.003) | (0.006) | (0.006) | (0.007) | (0.004) | (0.005) | (0.005) | (0.004) |
| Dairy | -0.048*** | **-1.070\*\*\*** | -0.034*** | -0.063*** | -0.049*** | 0.090*** | 0.042*** | 0.036*** | 0.034*** | 0.045*** |
|  | (0.004) | **(0.009)** | (0.004) | (0.006) | (0.005) | (0.008) | (0.006) | (0.007) | (0.009) | (0.005) |
| Pulses | 0.009** | -0.054*** | **-0.801\*\*\*** | -0.066*** | -0.118*** | 0.188*** | -0.059*** | -0.013 | -0.004 | 0.030*** |
|  | (0.004) | (0.010) | **(0.015)** | (0.018) | (0.012) | (0.020) | (0.009) | (0.010) | (0.007) | (0.004) |
| Oils & spices | -0.007*** | -0.003 | -0.019*** | **-0.507\*\*\*** | 0.043*** | 0.042*** | -0.008 | -0.005 | 0.014*** | 0.044*** |
|  | (0.002) | (0.005) | (0.005) | **(0.008)** | (0.005) | (0.008) | (0.005) | (0.005) | (0.004) | (0.003) |
| F&V | 0.014*** | -0.013*** | -0.045*** | 0.018*** | **-0.684\*\*\*** | -0.111*** | -0.017*** | -0.008 | 0.004 | 0.002 |
|  | (0.002) | (0.005) | (0.004) | (0.005) | **(0.006)** | (0.007) | (0.005) | (0.005) | (0.005) | (0.003) |
| Animal meats, fresh | 0.002 | 0.154*** | 0.086*** | 0.020 | -0.226*** | **-0.552\*\*\*** | 0.249*** | -0.036*** | -0.015 | 0.017** |
|  | (0.006) | (0.013) | (0.011) | (0.016) | (0.012) | **(0.032)** | (0.013) | (0.014) | (0.013) | (0.007) |
| Packaged proc. foods | -0.033*** | 0.126*** | -0.077*** | -0.093*** | -0.095*** | 0.453*** | **-0.923\*\*\*** | -0.015 | 0.025*** | -0.008 |
|  | (0.006) | (0.018) | (0.009) | (0.019) | (0.015) | (0.023) | **(0.015)** | (0.011) | (0.008) | (0.006) |
| Sweets | 0.009 | 0.152*** | -0.031** | -0.079*** | -0.057*** | -0.079** | -0.014 | **-0.428\*\*\*** | -0.019 | -0.054*** |
|  | (0.009) | (0.026) | (0.012) | (0.023) | (0.021) | (0.032) | (0.014) | **(0.022)** | (0.012) | (0.009) |
| SSBs | 0.105*** | 0.344*** | -0.047* | 0.062 | -0.048 | -0.136 | 0.079*** | -0.075** | **-1.214\*\*\*** | 0.120*** |
|  | (0.028) | (0.102) | (0.024) | (0.055) | (0.054) | (0.086) | (0.030) | (0.035) | **(0.025)** | (0.026) |
| Non-SSBs | -0.012** | 0.166*** | 0.021*** | 0.119*** | 0.005 | 0.049*** | 0.004 | -0.037*** | 0.040*** | **-1.456\*\*\*** |
|  | (0.006) | (0.016) | (0.004) | (0.010) | (0.009) | (0.014) | (0.006) | (0.007) | (0.007) | **(0.010)** |

Notes: Estimated based on NSS Household Consumption Expenditure survey 2022-23 [1] and using Deaton (1988)'s quality-adjusted Almost Ideal Demand System model [4]. Income groups are defined by terciles of monthly per capita expenditure. Own-price elasticities and their standard errors are denoted in bold. Bootstrapped standard errors after 300 replications. Pack.: Packaged; proc.: processed; F&V: fruits & vegetables; SSBs: sugar-sweetened beverages. \* p<0.1, \*\* p<0.05, \*\*\* p<0.01.

**Table A8.**
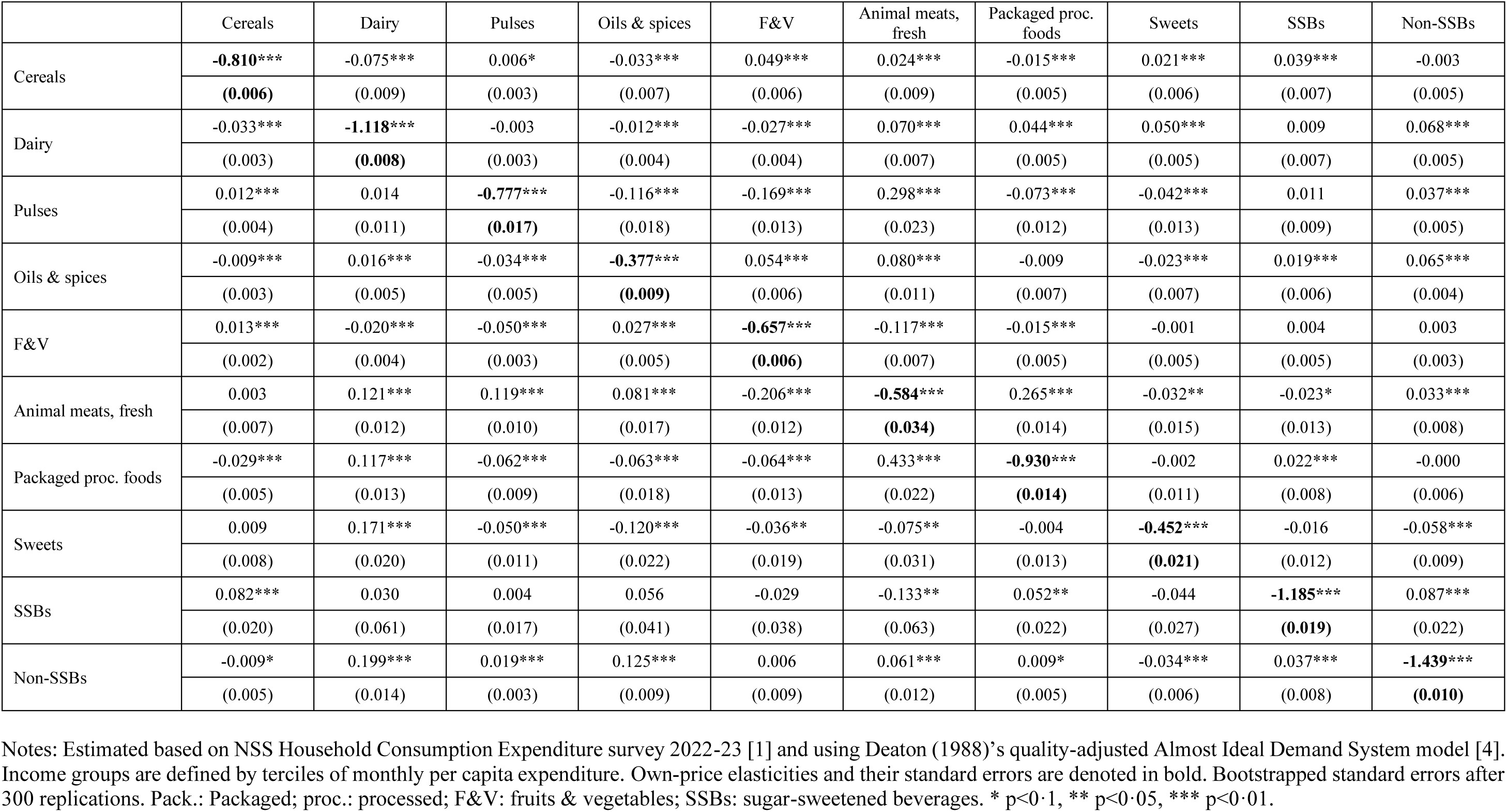
Estimated average price elasticities, middle-income group.

|  | Cereals | Dairy | Pulses | Oils & spices | F&V | Animal meats, fresh | Packaged proc. foods | Sweets | SSBs | Non-SSBs |
| --- | --- | --- | --- | --- | --- | --- | --- | --- | --- | --- |
| Cereals | **-0.810\*\*\*** | -0.075*** | 0.006* | -0.033*** | 0.049*** | 0.024*** | -0.015*** | 0.021*** | 0.039*** | -0.003 |
|  | **(0.006)** | (0.009) | (0.003) | (0.007) | (0.006) | (0.009) | (0.005) | (0.006) | (0.007) | (0.005) |
| Dairy | -0.033*** | **-1.118\*\*\*** | -0.003 | -0.012*** | -0.027*** | 0.070*** | 0.044*** | 0.050*** | 0.009 | 0.068*** |
|  | (0.003) | **(0.008)** | (0.003) | (0.004) | (0.004) | (0.007) | (0.005) | (0.005) | (0.007) | (0.005) |
| Pulses | 0.012*** | 0.014 | **-0.777\*\*\*** | -0.116*** | -0.169*** | 0.298*** | -0.073*** | -0.042*** | 0.011 | 0.037*** |
|  | (0.004) | (0.011) | **(0.017)** | (0.018) | (0.013) | (0.023) | (0.012) | (0.013) | (0.009) | (0.005) |
| Oils & spices | -0.009*** | 0.016*** | -0.034*** | **-0.377\*\*\*** | 0.054*** | 0.080*** | -0.009 | -0.023*** | 0.019*** | 0.065*** |
|  | (0.003) | (0.005) | (0.005) | **(0.009)** | (0.006) | (0.011) | (0.007) | (0.007) | (0.006) | (0.004) |
| F&V | 0.013*** | -0.020*** | -0.050*** | 0.027*** | **-0.657\*\*\*** | -0.117*** | -0.015*** | -0.001 | 0.004 | 0.003 |
|  | (0.002) | (0.004) | (0.003) | (0.005) | **(0.006)** | (0.007) | (0.005) | (0.005) | (0.005) | (0.003) |
| Animal meats, fresh | 0.003 | 0.121*** | 0.119*** | 0.081*** | -0.206*** | **-0.584\*\*\*** | 0.265*** | -0.032** | -0.023* | 0.033*** |
|  | (0.007) | (0.012) | (0.010) | (0.017) | (0.012) | **(0.034)** | (0.014) | (0.015) | (0.013) | (0.008) |
| Packaged proc. foods | -0.029*** | 0.117*** | -0.062*** | -0.063*** | -0.064*** | 0.433*** | **-0.930\*\*\*** | -0.002 | 0.022*** | -0.000 |
|  | (0.005) | (0.013) | (0.009) | (0.018) | (0.013) | (0.022) | **(0.014)** | (0.011) | (0.008) | (0.006) |
| Sweets | 0.009 | 0.171*** | -0.050*** | -0.120*** | -0.036** | -0.075** | -0.004 | **-0.452\*\*\*** | -0.016 | -0.058*** |
|  | (0.008) | (0.020) | (0.011) | (0.022) | (0.019) | (0.031) | (0.013) | **(0.021)** | (0.012) | (0.009) |
| SSBs | 0.082*** | 0.030 | 0.004 | 0.056 | -0.029 | -0.133** | 0.052** | -0.044 | **-1.185\*\*\*** | 0.087*** |
|  | (0.020) | (0.061) | (0.017) | (0.041) | (0.038) | (0.063) | (0.022) | (0.027) | **(0.019)** | (0.022) |
| Non-SSBs | -0.009* | 0.199*** | 0.019*** | 0.125*** | 0.006 | 0.061*** | 0.009* | -0.034*** | 0.037*** | **-1.439\*\*\*** |
|  | (0.005) | (0.014) | (0.003) | (0.009) | (0.009) | (0.012) | (0.005) | (0.006) | (0.008) | **(0.010)** |

Notes: Estimated based on NSS Household Consumption Expenditure survey 2022-23 [1] and using Deaton (1988)'s quality-adjusted Almost Ideal Demand System model [4]. Income groups are defined by terciles of monthly per capita expenditure. Own-price elasticities and their standard errors are denoted in bold. Bootstrapped standard errors after 300 replications. Pack.: Packaged; proc.: processed; F&V: fruits & vegetables; SSBs: sugar-sweetened beverages. \* p<0.1, \*\* p<0.05, \*\*\* p<0.01.

**Table A9.** Estimated average price elasticities, high-income group.

|  | Cereals | Dairy | Pulses | Oils & spices | F&V | Animal meats, fresh | Packaged proc. foods | Sweets | SSBs | Non-SSBs |
| --- | --- | --- | --- | --- | --- | --- | --- | --- | --- | --- |
| Cereals | **-0.772\*\*\*** | -0.140*** | 0.013** | -0.068*** | 0.079*** | 0.024 | -0.034*** | 0.032*** | 0.066*** | -0.003 |
|  | **(0.008)** | (0.015) | (0.006) | (0.012) | (0.010) | (0.019) | (0.009) | (0.010) | (0.013) | (0.009) |
| Dairy | -0.030*** | **-1.190\*\*\*** | 0.005 | 0.028*** | -0.024*** | 0.083*** | 0.062*** | 0.075*** | -0.002 | 0.103*** |
|  | (0.003) | **(0.008)** | (0.003) | (0.005) | (0.005) | (0.010) | (0.006) | (0.007) | (0.010) | (0.007) |
| Pulses | 0.013** | 0.035** | **-0.673\*\*\*** | -0.350*** | -0.246*** | 0.507*** | -0.077*** | -0.055*** | 0.032** | 0.053*** |
|  | (0.006) | (0.017) | **(0.025)** | (0.029) | (0.023) | (0.042) | (0.021) | (0.021) | (0.016) | (0.008) |
| Oils & spices | -0.018*** | 0.058*** | -0.108*** | **-0.139\*\*\*** | 0.089*** | 0.170*** | 0.003 | -0.031*** | 0.048*** | 0.108*** |
|  | (0.004) | (0.008) | (0.009) | **(0.016)** | (0.009) | (0.019) | (0.011) | (0.011) | (0.010) | (0.006) |
| F&V | 0.014*** | -0.027*** | -0.060*** | 0.052*** | **-0.609\*\*\*** | -0.150*** | -0.019*** | 0.004 | -0.005 | 0.010** |
|  | (0.002) | (0.006) | (0.005) | (0.006) | **(0.007)** | (0.010) | (0.007) | (0.007) | (0.007) | (0.005) |
| Animal meats, fresh | 0.004 | 0.155*** | 0.193*** | 0.196*** | -0.255*** | **-0.476\*\*\*** | 0.351*** | -0.032 | -0.026 | 0.052*** |
|  | (0.008) | (0.018) | (0.016) | (0.024) | (0.017) | **(0.059)** | (0.021) | (0.023) | (0.023) | (0.014) |
| Packaged proc. foods | -0.023*** | 0.151*** | -0.046*** | -0.017 | -0.056*** | 0.472*** | **-0.926\*\*\*** | 0.017 | 0.023** | 0.007 |
|  | (0.005) | (0.015) | (0.011) | (0.019) | (0.015) | (0.027) | **(0.017)** | (0.012) | (0.011) | (0.009) |
| Sweets | 0.010 | 0.212*** | -0.042*** | -0.095*** | -0.011 | -0.062* | 0.016 | **-0.461\*\*\*** | -0.023* | -0.060*** |
|  | (0.006) | (0.020) | (0.013) | (0.023) | (0.019) | (0.037) | (0.014) | **(0.022)** | (0.013) | (0.010) |
| SSBs | 0.053*** | -0.035 | 0.025 | 0.125*** | -0.053 | -0.083 | 0.038* | -0.041* | **-1.181\*\*\*** | 0.079*** |
|  | (0.013) | (0.050) | (0.016) | (0.033) | (0.033) | (0.061) | (0.022) | (0.021) | **(0.015)** | (0.023) |
| Non-SSBs | -0.005 | 0.221*** | 0.020*** | 0.138*** | 0.018** | 0.060*** | 0.011 | -0.034*** | 0.040*** | **-1.493\*\*\*** |
|  | (0.004) | (0.015) | (0.004) | (0.008) | (0.009) | (0.016) | (0.007) | (0.007) | (0.010) | **(0.015)** |

Notes: Estimated based on NSS Household Consumption Expenditure survey 2022-23 [1] and using Deaton (1988)'s quality-adjusted Almost Ideal Demand System model [4]. Income groups are defined by terciles of monthly per capita expenditure. Own-price elasticities and their standard errors are denoted in bold. Bootstrapped standard errors after 300 replications. Pack.: Packaged; proc.: processed; F&V: fruits & vegetables; SSBs: sugar-sweetened beverages. \* p<0.1, \*\* p<0.05, \*\*\* p<0.01.

**Figure A4.**
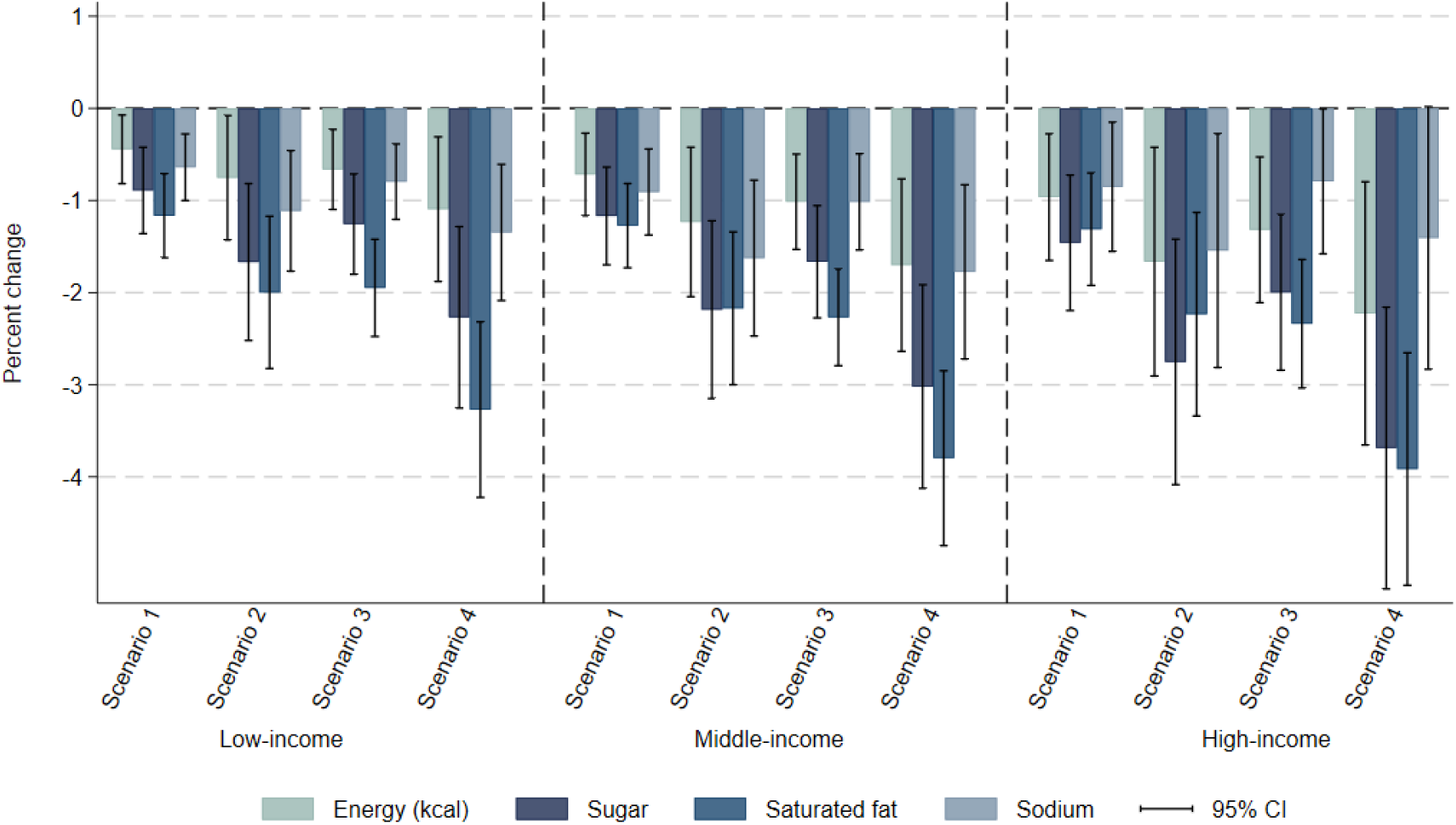
Immediate impact of fiscal policy scenarios on average daily energy and nutrient intake, by income group. Notes: Vertical segments represent the 95% confidence intervals. Survey weighted. Scenario 1: defining items for which GST rate is increased to 28% based on the definition of foods and beverages high in fat, sugar, and sodium by the Food Safety and Standards Authority of India [2]; Scenario 2: adding a 12% top-up to the tax rate applied on HFSS foods and beverages in Scenario 1. Scenario 3: scenario defining items for which GST rate is increased to 28% if their nutrient content is above at least one of the respective thresholds set by the World Health Organization South East Asia Region nutrient profile model [3]; Scenarios 4: adding a 12% top-up to the tax rate applied on HFSS foods and beverages in Scenario 3. GST: Goods and Services Tax. HFSS: High in fat, sodium, and sugar.

**Figure A5.**
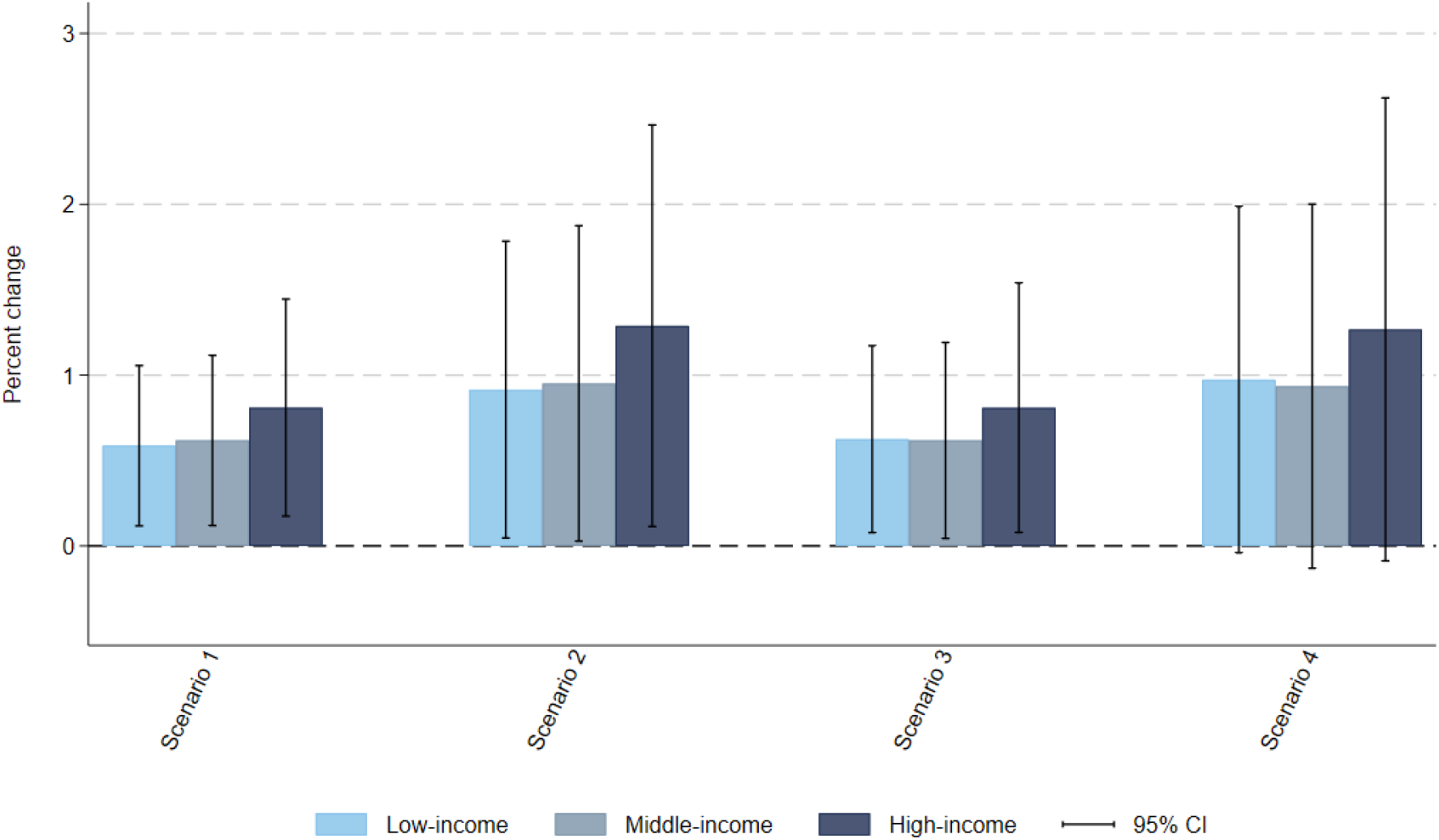
Immediate impact of fiscal policy scenarios on average total household expenditure on food and beverages, by income group. Notes: Vertical segments represent the 95% confidence intervals. Survey weighted. Scenario 1: defining items for which GST rate is increased to 28% based on the definition of foods and beverages high in fat, sugar, and sodium by the Food Safety and Standards Authority of India [2]; Scenario 2: adding a 12% top-up to the tax rate applied on HFSS foods and beverages in Scenario 1. Scenario 3: scenario defining items for which GST rate is increased to 28% if their nutrient content is above at least one of the respective thresholds set by the World Health Organization South East Asia Region nutrient profile model [3]; Scenarios 4: adding a 12% top-up to the tax rate applied on HFSS foods and beverages in Scenario 3. GST: Goods and Services Tax. HFSS: High in fat, sodium, and sugar.

**Figure A6.**
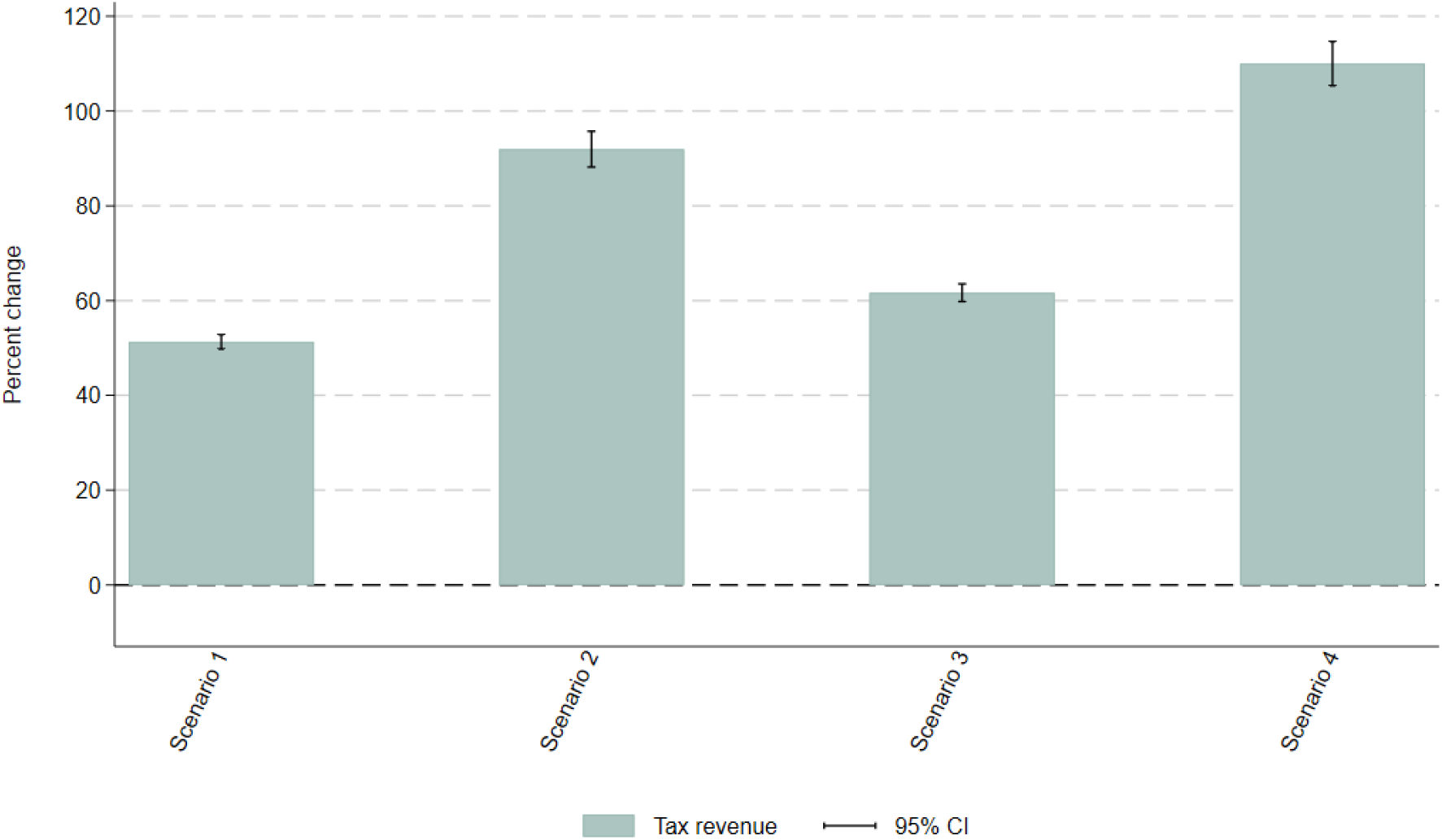
Impact of fiscal policy scenarios on government tax revenue from foods and beverages. Notes: Vertical segments represent the 95% confidence intervals. Survey weighted. Scenario 1: defining items for which GST rate is increased to 28% based on the definition of foods and beverages high in fat, sugar, and sodium by the Food Safety and Standards Authority of India [2]; Scenario 2: adding a 12% top-up to the tax rate applied on HFSS foods and beverages in Scenario 1. Scenario 3: scenario defining items for which GST rate is increased to 28% if their nutrient content is above at least one of the respective thresholds set by the World Health Organization South East Asia Region nutrient profile model [3]; Scenarios 4: adding a 12% top-up to the tax rate applied on HFSS foods and beverages in Scenario 3. GST: Goods and Services Tax. HFSS: High in fat, sodium, and sugar.

**Figure A7.**
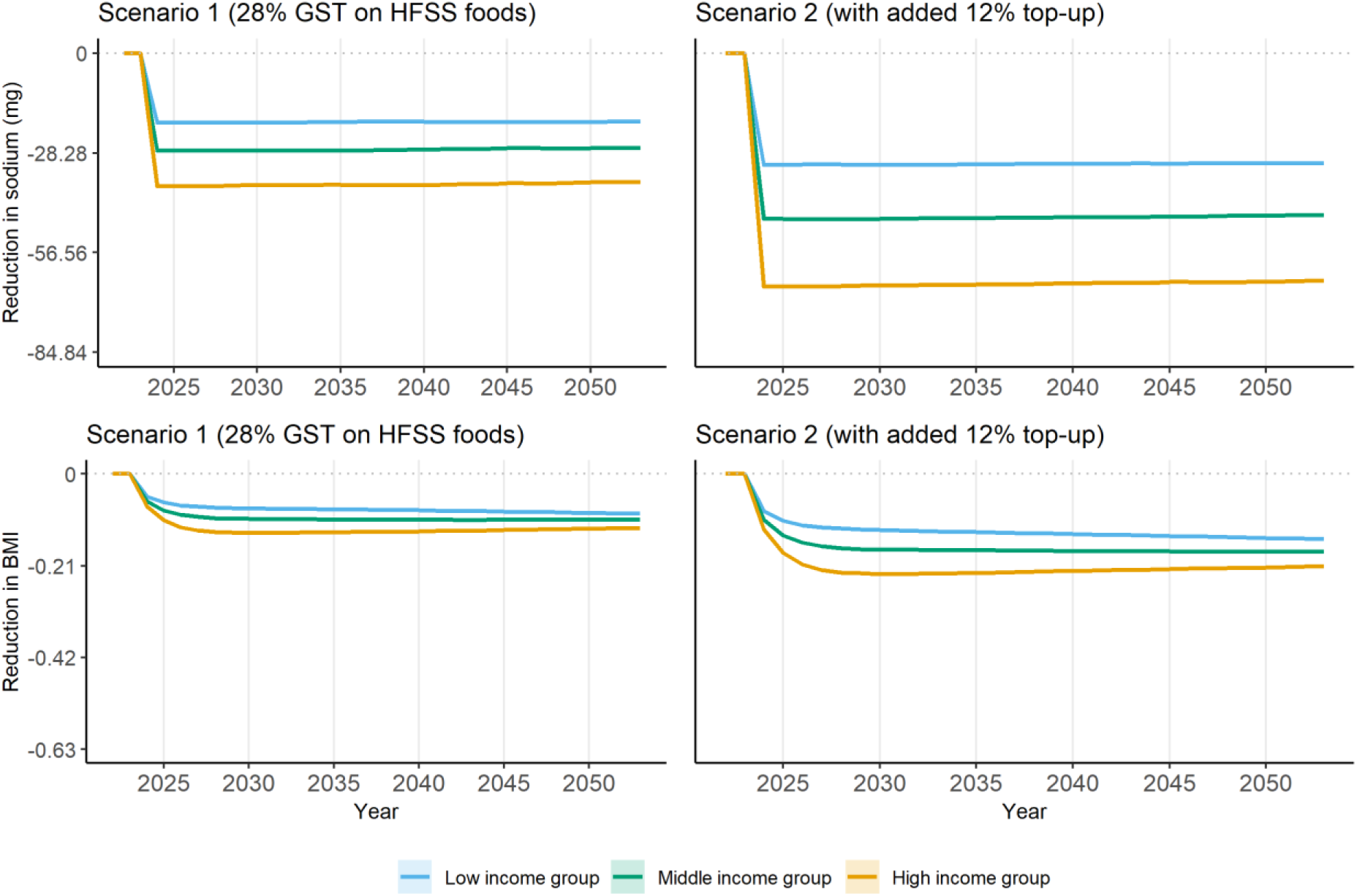
Lower bound of reduction in sodium and body mass index compared to no policy change, by income group, scenarios 1 and 2. Notes: Lower bound estimates (upper bound estimates can be found in **Figure 3**). 95% confidence interval reported as shaded area. In the top two plots on sodium, a unit of 28.28 in y-axis corresponds to 1% of adjusted baseline sodium intake (2,828mg after adjustment according to height and weight distributions). In the bottom two plots on BMI, a unit of 0.21 in y-axis corresponds to 1% of baseline average BMI (21.43kg/m^2^). Policy is introduced in 2024. Scenario 1: defining items for which GST rate is increased to 28% based on the definition of foods and beverages high in fat, sugar, and sodium by the Food Safety and Standards Authority of India [2]; Scenario 2: adding a 12% top-up to the tax rate applied on HFSS foods and beverages in Scenario 1. BMI: body mass index; GST: Goods and Services Tax. HFSS: High in fat, sodium, and sugar.

**Figure A8.**
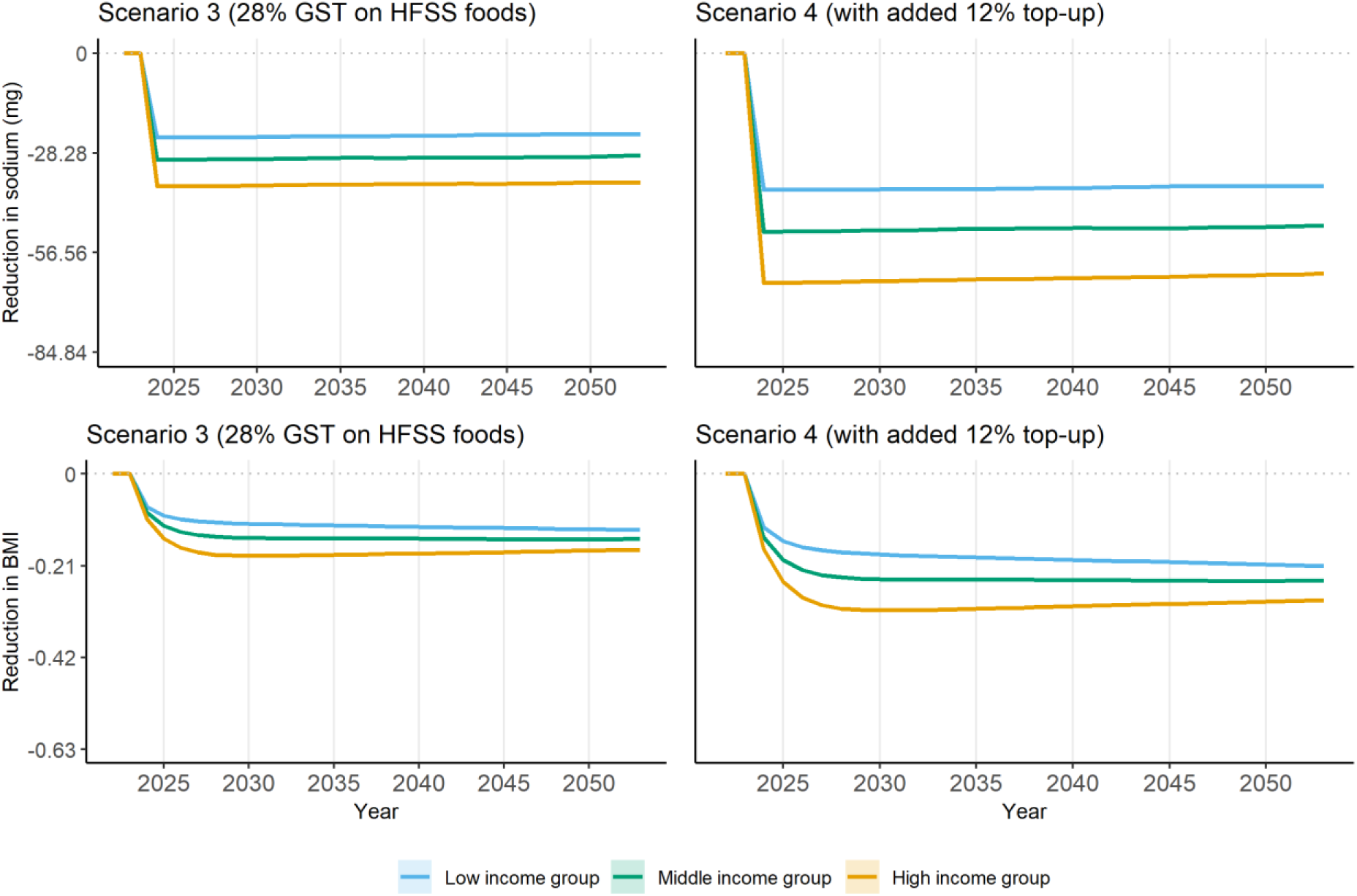
Upper bound of reduction in sodium and body mass index compared to no policy change, by income group, scenarios 3 and 4. Notes: Upper bound estimates (lower bound estimates can be found in **Figure A9**). 95% confidence interval reported as shaded area. In the top two plots on sodium, a unit of 28.28 in y-axis corresponds to 1% of adjusted baseline sodium intake (2,828mg after adjustment according to height and weight distributions). In the bottom two plots on BMI, a unit of 0.21 in y-axis corresponds to 1% of baseline average BMI (21.43kg/m^2^). Policy is introduced in 2024. Scenario 3: scenario defining items for which GST rate is increased to 28% if their nutrient content is above at least one of the respective thresholds set by the World Health Organization South East Asia Region nutrient profile model [3]. Scenarios 4: adding a 12% top-up to the tax rate applied on HFSS foods and beverages in Scenario 3. BMI: body mass index; GST: Goods and Services Tax. HFSS: High in fat, sodium, and sugar.

**Figure A9.**
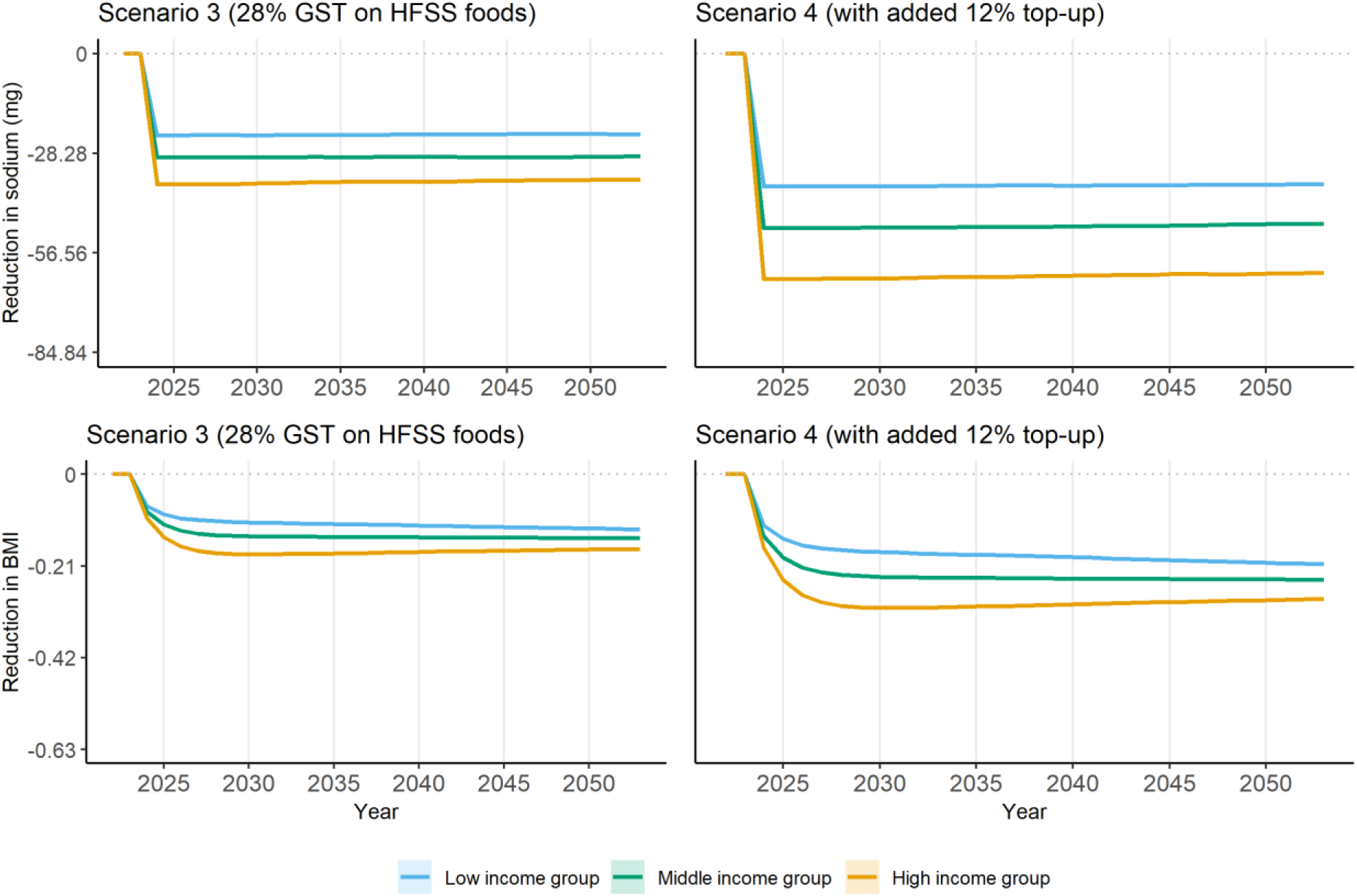
Lower bound of reduction in sodium and body mass index compared to no policy change, by income group, scenarios 3 and 4. Notes: Lower bound estimates (upper bound estimates can be found in **Figure A8**). 95% confidence interval reported as shaded area. In the top two plots on sodium, a unit of 28.28 in y-axis corresponds to 1% of adjusted baseline sodium intake (2,828mg after adjustment according to height and weight distributions). In the bottom two plots on BMI, a unit of 0.21 in y-axis corresponds to 1% of baseline average BMI (21.43kg/m^2^). Policy is introduced in 2024. Scenario 3: scenario defining items for which GST rate is increased to 28% if their nutrient content is above at least one of the respective thresholds set by the World Health Organization South East Asia Region nutrient profile model [3]. Scenarios 4: adding a 12% top-up to the tax rate applied on HFSS foods and beverages in Scenario 3. BMI: body mass index; GST: Goods and Services Tax. HFSS: High in fat, sodium, and sugar.

**Table A10.** Upper bound cumulative reduction in the incidence of five key diseases in the entire population and associated change in total health expenditure and DALYs over 30 years of policy implementation, under four policy scenarios.

| Disease | Annual incidence at baseline*<br>(million cases) | Scenario 1 |  | Scenario 2 |  | Scenario 3 |  | Scenario 4 |  |
| --- | --- | --- | --- | --- | --- | --- | --- | --- | --- |
|  |  | CR<sup>a</sup><br>(million cases, %) | THE<sup>b</sup><br>(USD billion) | CR<sup>a</sup><br>(million cases, %) | THE<sup>b</sup><br>(USD billion) | CR<sup>a</sup><br>(million cases, %) | THE<sup>b</sup><br>(USD billion) | CR<sup>a</sup><br>(million cases, %) | THE<sup>b</sup><br>(USD billion) |
| IHD | 6.45 | -1.85 (-0.96) | -4.53 | -3.06 (-1.58) | -7.50 | -2.44 (-1.26) | -5.98 | -4.00 (-2.07) | -9.83 |
| CKD | 2.10 | -0.34 (-0.54) | -0.92 | -0.59 (-0.93) | -1.59 | -0.45 (-0.72) | -1.23 | -0.77 (-1.22) | -2.07 |
| Stroke | 1.20 | -0.22 (-0.61) | -0.69 | -0.34 (-0.94) | -1.08 | -0.25 (-0.71) | -0.81 | -0.41 (-1.15) | -1.32 |
| Diabetes | 3.40 | -1.80 (-1.76) | -2.79 | -3.05 (-2.99) | -4.73 | -2.52 (-2.47) | -3.90 | -4.07 (-3.99) | -6.31 |
| Asthma | 4.77 | -1.24 (-0.87) | -1.77 | -2.20 (-1.53) | -3.13 | -1.84 (-1.28) | -2.62 | -2.95 (-2.06) | -4.21 |
| **Total** | **17.92** | **-5.44 (-1.01)** | **-10.70** | **-9.23 (-1.72)** | **-18.03** | **-7.50 (-1.40)** | **-14.55** | **-12.20 (-2.27)** | **-23.74** |
| **Cumulative reduction in DALYs (million)** |  | **-10.36** |  | **-18.98** |  | **-15.55** |  | **-27.37** |  |

Notes: Upper bound estimates (lower bound estimates can be found in **Table A11**). Results estimated over a 30-year period (2024-2053). <sup>a</sup>CR: Cumulative reduction over 30 years, in million cases. Percentage of annual baseline incidence number (2019) in India per year in parenthesis, calculated as cumulative reduction over 30 years ÷ (annual incidence × 30). <sup>b</sup>THE: Total health expenditure over 30 years, in billion US dollar 2024. Scenario 1: defining items for which GST rate is increased to 28% based on the definition of foods and beverages high in fat, sugar, and sodium by the Food Safety and Standards Authority of India [2]; Scenario 2: adding a 12% top-up to the tax rate applied on HFSS foods and beverages in Scenario 1. Scenario 3: defining items for which GST rate is increased to 28% if their nutrient content is above at least one of the respective thresholds set by the World Health Organization South East Asia Region nutrient profile model [3]; Scenarios 4: adding a 12% top-up to the tax rate applied on HFSS foods and beverages in Scenario 3. IHD: Ischemic heart disease. CKD: Chronic kidney disease. GST: Goods and Services Tax. HFSS: High in fat, sodium, and sugar. \*Source: Institute for Health Metrics and Evaluation (IHME), Global Burden of Disease Study 2019 (GBD 2019) results [5].

**Table A11.** Lower bound cumulative reduction in the incidence of five key diseases in the entire population and associated change in total health expenditure and DALYs over 30 years of policy implementation, under four policy scenarios.

| Disease | Annual incidence at baseline*<br>(million cases) | Scenario 1 |  | Scenario 2 |  | Scenario 3 |  | Scenario 4 |  |
| --- | --- | --- | --- | --- | --- | --- | --- | --- | --- |
|  |  | CR<sup>a</sup><br>(million cases, %) | THE<sup>b</sup><br>(USD billion) | CR<sup>a</sup><br>(million cases, %) | THE<sup>b</sup><br>(USD billion) | CR<sup>a</sup><br>(million cases, %) | THE<sup>b</sup><br>(USD billion) | CR<sup>a</sup><br>(million cases, %) | THE<sup>b</sup><br>(USD billion) |
| IHD | 6.45 | -1.78 (-0.92) | -4.38 | -3.06 (-1.58) | -7.50 | -2.44 (-1.26) | -5.98 | -3.94 (-2.03) | -9.66 |
| CKD | 2.10 | -0.37 (-0.58) | -0.99 | -0.63 (-1.00) | -1.71 | -0.48 (-0.76) | -1.29 | -0.81 (-1.28) | -2.18 |
| Stroke | 1.20 | -0.21 (-0.57) | -0.66 | -0.30 (-0.84) | -0.97 | -0.26 (-0.71) | -0.82 | -0.40 (-1.11) | -1.27 |
| Diabetes | 3.40 | -1.77 (-1.74) | -2.75 | -3.02 (-2.96) | -4.68 | -2.50 (-2.45) | -3.87 | -4.01 (-3.93) | -6.21 |
| Asthma | 4.77 | -1.25 (-0.88) | -1.79 | -2.05 (-1.44) | -2.93 | -1.78 (-1.24) | -2.54 | -2.87 (-2.01) | -4.10 |
| **Total** | **17.92** | **-5.39 (-1.00)** | **-10.57** | **-9.06 (-1.69)** | **-17.78** | **-7.45 (-1.39)** | **-14.50** | **-12.02 (-2.24)** | **-23.42** |
| **Cumulative reduction in DALYs (million)** |  | **-12.26** |  | **-19.97** |  | **-17.68** |  | **-24.82** |  |

Notes: Lower bound estimates (upper bound estimates can be found in **Table A10**). Results estimated over a 30-year period (2024-2053). <sup>a</sup>CR: Cumulative reduction over 30 years, in million cases. Percentage of annual baseline incidence number (2019) in India per year in parenthesis, calculated as cumulative reduction over 30 years ÷ (annual incidence × 30). <sup>b</sup>THE: Total health expenditure over 30 years, in billion US dollar 2024. Scenario 1: defining items for which GST rate is increased to 28% based on the definition of foods and beverages high in fat, sugar, and sodium by the Food Safety and Standards Authority of India [2]; Scenario 2: adding a 12% top-up to the tax rate applied on HFSS foods and beverages in Scenario 1. Scenario 3: defining items for which GST rate is increased to 28% if their nutrient content is above at least one of the respective thresholds set by the World Health Organization South East Asia Region nutrient profile model [3]; Scenarios 4: adding a 12% top-up to the tax rate applied on HFSS foods and beverages in Scenario 3. IHD: Ischemic heart disease. CKD: Chronic kidney disease. GST: Goods and Services Tax. HFSS: High in fat, sodium, and sugar. \*Source: Institute for Health Metrics and Evaluation (IHME), Global Burden of Disease Study 2019 (GBD 2019) results [5].

**Table A12.** Lower bound of cumulative reduction in disease incidence rate (cases per 100,000 population) compared to no policy change, by income group, scenarios 1 and 2.

| Disease | Scenario 1 |  |  | Scenario 2 |  |  |
| --- | --- | --- | --- | --- | --- | --- |
|  | Low income | Middle income | High income | Low income | Middle income | High income |
| IHD | -68 | -110 | -191 | -115 | -188 | -332 |
| CKD | -13 | -27 | -36 | -25 | -45 | -60 |
| Stroke | -9 | -12 | -22 | -14 | -18 | -32 |
| Diabetes | -72 | -119 | -171 | -119 | -197 | -305 |
| Asthma | -67 | -79 | -105 | -101 | -130 | -184 |
| **Total** | **-230** | **-347** | **-525** | **-374** | **-577** | **-913** |

Notes: Lower bound estimates (upper bound estimates can be found in **Table 3**). Results estimated over a 30-year period (2024-2053). Scenario 1: GST rate is increased to 28% for foods and beverages high in fat, sugar, and sodium based on the definition by the Food Safety and Standards Authority of India [2]; Scenario 2: adding a 12% top-up to the tax rate applied on HFSS foods and beverages in Scenario 1. IHD: Ischemic heart disease. CKD: Chronic kidney disease. GST: Goods and Services Tax. HFSS: High in fat, sugar, and sodium.

**Table A13.** Upper bound of cumulative reduction in disease incidence rate (cases per 100,000 population) compared to no policy change, by income group, scenarios 3 and 4.

| Disease | Scenario 3 |  |  | Scenario 4 |  |  |
| --- | --- | --- | --- | --- | --- | --- |
|  | Low income | Middle income | High income | Low income | Middle income | High income |
| IHD | -96 | -161 | -243 | -154 | -267 | -403 |
| CKD | -17 | -31 | -49 | -31 | -47 | -84 |
| Stroke | -9 | -18 | -27 | -14 | -32 | -41 |
| Diabetes | -117 | -158 | -234 | -179 | -260 | -389 |
| Asthma | -86 | -123 | -164 | -136 | -194 | -270 |
| **Total** | **-325** | **-490** | **-717** | **-513** | **-800** | **-1188** |

Notes: Upper bound estimates (lower bound estimates can be found in **Table A14**). Results estimated over a 30-year period (2024-2053). Scenario 3: scenario defining items for which GST rate is increased to 28% if their nutrient content is above at least one of the respective thresholds set by the World Health Organization South East Asia Region nutrient profile model [3]. Scenarios 4: adding a 12% top-up to the tax rate applied on HFSS foods and beverages in Scenario 3. IHD: Ischemic heart disease. CKD: Chronic kidney disease. GST: Goods and Services Tax. HFSS: High in fat, sugar, and sodium.

**Table A14.** Lower bound of cumulative reduction in disease incidence rate (cases per 100,000 population) compared to no policy change, by income group, scenarios 3 and 4.

| Disease | Scenario 3 |  |  | Scenario 4 |  |  |
| --- | --- | --- | --- | --- | --- | --- |
|  | Low income | Middle income | High income | Low income | Middle income | High income |
| IHD | -100 | -153 | -248 | -148 | -253 | -413 |
| CKD | -24 | -28 | -47 | -33 | -54 | -80 |
| Stroke | -11 | -14 | -29 | -19 | -21 | -44 |
| Diabetes | -108 | -162 | -239 | -167 | -253 | -400 |
| Asthma | -86 | -120 | -151 | -139 | -188 | -253 |
| **Total** | **-329** | **-477** | **-714** | **-506** | **-768** | **-1190** |

Notes: Lower bound estimates (upper bound estimates can be found in **Table A13**). Results estimated over a 30-year period (2024-2053). Scenario 3: scenario defining items for which GST rate is increased to 28% if their nutrient content is above at least one of the respective thresholds set by the World Health Organization South East Asia Region nutrient profile model [3]. Scenarios 4: adding a 12% top-up to the tax rate applied on HFSS foods and beverages in Scenario 3. IHD: Ischemic heart disease. CKD: Chronic kidney disease. GST: Goods and Services Tax. HFSS: High in fat, sugar, and sodium.

**Figure A10.**
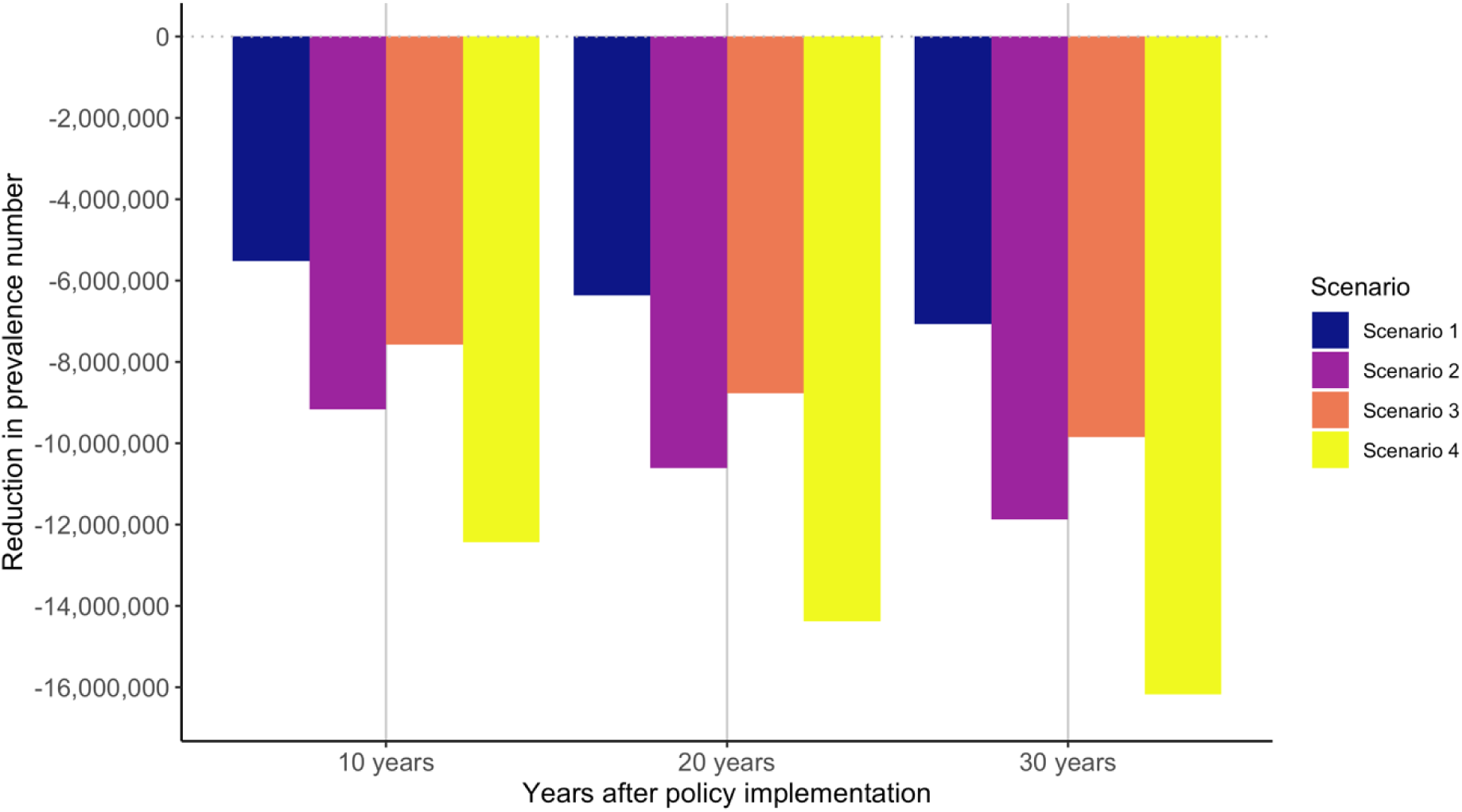
Upper bound reduction in the prevalence number of hypertension in the entire population in 10, 20 and 30 years after policy implementation under four policy scenarios. Notes: Reduction in the prevalence number of hypertension is calculated after microsimulation, with the reduction in sodium intake and BMI from Health-GPS microsimulation and relative risks of sodium and BMI on the incidence of hypertension from the literature. Upper bound estimates (lower bound estimates can be found in **Figure A10**). Scenario 1: defining items for which GST rate is increased to 28% based on the definition of foods and beverages high in fat, sugar, and sodium by the Food Safety and Standards Authority of India [2]; Scenario 2: adding a 12% top-up to the tax rate applied on HFSS foods and beverages in Scenario 1. Scenario 3: defining items for which GST rate is increased to 28% if their nutrient content is above at least one of the respective thresholds set by the World Health Organization South East Asia Region nutrient profile model [3]; Scenarios 4: adding a 12% top-up to the tax rate applied on HFSS foods and beverages in Scenario 3.GST: Goods and Services Tax. HFSS: High in fat, sodium, and sugar.

**Figure A11.**
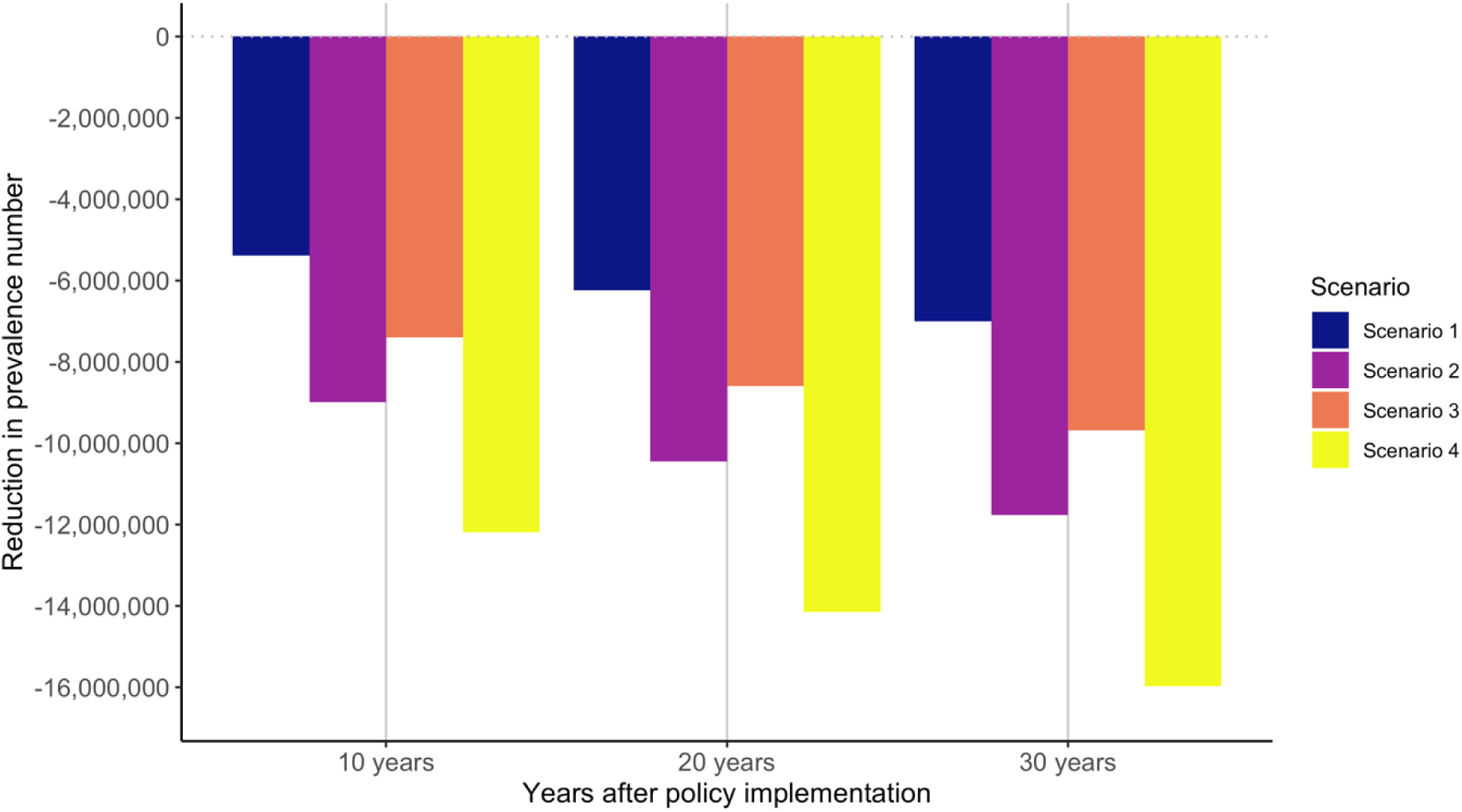
Lower bound reduction in the prevalence number of hypertension in the entire population in 10, 20 and 30 years after policy implementation under four policy scenarios. Notes: Reduction in the prevalence number of hypertension is calculated after microsimulation, with the reduction in sodium intake and BMI from Health-GPS microsimulation and relative risks of sodium and BMI on the incidence of hypertension from the literature. Lower bound estimates (upper bound estimates can be found in **Figure A9**). Scenario 1: defining items for which GST rate is increased to 28% based on the definition of foods and beverages high in fat, sugar, and sodium by the Food Safety and Standards Authority of India [2]; Scenario 2: adding a 12% top-up to the tax rate applied on HFSS foods and beverages in Scenario 1. Scenario 3: defining items for which GST rate is increased to 28% if their nutrient content is above at least one of the respective thresholds set by the World Health Organization South East Asia Region nutrient profile model [3]; Scenarios 4: adding a 12% top-up to the tax rate applied on HFSS foods and beverages in Scenario 3. GST: Goods and Services Tax. HFSS: High in fat, sodium, and sugar.

**Figure A12.**
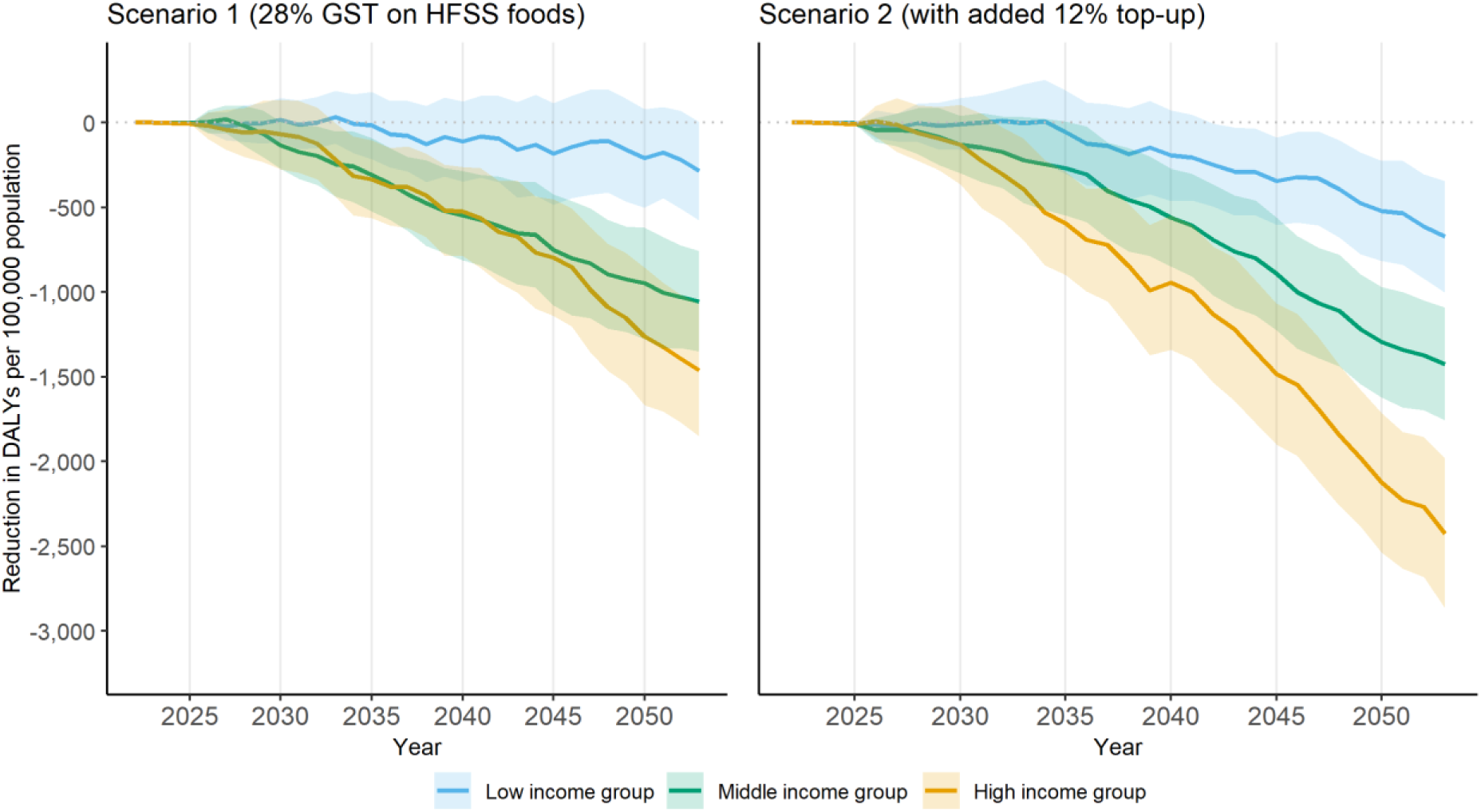
Lower bound of cumulative reduction in DALYs per 100,000 population compared to no policy change, by income group, scenarios 1 and 2. Notes: Cumulative reduction in DALYs per 100,000 population of five key diseases including ischemic heart disease, chronic kidney disease, stroke, diabetes and asthma over 2022-2053. Lower bound estimates (upper bound estimates can be found in **Figure 4**). 95% confidence interval reported as shaded area. Policy is introduced in 2024. Scenario 1: GST rate is increased to 28% for foods and beverages high in fat, sugar, and sodium based on the definition by the Food Safety and Standards Authority of India [2]; Scenario 2: adding a 12% top-up to the tax rate applied on HFSS foods and beverages in Scenario 1. DALYs: Disability-Adjusted Life Years. HFSS: High in fat, sodium, and sugar.

**Figure A13.**
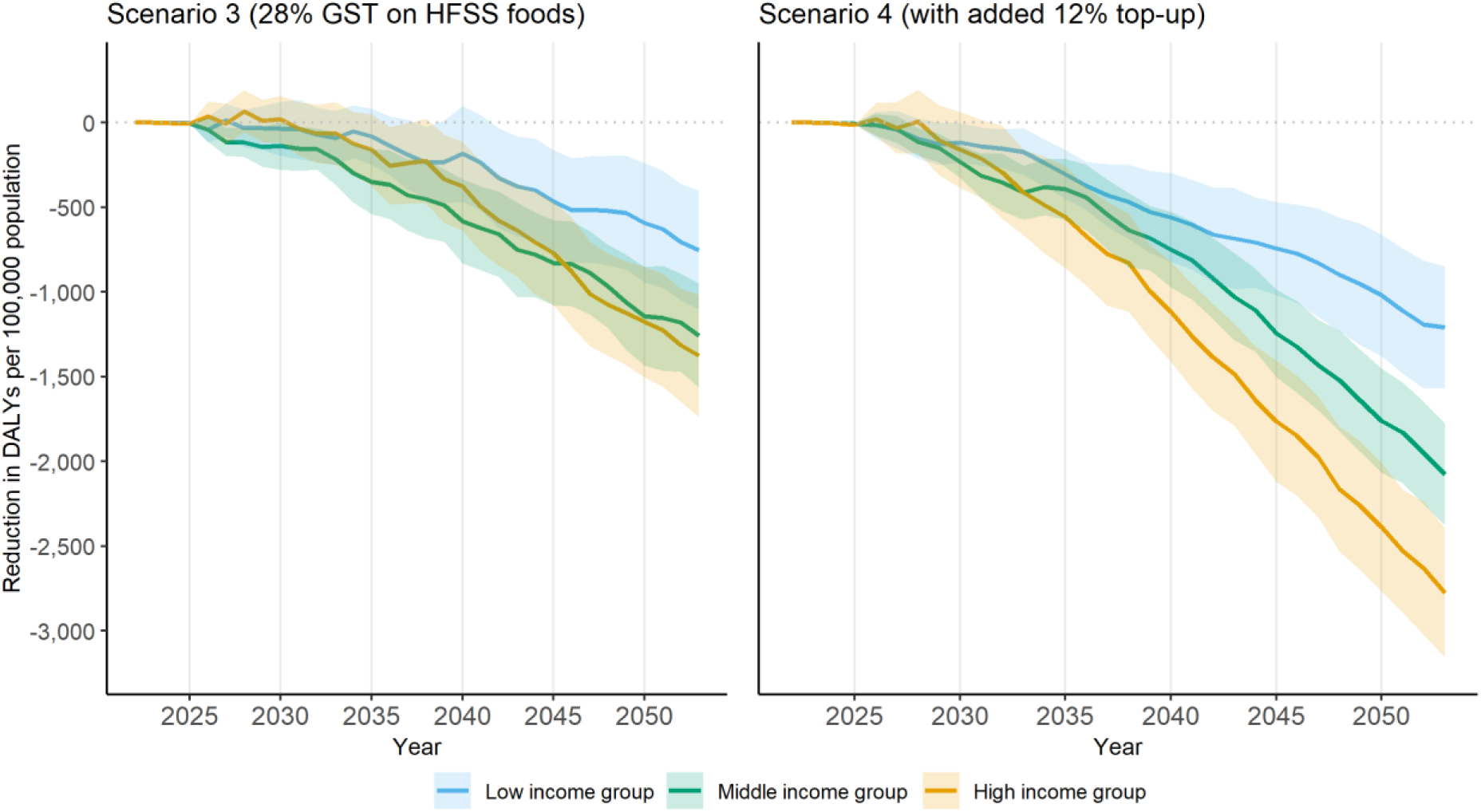
Upper bound of cumulative reduction in DALYs per 100,000 population compared to baseline, scenarios 3 and 4. Notes: Cumulative reduction in DALYs per 100,000 population of five key diseases including ischemic heart disease, chronic kidney disease, stroke, diabetes and asthma over 2022-2053. Upper bound estimates (lower bound estimates can be found in **Figure A14**). 95% confidence interval reported as shaded area. Policy is introduced in 2024. Scenario 3: defining items for which GST rate is increased to 28% if their nutrient content is above at least one of the respective thresholds set by the World Health Organization South East Asia Region nutrient profile model [3]. Scenarios 4: adding a 12% top-up to the tax rate applied on HFSS foods and beverages in Scenario 3. DALYs: Disability-Adjusted Life Years. HFSS: High in fat, sodium, and sugar.

**Figure A14.**
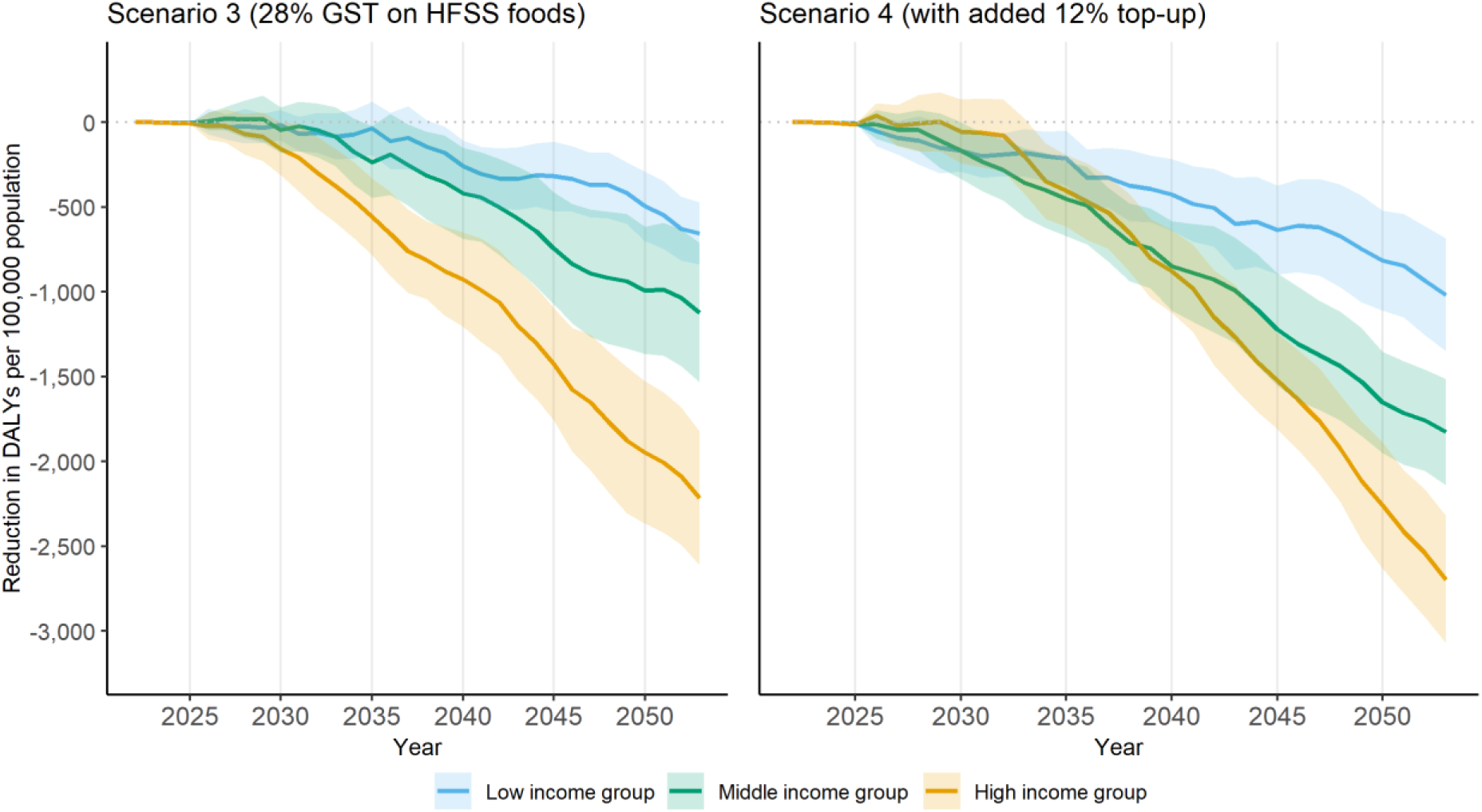
Lower bound of cumulative reduction in DALYs per 100,000 population compared to baseline, scenarios 3 and 4. Notes: Cumulative reduction in DALYs per 100,000 population of five key diseases including ischemic heart disease, chronic kidney disease, stroke, diabetes and asthma over 2022-2053. Lower bound estimates (upper bound estimates can be found in **Figure A13**). 95% confidence interval reported as shaded area. Policy is introduced in 2024. Scenario 3: defining items for which GST rate is increased to 28% if their nutrient content is above at least one of the respective thresholds set by the World Health Organization South East Asia Region nutrient profile model [3]. Scenarios 4: adding a 12% top-up to the tax rate applied on HFSS foods and beverages in Scenario 3. DALYs: Disability-Adjusted Life Years. HFSS: High in fat, sodium, and sugar.

**Table A15.** Summary of lower bound results, scenarios 1 and 2.

|  |  | Immediate effect at implementation |  | Cumulative results for the five key diseases over 2024-2053 |  |  |
| --- | --- | --- | --- | --- | --- | --- |
|  |  | Change in HH expenditure on foods and beverages (%) | Change in tax revenue from foods and beverages (%) | Change in incidence rate per 100,000 population | Change in DALYs per 100,000 population | Change in total health expenditure (USD billion) |
| Scenario 1 | Low income | +0.6% | +51.3% | -230 | -285 | -10.6 |
|  | Middle income | +0.6% |  | -347 | -1053 |  |
|  | High income | +0.8% |  | -525 | -1460 |  |
| Scenario 2 | Low income | +0.9% | +92.0% | -374 | -672 | -17.8 |
|  | Middle income | +1.0% |  | -577 | -1424 |  |
|  | High income | +1.3% |  | -913 | -2424 |  |

Notes: Lower bound estimates (upper bound estimates can be found in **Table 4**). Results estimated over a 30-year period (2024-2053). Scenario 1: GST rate is increased to 28% for foods and beverages high in fat, sugar, and sodium based on the definition by the Food Safety and Standards Authority of India [2]; Scenario 2: adding a 12% top-up to the tax rate applied on HFSS foods and beverages in Scenario 1. DALYs: Disability-adjusted life years. GST: Goods and Services Tax. HFSS: High in fat, sugar, and sodium. USD: US dollar 2024.

**Table A16.** Summary of upper bound results, scenarios 3 and 4.

|  |  | Immediate effect at implementation |  | Cumulative results for the five key diseases over 2024-2053 |  |  |
| --- | --- | --- | --- | --- | --- | --- |
|  |  | Change in HH expenditure on foods and beverages (%) | Change in tax revenue from foods and beverages (%) | Change in incidence rate per 100,000 population | Change in DALYs per 100,000 population | Change in total health expenditure (USD billion) |
| Scenario 3 | Low income | +0.6% | +61.7% | -325 | -752 | -14.6 |
|  | Middle income | +0.6% |  | -490 | -1256 |  |
|  | High income | +0.8% |  | -717 | -1375 |  |
| Scenario 4 | Low income | +1.0% | +110.0% | -513 | -1210 | -23.7 |
|  | Middle income | +0.9% |  | -800 | -2077 |  |
|  | High income | +1.3% |  | -1188 | -2776 |  |

Notes: Upper bound estimates (lower bound estimates can be found in **Table A17**). Results estimated over a 30-year period (2024-2053). Scenario 3: scenario defining items for which GST rate is increased to 28% if their nutrient content is above at least one of the respective thresholds set by the World Health Organization South East Asia Region nutrient profile model [3]. Scenarios 4: adding a 12% top-up to the tax rate applied on HFSS foods and beverages in Scenario 3. DALYs: Disability-adjusted life years. GST: Goods and Services Tax. HFSS: High in fat, sugar, and sodium. USD: US dollar 2024.

**Table A17.** Summary of lower bound results, scenarios 3 and 4.

|  |  | Immediate effect at implementation |  | Cumulative results for the five key diseases over 2024-2053 |  |  |
| --- | --- | --- | --- | --- | --- | --- |
|  |  | Change in HH expenditure on foods and beverages (%) | Change in tax revenue from foods and beverages (%) | Change in incidence rate per 100,000 population | Change in DALYs per 100,000 population | Change in total health expenditure (USD billion) |
| Scenario 3 | Low income | +0.6% | +61.7% | -329 | -654 | -14.5 |
|  | Middle income | +0.6% |  | -477 | -1122 |  |
|  | High income | +0.8% |  | -714 | -2216 |  |
| Scenario 4 | Low income | +1.0% | +110.0% | -506 | -1018 | -23.4 |
|  | Middle income | +0.9% |  | -768 | -1826 |  |
|  | High income | +1.3% |  | -1190 | -2696 |  |

Notes: Lower bound estimates (upper bound estimates can be found in **Table A16**). Results estimated over a 30-year period (2024-2053). Scenario 3: scenario defining items for which GST rate is increased to 28% if their nutrient content is above at least one of the respective thresholds set by the World Health Organization South East Asia Region nutrient profile model [3]. Scenarios 4: adding a 12% top-up to the tax rate applied on HFSS foods and beverages in Scenario 3. DALYs: Disability-adjusted life years. GST: Goods and Services Tax. HFSS: High in fat, sugar, and sodium. USD: US dollar 2024.

### Supporting information file 2

#### Appendix B. Sensitivity scenarios

Appendix B forms part of the revised submission.

Supplement to: Roche M, Zhu J, Olney J, Laydon DJ, Joe W, Sharma M, Steele L, Sassi F. Taxation of foods high in saturated fat, sugar, and sodium in India: A modelling study of health and economic impacts. Submitted after revision on 19 September 2025.

**Table B1.**
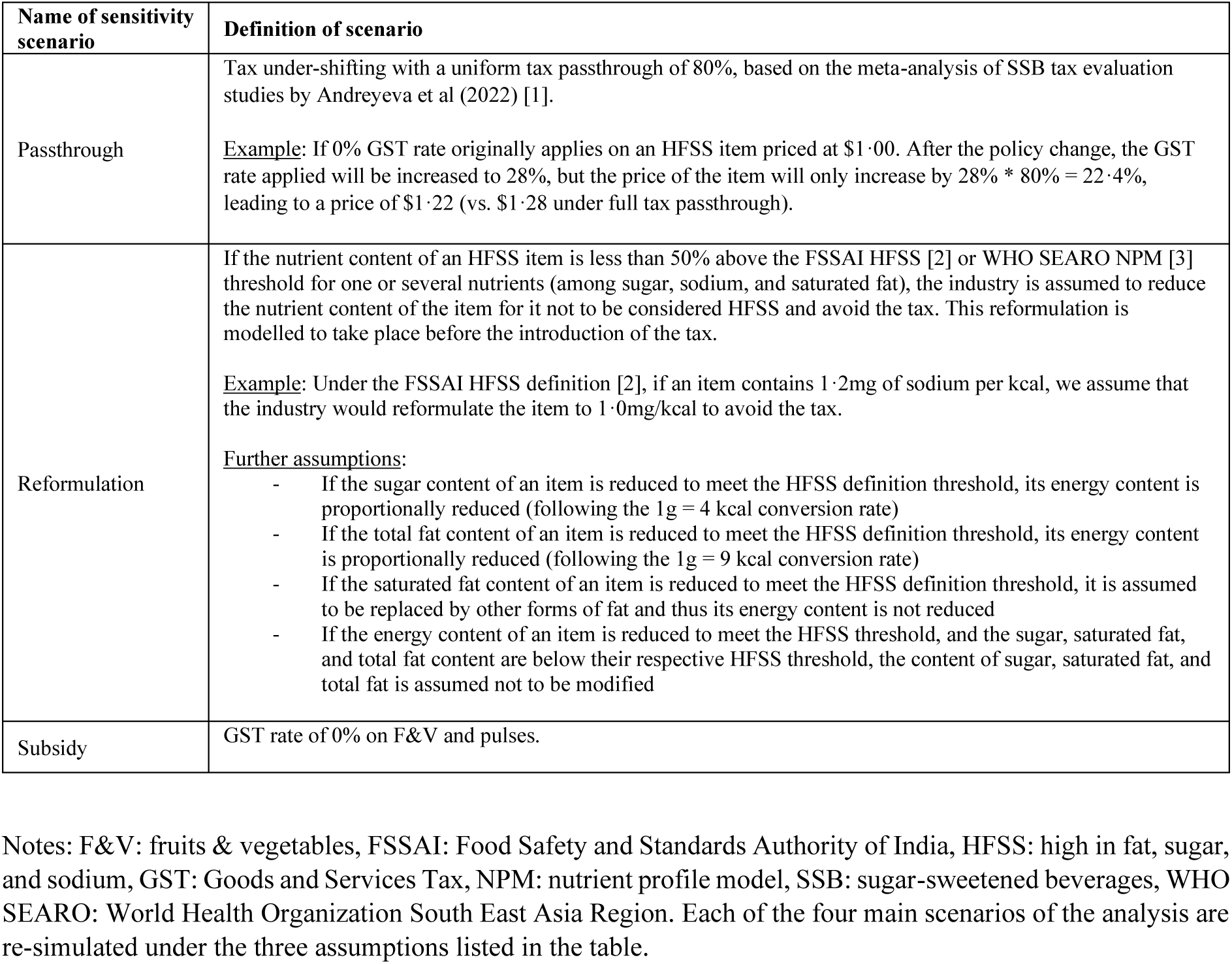
Sensitivity scenarios definition.

| Name of sensitivity scenario | Definition of scenario |
| --- | --- |
| Passthrough | Tax under-shifting with a uniform tax passthrough of 80%, based on the meta-analysis of SSB tax evaluation studies by Andreyeva et al (2022) [1].<br><u>Example:</u> If 0% GST rate originally applies on an HFSS item priced at \$1·00. After the policy change, the GST rate applied will be increased to 28%, but the price of the item will only increase by $28\% \cdot 80\% = 22\cdot4\%$, leading to a price of \$1·22 (vs. \$1·28 under full tax passthrough). |
| Reformulation | If the nutrient content of an HFSS item is less than 50% above the FSSAI HFSS [2] or WHO SEARO NPM [3] threshold for one or several nutrients (among sugar, sodium, and saturated fat), the industry is assumed to reduce the nutrient content of the item for it not to be considered HFSS and avoid the tax. This reformulation is modelled to take place before the introduction of the tax.<br><u>Example:</u> Under the FSSAI HFSS definition [2], if an item contains 1·2mg of sodium per kcal, we assume that the industry would reformulate the item to 1·0mg/kcal to avoid the tax.<br><u>Further assumptions:</u><br>- If the sugar content of an item is reduced to meet the HFSS definition threshold, its energy content is proportionally reduced (following the 1g = 4 kcal conversion rate)<br>- If the total fat content of an item is reduced to meet the HFSS definition threshold, its energy content is proportionally reduced (following the 1g = 9 kcal conversion rate)<br>- If the saturated fat content of an item is reduced to meet the HFSS definition threshold, it is assumed to be replaced by other forms of fat and thus its energy content is not reduced<br>- If the energy content of an item is reduced to meet the HFSS threshold, and the sugar, saturated fat, and total fat content are below their respective HFSS threshold, the content of sugar, saturated fat, and total fat is assumed not to be modified |
| Subsidy | GST rate of 0% on F&V and pulses. |

Notes: F&V: fruits & vegetables, FSSAI: Food Safety and Standards Authority of India, HFSS: high in fat, sugar, and sodium, GST: Goods and Services Tax, NPM: nutrient profile model, SSB: sugar-sweetened beverages, WHO SEARO: World Health Organization South East Asia Region. Each of the four main scenarios of the analysis are re-simulated under the three assumptions listed in the table.

**Figure B1.**
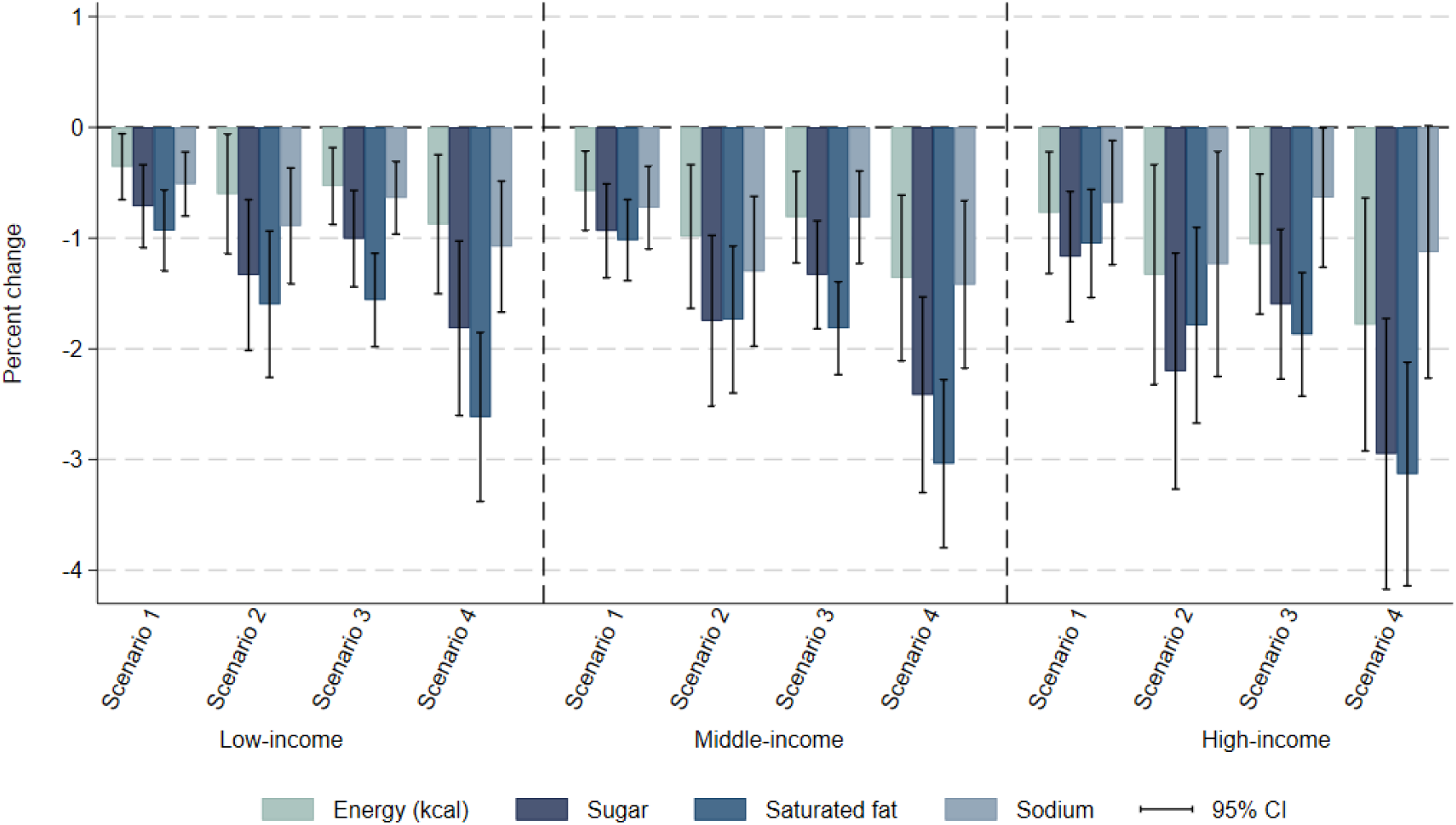
Sensitivity scenario: Passthrough. Immediate impact of fiscal policy scenarios on average daily energy and nutrient intake, by income group. Notes: This is a sensitivity analysis using an incomplete passthrough of tax rate changes to prices of 80%. Vertical segments represent the 95% confidence intervals. Survey weighted. Scenario 1: defining items for which GST rate is increased to 28% based on the definition of foods and beverages high in fat, sugar, and sodium by the Food Safety and Standards Authority of India [2]; Scenario 2: adding a 12% top-up to the tax rate applied on HFSS foods and beverages in Scenario 1. Scenario 3: scenario defining items for which GST rate is increased to 28% if their nutrient content is above at least one of the respective thresholds set by the World Health Organization South East Asia Region nutrient profile model [3]; Scenarios 4: adding a 12% top-up to the tax rate applied on HFSS foods and beverages in Scenario 3. GST: Goods and Services Tax. HFSS: High in fat, sodium, and sugar.

**Figure B2.**
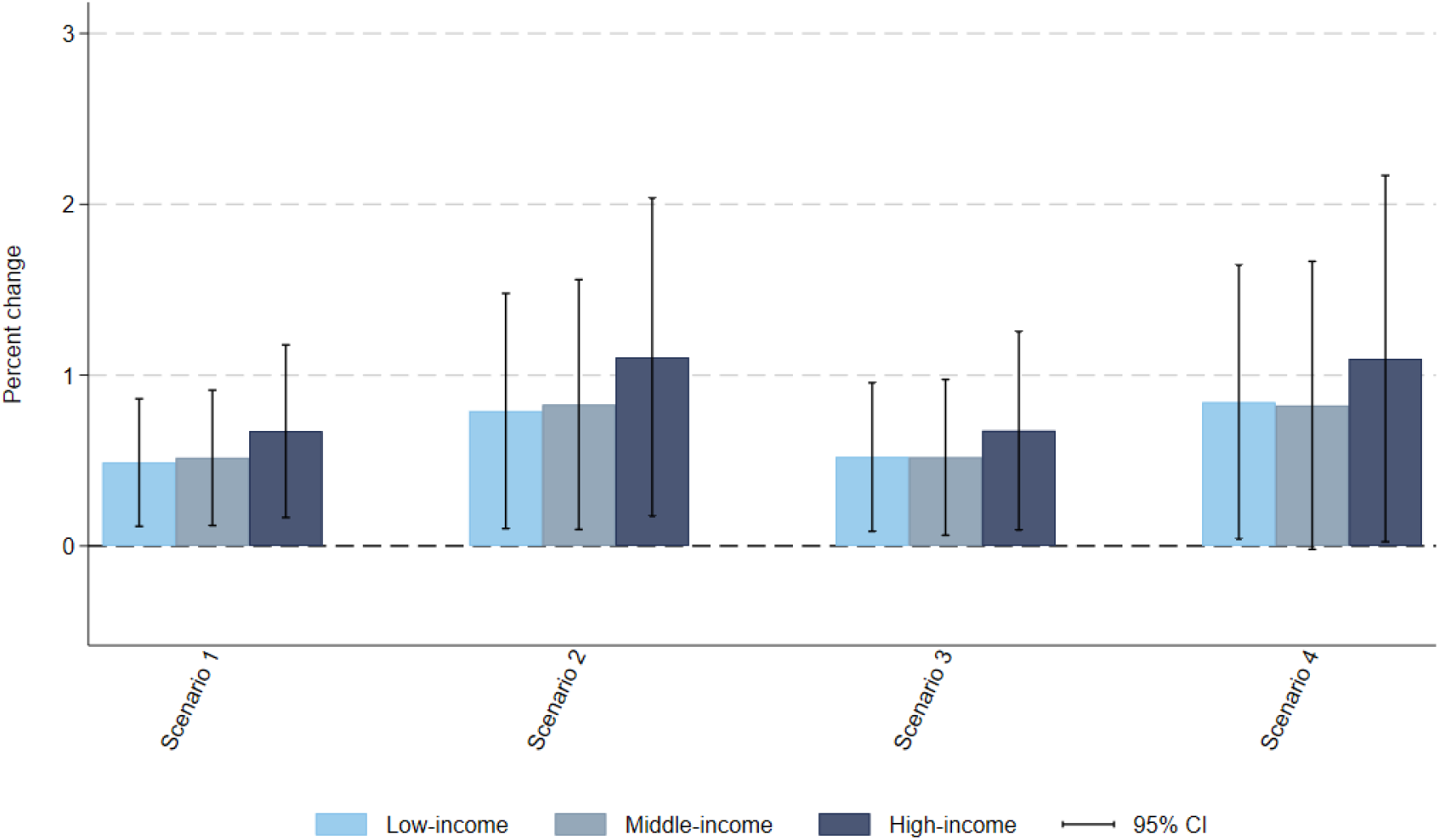
Sensitivity scenario: Passthrough. Impact of fiscal policy scenarios on average total household expenditure on food and beverages, by income group. Notes: This is a sensitivity analysis using an incomplete passthrough of tax rate changes to prices of 80%. Vertical segments represent the 95% confidence intervals. Survey weighted. Scenario 1: defining items for which GST rate is increased to 28% based on the definition of foods and beverages high in fat, sugar, and sodium by the Food Safety and Standards Authority of India [2]; Scenario 2: adding a 12% top-up to the tax rate applied on HFSS foods and beverages in Scenario 1. Scenario 3: scenario defining items for which GST rate is increased to 28% if their nutrient content is above at least one of the respective thresholds set by the World Health Organization South East Asia Region nutrient profile model [3]; Scenarios 4: adding a 12% top-up to the tax rate applied on HFSS foods and beverages in Scenario 3. GST: Goods and Services Tax. HFSS: High in fat, sodium, and sugar.

**Figure B3.**
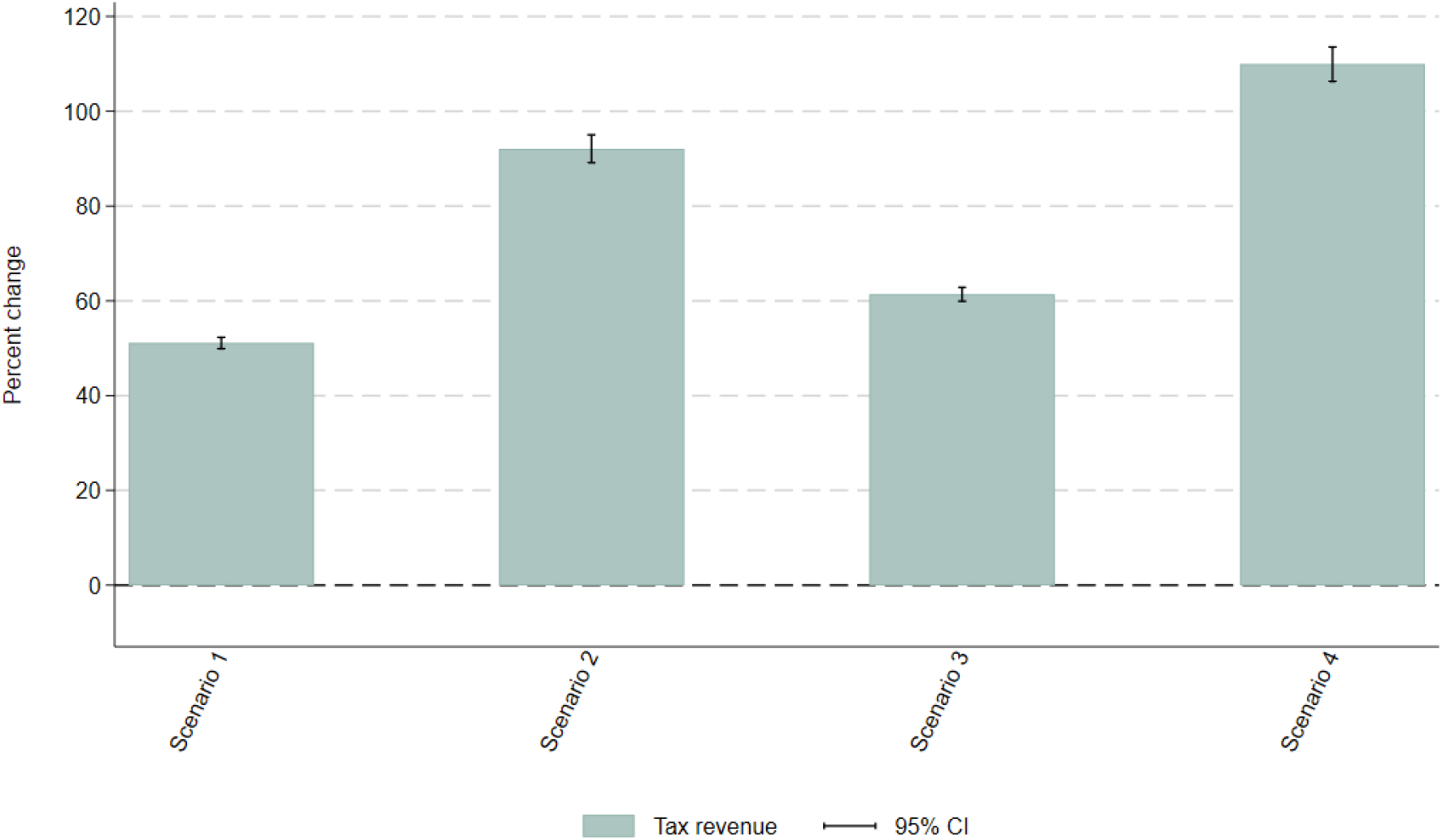
Sensitivity scenario: Passthrough. Impact of fiscal policy scenarios on government tax revenue from foods and beverages. Notes: This is a sensitivity analysis using an incomplete passthrough of tax rate changes to prices of 80%. Vertical segments represent the 95% confidence intervals. Survey weighted. Scenario 1: defining items for which GST rate is increased to 28% based on the definition of foods and beverages high in fat, sugar, and sodium by the Food Safety and Standards Authority of India [2]; Scenario 2: adding a 12% top-up to the tax rate applied on HFSS foods and beverages in Scenario 1. Scenario 3: scenario defining items for which GST rate is increased to 28% if their nutrient content is above at least one of the respective thresholds set by the World Health Organization South East Asia Region nutrient profile model [3]; Scenarios 4: adding a 12% top-up to the tax rate applied on HFSS foods and beverages in Scenario 3. GST: Goods and Services Tax. HFSS: High in fat, sodium, and sugar.

**Figure B4.**
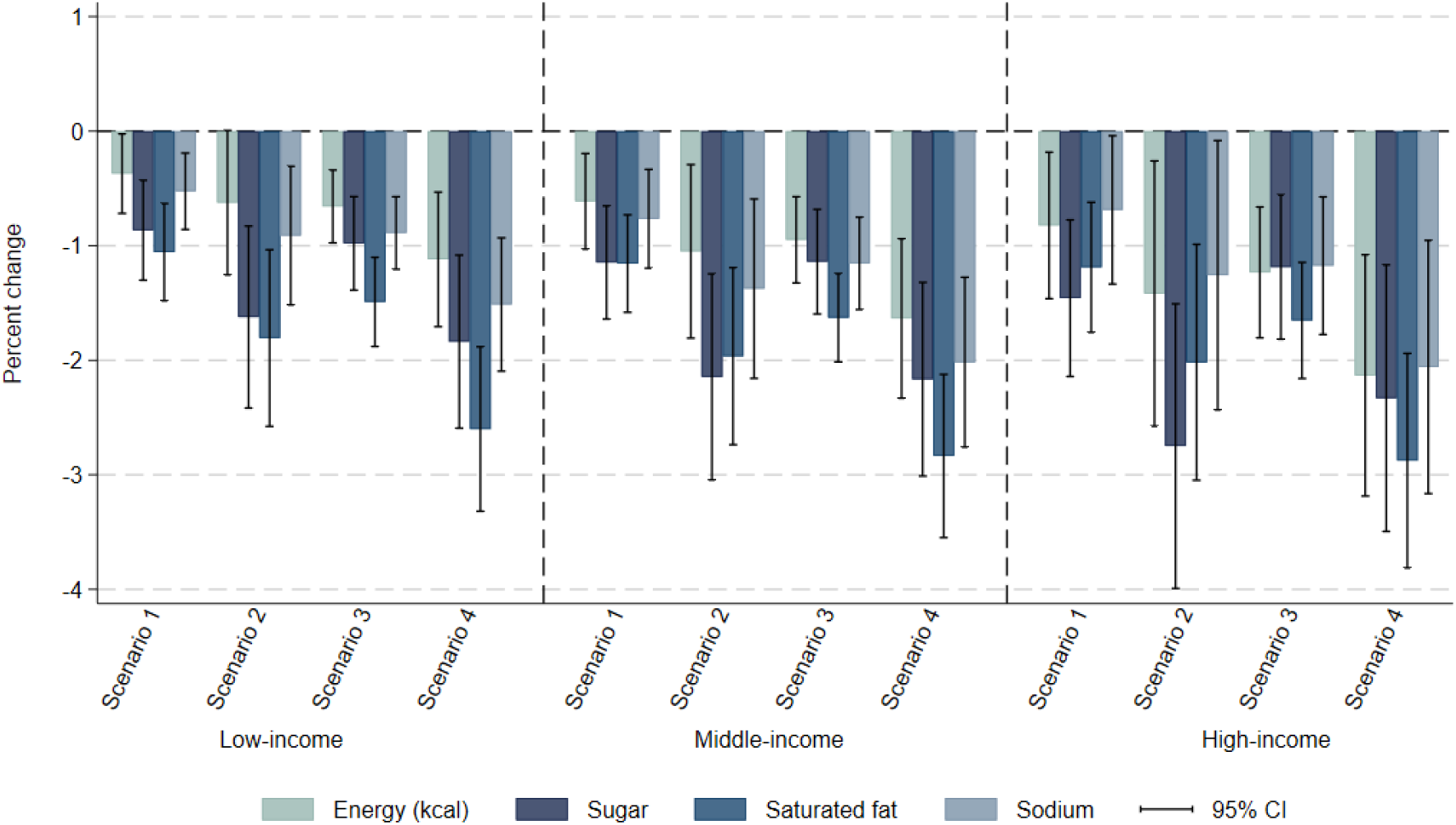
Sensitivity scenario: Reformulation. Immediate impact of fiscal policy scenarios on average daily energy and nutrient intake, by income group. Notes: This is a sensitivity analysis assuming the reformulation of HFSS food items by the industry. If the nutrient content of an HFSS item is less than 50% above the FSSAI HFSS or WHO SEARO NPM threshold for one or several nutrients (among sugar, sodium, and saturated fat), the industry is assumed to reduce the nutrient content of the item for it not to be considered HFSS and avoid the tax. This reformulation is modeled to take place before the introduction of the tax. Vertical segments represent the 95% confidence intervals. Survey weighted. Scenario 1: defining items for which GST rate is increased to 28% based on the definition of foods and beverages high in fat, sugar, and sodium by the Food Safety and Standards Authority of India [2]; Scenario 2: adding a 12% top-up to the tax rate applied on HFSS foods and beverages in Scenario 1. Scenario 3: scenario defining items for which GST rate is increased to 28% if their nutrient content is above at least one of the respective thresholds set by the World Health Organization South East Asia Region nutrient profile model [3]; Scenarios 4: adding a 12% top-up to the tax rate applied on HFSS foods and beverages in Scenario 3. GST: Goods and Services Tax. HFSS: High in fat, sodium, and sugar.

**Figure B5.**
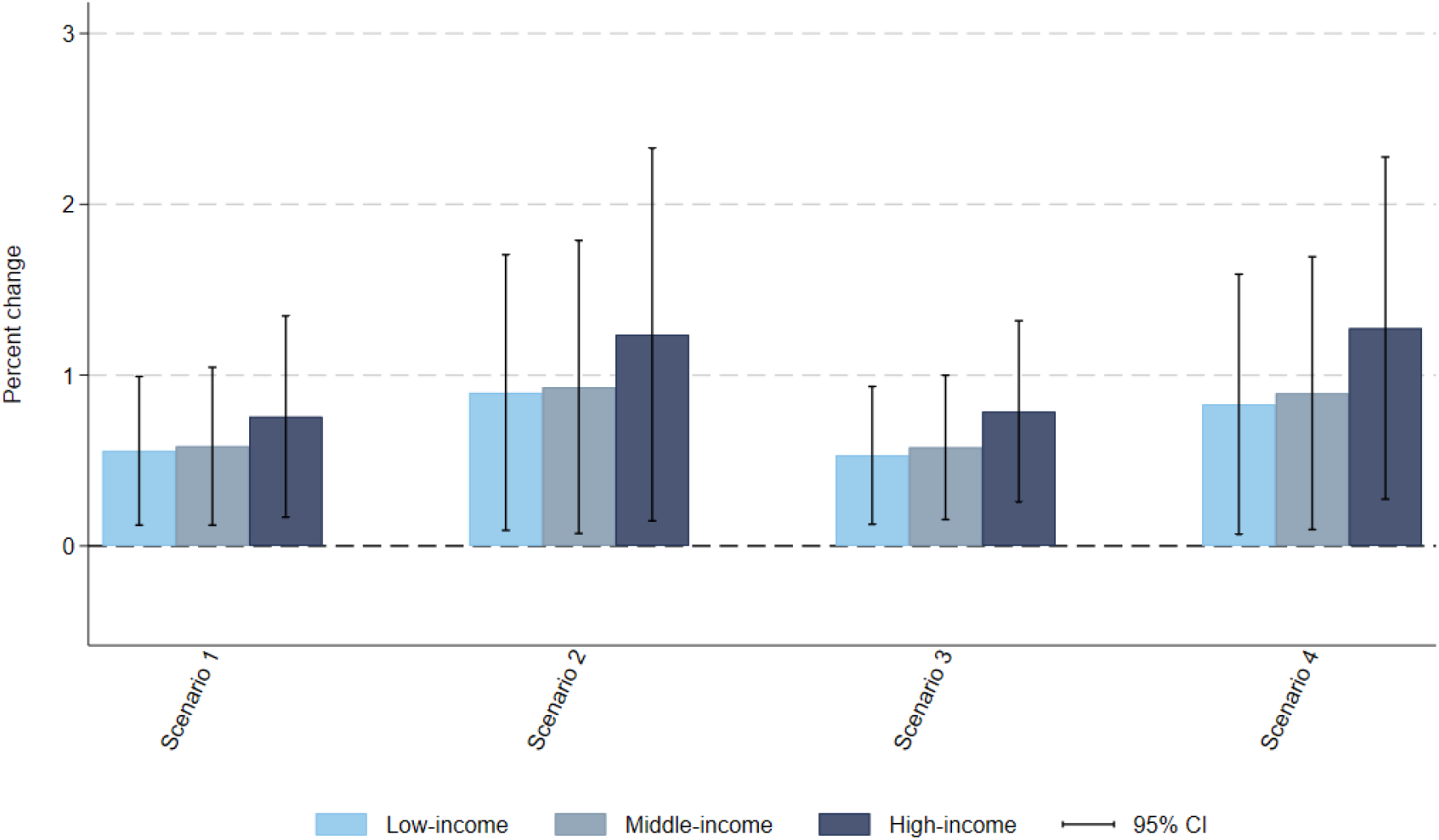
Sensitivity scenario: Reformulation. Impact of fiscal policy scenarios on average total household expenditure on food and beverages, by income group. Notes: This is a sensitivity analysis assuming the reformulation of HFSS food items by the industry. If the nutrient content of an HFSS item is less than 50% above the FSSAI HFSS or WHO SEARO NPM threshold for one or several nutrients (among sugar, sodium, and saturated fat), the industry is assumed to reduce the nutrient content of the item for it not to be considered HFSS and avoid the tax. This reformulation is modeled to take place before the introduction of the tax. Vertical segments represent the 95% confidence intervals. Survey weighted. Scenario 1: defining items for which GST rate is increased to 28% based on the definition of foods and beverages high in fat, sugar, and sodium by the Food Safety and Standards Authority of India [2]; Scenario 2: adding a 12% top-up to the tax rate applied on HFSS foods and beverages in Scenario 1. Scenario 3: scenario defining items for which GST rate is increased to 28% if their nutrient content is above at least one of the respective thresholds set by the World Health Organization South East Asia Region nutrient profile model [3]; Scenarios 4: adding a 12% top-up to the tax rate applied on HFSS foods and beverages in Scenario 3. GST: Goods and Services Tax. HFSS: High in fat, sodium, and sugar.

**Figure B6.**
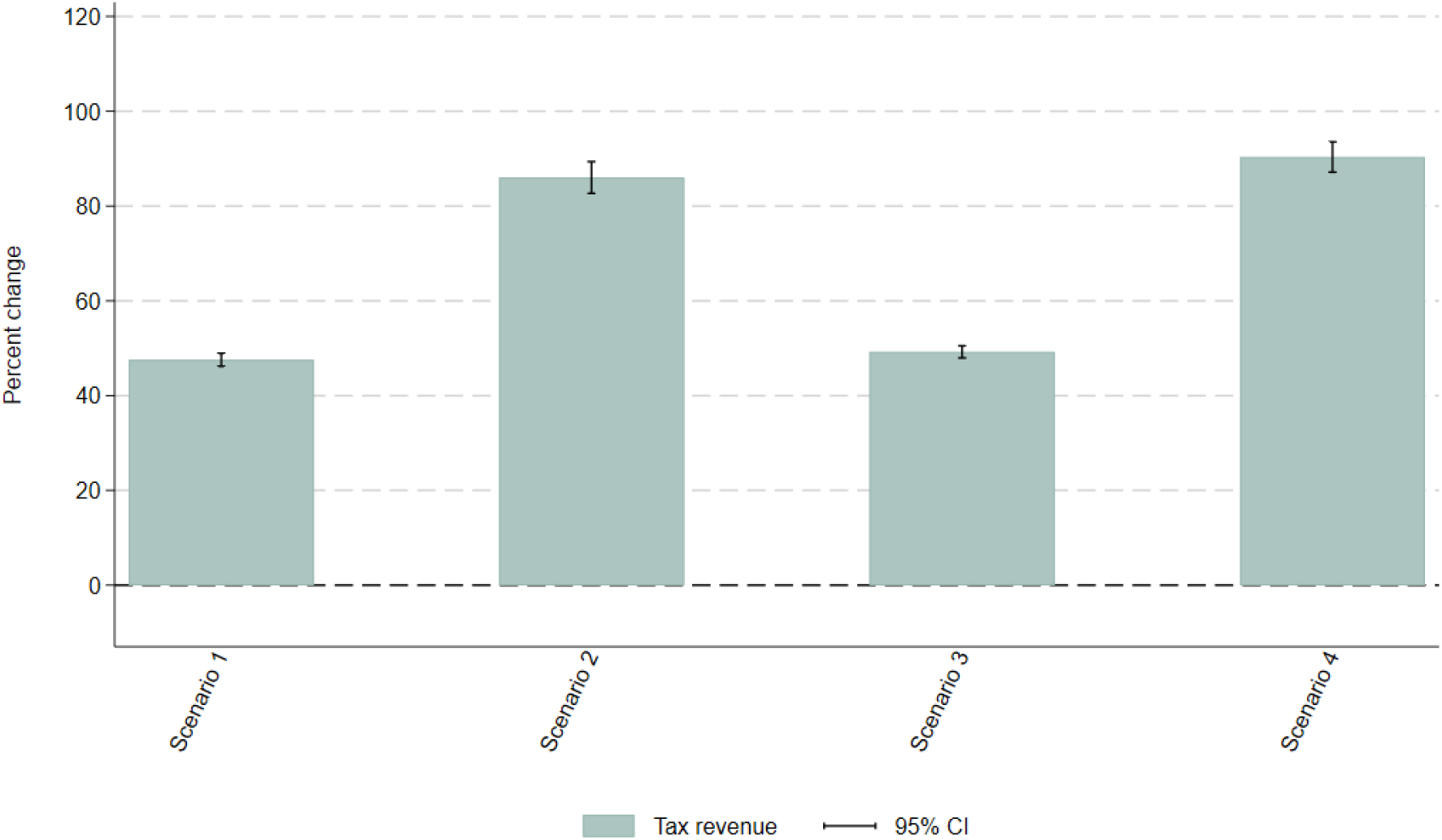
Sensitivity scenario: Reformulation. Impact of fiscal policy scenarios on government tax revenue from foods and beverages. Notes: This is a sensitivity analysis assuming the reformulation of HFSS food items by the industry. If the nutrient content of an HFSS item is less than 50% above the FSSAI HFSS or WHO SEARO NPM threshold for one or several nutrients (among sugar, sodium, and saturated fat), the industry is assumed to reduce the nutrient content of the item for it not to be considered HFSS and avoid the tax. This reformulation is modeled to take place before the introduction of the tax. Vertical segments represent the 95% confidence intervals. Survey weighted. Scenario 1: defining items for which GST rate is increased to 28% based on the definition of foods and beverages high in fat, sugar, and sodium by the Food Safety and Standards Authority of India [2]; Scenario 2: adding a 12% top-up to the tax rate applied on HFSS foods and beverages in Scenario 1. Scenario 3: scenario defining items for which GST rate is increased to 28% if their nutrient content is above at least one of the respective thresholds set by the World Health Organization South East Asia Region nutrient profile model [3]; Scenarios 4: adding a 12% top-up to the tax rate applied on HFSS foods and beverages in Scenario 3. GST: Goods and Services Tax. HFSS: High in fat, sodium, and sugar.

**Figure B7.**
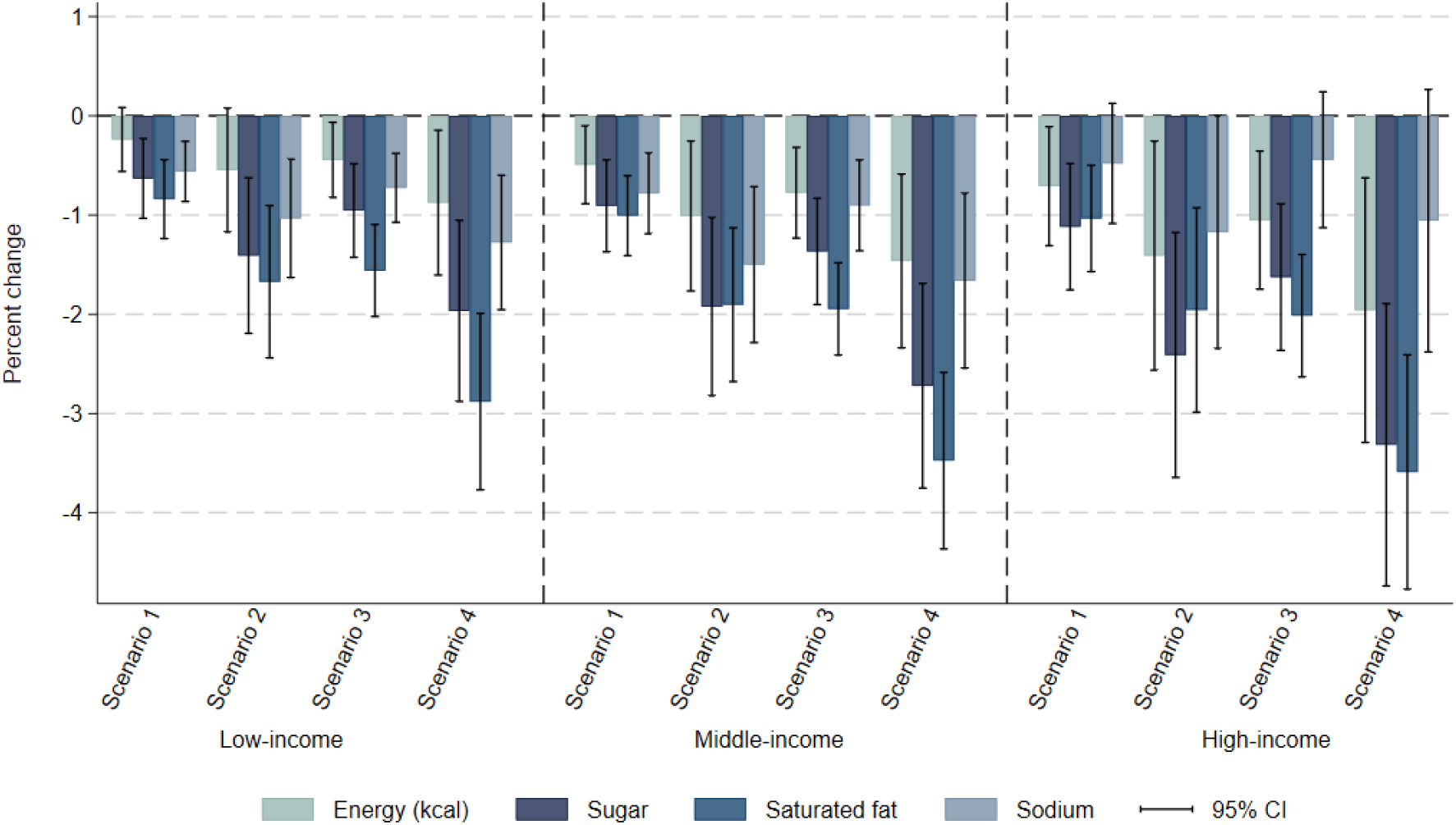
Sensitivity scenario: Subsidy. Immediate impact of fiscal policy scenarios on average daily energy and nutrient intake, by income group. Notes: This is a sensitivity analysis additionally zero-rating F&V and pulses (GST rate = 0%). Most F&V are already zero-rated at baseline so most of the subsidy effect comes from pulses of which many are taxed with a GST rate of 5% at baseline (**Figure A1**). Vertical segments represent the 95% confidence intervals. Survey weighted. Scenario 1: defining items for which GST rate is increased to 28% based on the definition of foods and beverages high in fat, sugar, and sodium by the Food Safety and Standards Authority of India [2]; Scenario 2: adding a 12% top-up to the tax rate applied on HFSS foods and beverages in Scenario 1. Scenario 3: scenario defining items for which GST rate is increased to 28% if their nutrient content is above at least one of the respective thresholds set by the World Health Organization South East Asia Region nutrient profile model [3]; Scenarios 4: adding a 12% top-up to the tax rate applied on HFSS foods and beverages in Scenario 3. GST: Goods and Services Tax. HFSS: High in fat, sodium, and sugar.

**Figure B8.**
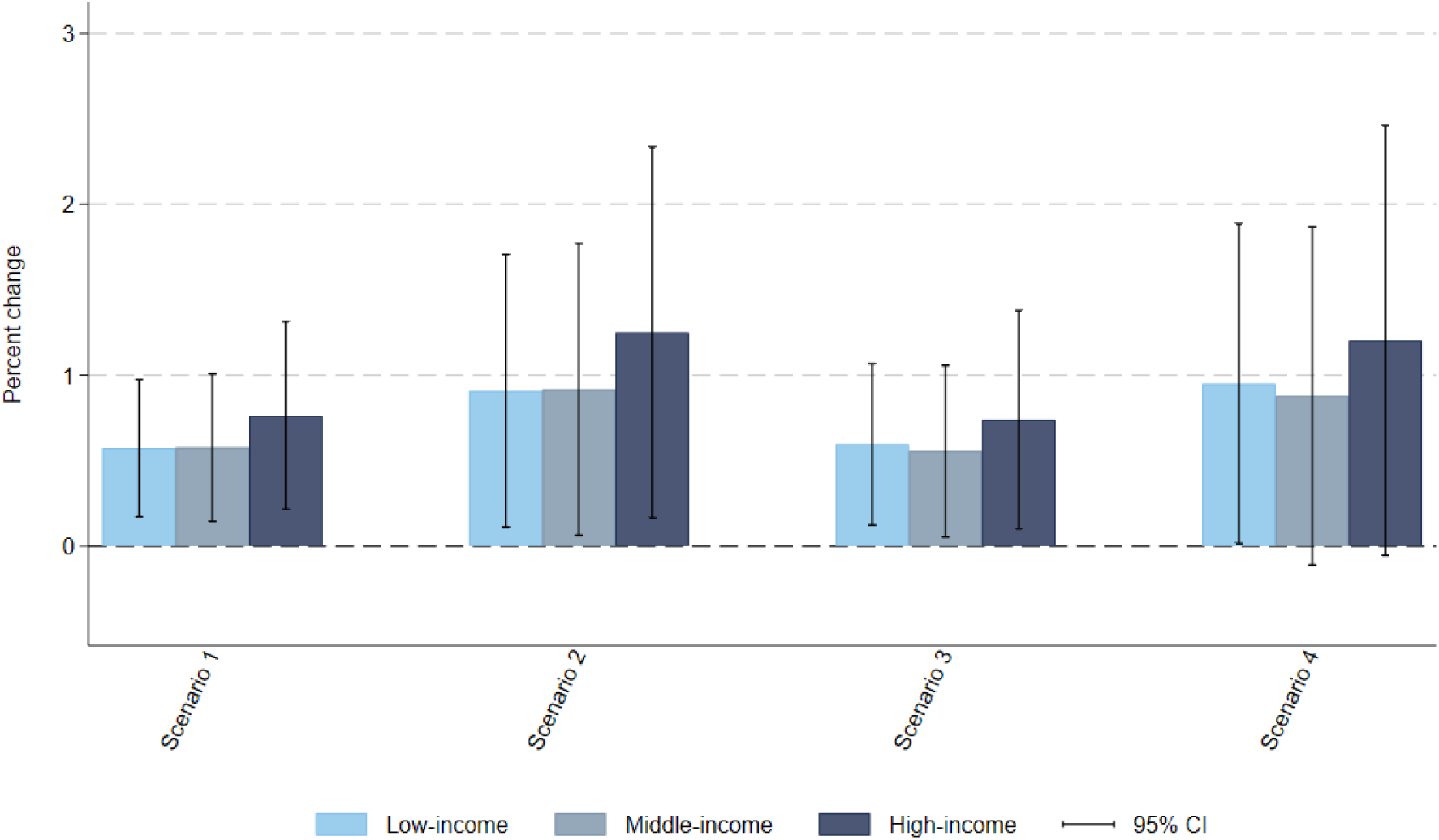
Sensitivity scenario: Subsidy. Impact of fiscal policy scenarios on average total household expenditure on food and beverages, by income group. Notes: This is a sensitivity analysis additionally zero-rating F&V and pulses (GST rate = 0%). Most F&V are already zero-rated at baseline so most of the subsidy effect comes from pulses of which many are taxed with a GST rate of 5% at baseline (**Figure A1**). Vertical segments represent the 95% confidence intervals. Survey weighted. Scenario 1: defining items for which GST rate is increased to 28% based on the definition of foods and beverages high in fat, sugar, and sodium by the Food Safety and Standards Authority of India [2]; Scenario 2: adding a 12% top-up to the tax rate applied on HFSS foods and beverages in Scenario 1. Scenario 3: scenario defining items for which GST rate is increased to 28% if their nutrient content is above at least one of the respective thresholds set by the World Health Organization South East Asia Region nutrient profile model [3]; Scenarios 4: adding a 12% top-up to the tax rate applied on HFSS foods and beverages in Scenario 3. GST: Goods and Services Tax. HFSS: High in fat, sodium, and sugar.

**Figure B9.**
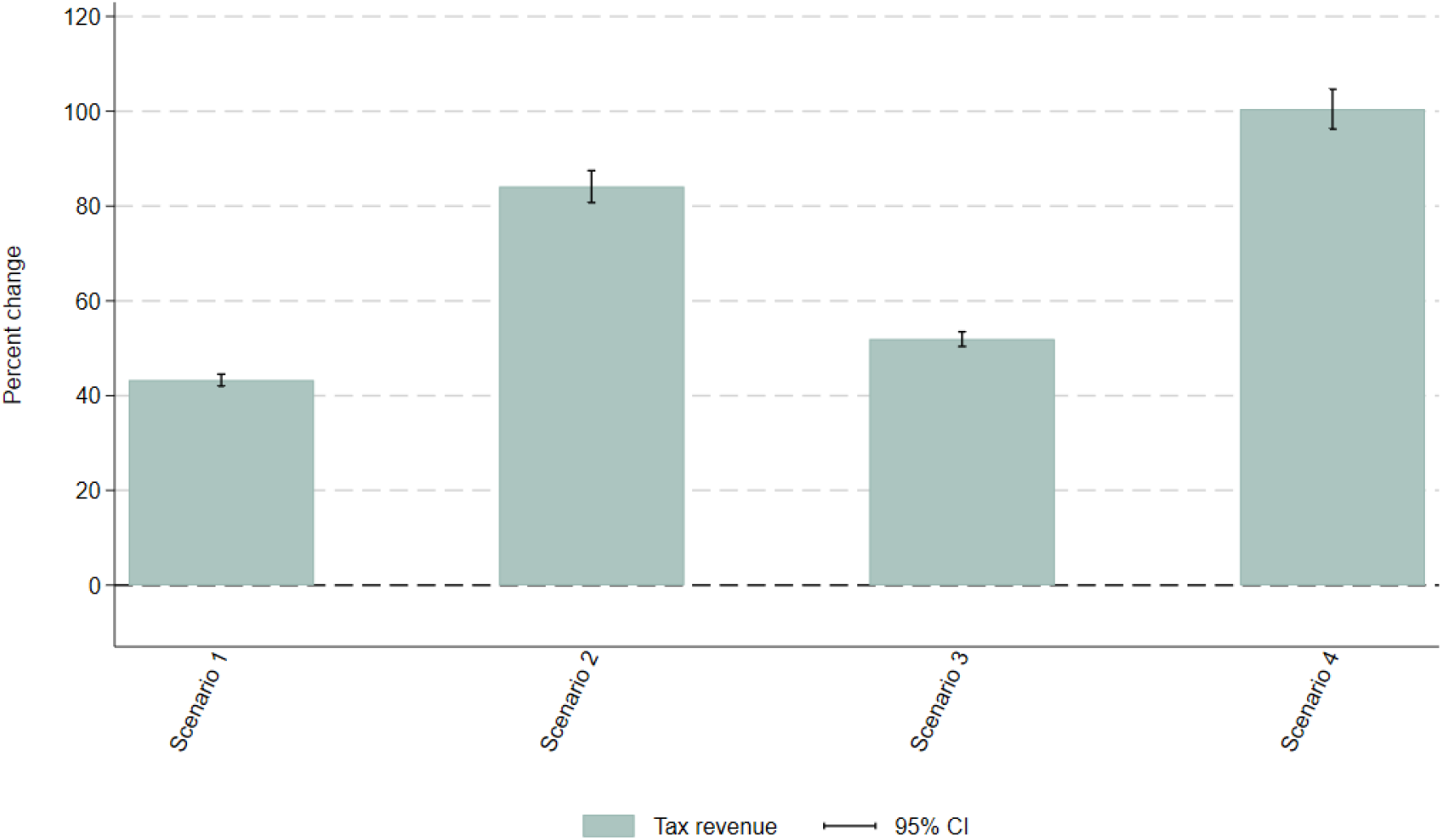
Sensitivity scenario: Subsidy. Impact of fiscal policy scenarios on government tax revenue from foods and beverages. Notes: This is a sensitivity analysis additionally zero-rating F&V and pulses (GST rate = 0%). Most F&V are already zero-rated at baseline so most of the subsidy effect comes from pulses of which many are taxed with a GST rate of 5% at baseline (**Figure A1**). Vertical segments represent the 95% confidence intervals. Survey weighted. Scenario 1: defining items for which GST rate is increased to 28% based on the definition of foods and beverages high in fat, sugar, and sodium by the Food Safety and Standards Authority of India [2]; Scenario 2: adding a 12% top-up to the tax rate applied on HFSS foods and beverages in Scenario 1. Scenario 3: scenario defining items for which GST rate is increased to 28% if their nutrient content is above at least one of the respective thresholds set by the World Health Organization South East Asia Region nutrient profile model [3]; Scenarios 4: adding a 12% top-up to the tax rate applied on HFSS foods and beverages in Scenario 3. GST: Goods and Services Tax. HFSS: High in fat, sodium, and sugar.

### Supporting information file 3

#### Appendix C. Almost Ideal Demand System (AIDS) demand model

Appendix C forms part of the revised submission.

Supplement to: Roche M, Zhu J, Olney J, Laydon DJ, Joe W, Sharma M, Steele L, Sassi F. Taxation of foods high in saturated fat, sugar, and sodium in India: A modelling study of health and economic impacts. Submitted after revision on 19 September 2025.

##### C1. The model

As with most other household budget surveys, the NSSO Household Consumption Expenditure survey 2023-24 does not provide price information, thus we rely on unit values (i.e., expenditure divided by quantity). However, unit values are subject to measurement errors in both expenditure and quantity and may lead to quality shading, where consumers may not only adjust quantity but also the quality of the goods they buy as a response to price changes. We thus estimate consumer responses to food price changes (technically, own- and cross-price elasticities of demand) using Deaton (1988)’s Almost Ideal Demand System (AIDS) model adjusting for quality shading and measurement error [1].

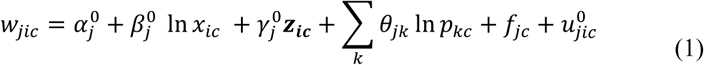

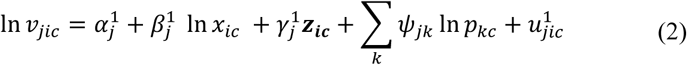

where *w*_*jic*_ represents the share of total expenditure on food and beverages (*x*_*ic*_) that household *i* in cluster *c* spends on good *j*; *p*_*kc*_ is the price of good *k* which does not vary in cluster *c*; *f*_*jc*_ are unobservable cluster fixed effects; *v*_*jic*_ is the unit value of good *j*; and *u*_*jic*_ are idiosyncratic error terms. ***z***_***ic***_ is a vector of household characteristics, including the logarithm of household size, the sector of the household (urban vs. rural), the sex of the head of household and a dummy for completion of primary education, and dummies capturing the religion of the household. We remove regional and seasonal effects using dummies for the six Indian administrative zones and the quarter of the year in which the household reports information.

The specification nests rich income (Engel-curve) responses and flexible cross-price substitution patterns while remaining consistent with standard consumer theory. Adding-up, homogeneity, and Slutsky symmetry are imposed so the estimates are integrable and interpretable as arising from well-behaved preferences in the price-independent generalized logarithmic preferences class (PIGLOG) [2].

The model assumes spatially varying prices, where all households within a near geographical area face the same price. As defined by NSSO, a cluster is equivalent to a village, or an urban block, surveyed during the same period. Any within-cluster variation in unit values is due to differences in the quality of the purchased items.

Spatial variations in unit values between clusters provide an identification strategy to avoid the endogeneity of prices and address quality shading [1]. We argue this assumption is justified in a middle-income country like India where transport is more difficult and relatively costlier, and markets are not always well-integrated [2]. To abide by our main assumption of spatially varying prices, we force households to face the same price if they are drawn from the same cluster, regardless of their income group. This takes care of a potential endogeneity in average prices that might arise if households from different income groups face different average prices despite being in the same cluster [3]. We drop clusters not having at least two households consuming at least an item for each group. Following the demand system literature [4], we normalize the *numeraire* group’s unit value to one.

We recover uncompensated price elasticity of demand estimates at the means through a three-step method including demeaned within-cluster regressions, between-cluster regressions of adjusted budget share on adjusted unit values estimated in the first step using an error-in-variable estimator, and finally separating quality and price effects.

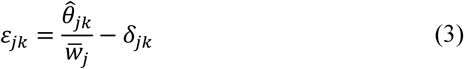

where *δ*_*jk*_ is the Kronecker delta equal to one if *j* = *k* (i.e., for own-price elasticities) or zero otherwise.

This approach has been used extensively to model the demand for food in low- and middle-income countries [5]. A key strength is its rigorous treatment of measurement noise. By residualizing within clusters, averaging to the cluster, and then applying measurement-error corrections that scale with buyer and household counts, it strips out spurious variation that would otherwise attenuate price effects while also tempering quality shading.

Nevertheless, the use of this model with NSSO Household Consumption Expenditure survey data has some limitations. First, censoring is only mitigated, not solved. Unit-value regressions exclude non-buyers while budget share regressions include everyone. Clusters with very few buyers are dropped to avoid unstable means and the measurement-error corrections down-weight clusters where buyer counts are small. These steps reduce the distortion created by many zeros, but they cannot remove selection bias if the decision not to buy is correlated with unobserved preferences, access, or local conditions that also relate to prices. Second, using household budget survey data, the underlying demand model cannot account for cross-price effects within food groups. Future research using detailed consumer panel data (e.g., home-scan) could estimate demand systems using more disaggregated groups, for example, by bundling items based on nutritional impact. Third, the NSSO also lacks information on household disposable income, which is commonly used as an instrumental variable to address the endogeneity in total expenditure arising from simultaneity bias [6].

Details regarding the microeconomic foundations of Deaton (1988)’s AIDS model, as well as its derivation and the steps involved in its estimation, have been described elsewhere [2]. The paper uses uncompensated price elasticities as opposed to compensated price elasticities, as uncompensated elasticities measure the effect of a price change for one good while keeping income and the prices of other goods constant, thereby reflecting both income and substitution effects.

##### C.2. Food groups

We estimate the model parameters for eleven groups: cereals, dairy, pulses, edible oils and spices (including salt and raw sugar), fruits and vegetables (including nuts, hereafter referred to as F&V), animal meats (fresh), packaged processed foods, sweets, SSBs, non-SSBs, and a numeraire group containing food-away-from-home and non-food expenditure. While we account for served processed foods (on-trade sector, i.e., food-away-from-home, like bars, restaurants, etc.) in the estimation of total nutrient intake, we do not include this aggregate as a standalone food group in our demand system. Instead, we group it with non-food expenditure within the *numeraire*.

The GST is applied differently to the on-trade sector in India, based on industry categorisation or types of premises rather than the specific type of food or beverage items served. Therefore, the GST scenarios simulated in this study only apply to the off-trade sector. In addition, food-away-from-home is not well captured in the NSS Household Consumption Expenditure survey, including only a limited number of broadly defined items (cooked meals purchased, cooked meals received free in the workplace, cooked meals received as assistance, cooked snacks purchased, other served processed food) [7]. While the simulated GST scenarios may impact the price of raw materials, it is not possible, given the available data, to estimate the extent to which this may affect the price of served processed foods.

**Table C1.** Estimated average income elasticity estimates, by income group.

|  | Low-income | Middle-income | High-income |
| --- | --- | --- | --- |
| Cereals | 0.329*** | 0.271*** | 0.122*** |
|  | (0.015) | (0.016) | (0.006) |
| Dairy | 1.175*** | 0.752*** | 0.496*** |
|  | (0.017) | (0.018) | (0.007) |
| Pulses | 0.450*** | 0.386*** | 0.281*** |
|  | (0.012) | (0.013) | (0.006) |
| Oils & spices | 0.404*** | 0.327*** | 0.229*** |
|  | (0.008) | (0.010) | (0.005) |
| F&V | 0.564*** | 0.470*** | 0.344*** |
|  | (0.009) | (0.011) | (0.005) |
| Animal meats, fresh | 0.980*** | 0.734*** | 0.477*** |
|  | (0.017) | (0.022) | (0.008) |
| Packaged proc. foods | 1.076*** | 0.861*** | 0.610*** |
|  | (0.015) | (0.017) | (0.008) |
| Sweets | 0.842*** | 0.846*** | 0.665*** |
|  | (0.018) | (0.025) | (0.011) |
| SSBs | 1.962*** | 1.560*** | 1.162*** |
|  | (0.042) | (0.048) | (0.020) |
| Non-SSBs | 1.093*** | 0.992*** | 0.867*** |
|  | (0.022) | (0.028) | (0.013) |

Notes: Estimated based on NSS Household Consumption Expenditure survey 2022-23 [8] and using Deaton (1988)'s quality-adjusted Almost Ideal Demand System model [1]. Bootstrapped standard errors after 300 replications. Vertical segments represent the 95% confidence intervals. Pack.: Packaged; proc.: processed; SSBs: sugar-sweetened beverages.

**Figure C1.**
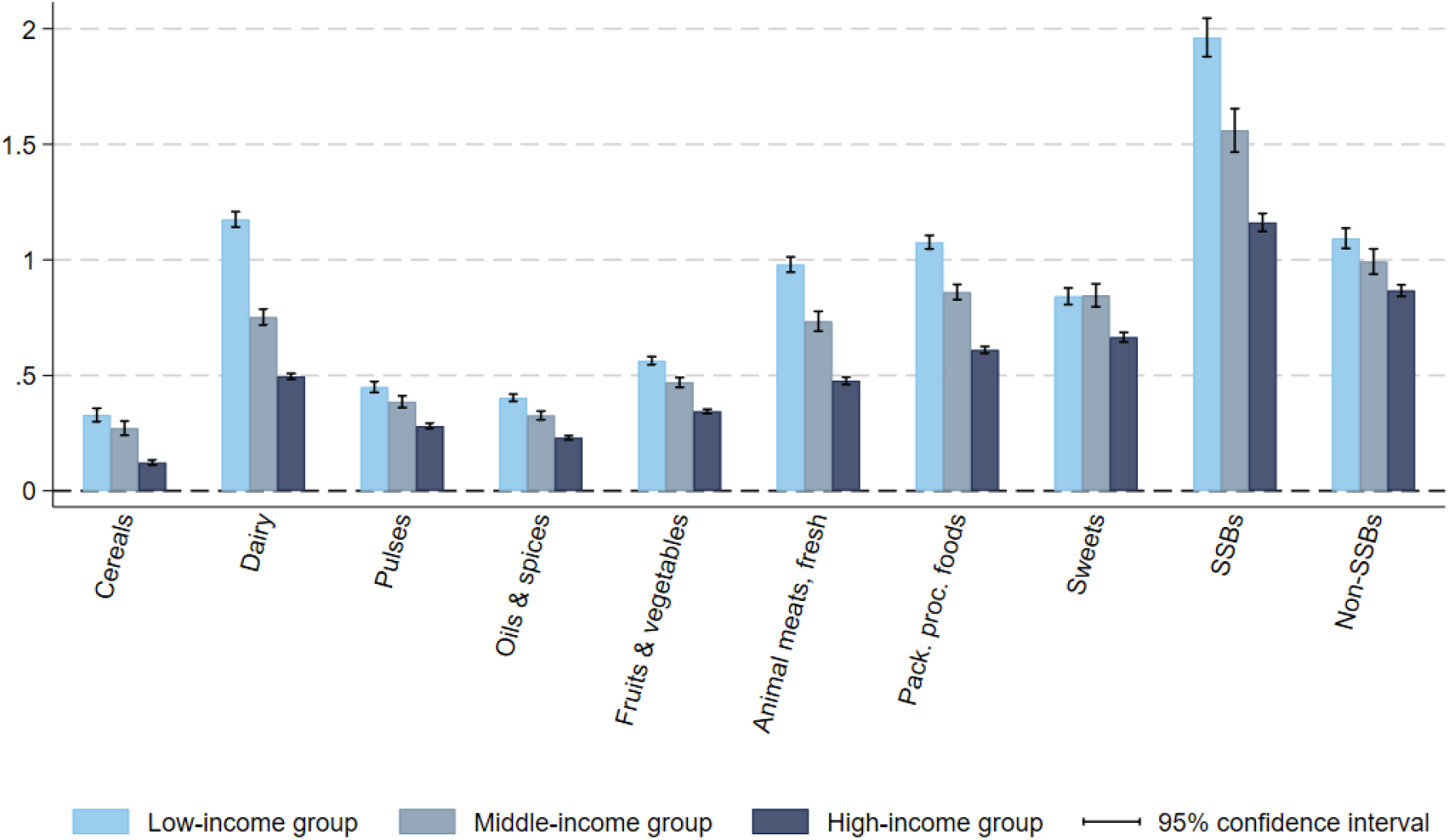
Estimated average income elasticity estimates, by income group. Notes: Estimated based on NSS Household Consumption Expenditure survey 2022-23 [8] and using Deaton (1988)’s quality-adjusted Almost Ideal Demand System model [1]. Bootstrapped standard errors after 300 replications. Vertical segments represent the 95% confidence intervals. Pack.: Packaged; proc.: processed; SSBs: sugar-sweetened beverages.

**Table C2.**
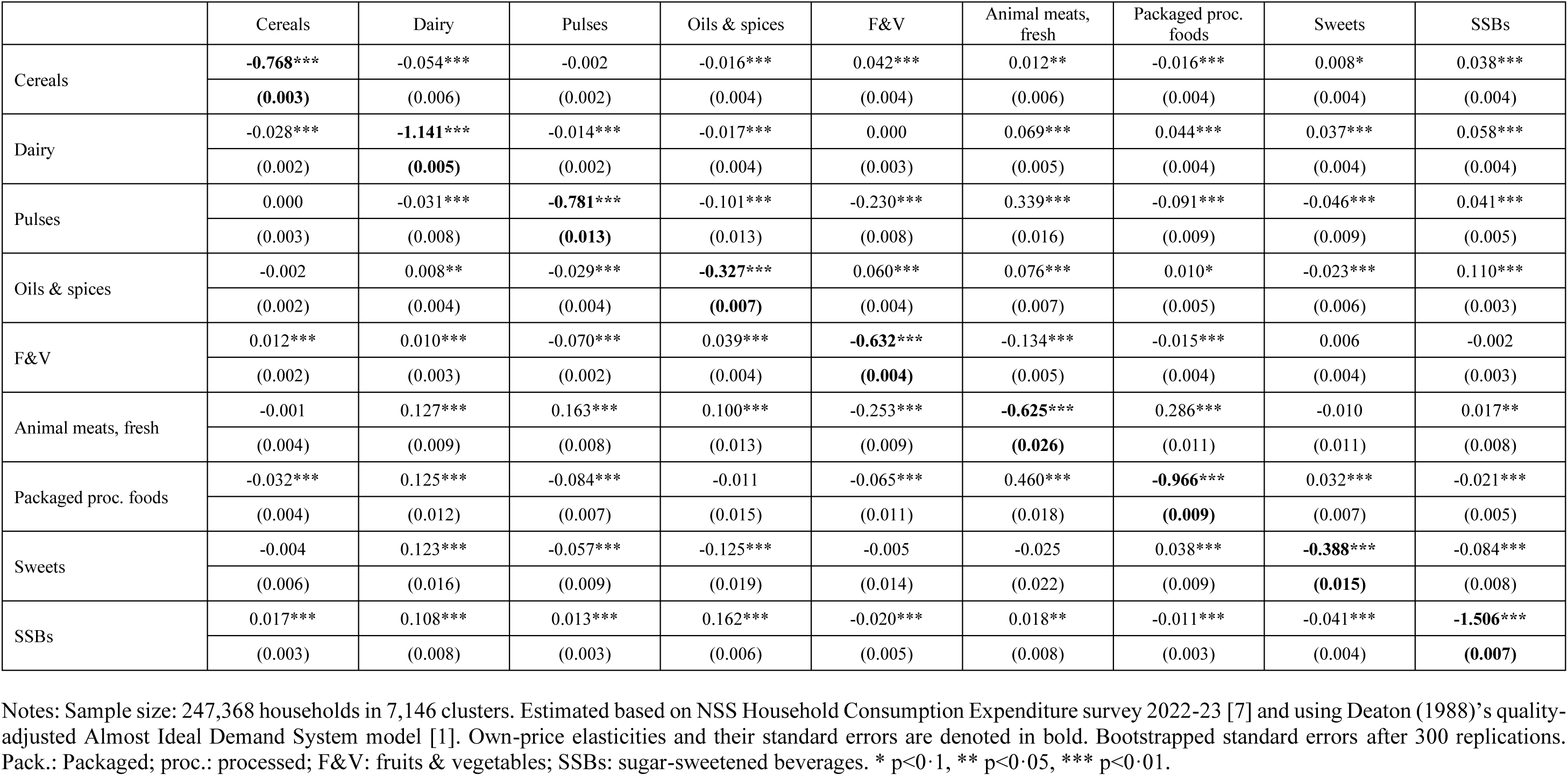
Robustness: Estimated average price elasticities using aggregated beverage grouping, full sample.

|  | Cereals | Dairy | Pulses | Oils & spices | F&V | Animal meats, fresh | Packaged proc. foods | Sweets | SSBs |
| --- | --- | --- | --- | --- | --- | --- | --- | --- | --- |
| Cereals | **-0.768\*\*\*** | -0.054*** | -0.002 | -0.016*** | 0.042*** | 0.012** | -0.016*** | 0.008* | 0.038*** |
|  | **(0.003)** | (0.006) | (0.002) | (0.004) | (0.004) | (0.006) | (0.004) | (0.004) | (0.004) |
| Dairy | -0.028*** | **-1.141\*\*\*** | -0.014*** | -0.017*** | 0.000 | 0.069*** | 0.044*** | 0.037*** | 0.058*** |
|  | (0.002) | **(0.005)** | (0.002) | (0.004) | (0.003) | (0.005) | (0.004) | (0.004) | (0.004) |
| Pulses | 0.000 | -0.031*** | **-0.781\*\*\*** | -0.101*** | -0.230*** | 0.339*** | -0.091*** | -0.046*** | 0.041*** |
|  | (0.003) | (0.008) | **(0.013)** | (0.013) | (0.008) | (0.016) | (0.009) | (0.009) | (0.005) |
| Oils & spices | -0.002 | 0.008** | -0.029*** | **-0.327\*\*\*** | 0.060*** | 0.076*** | 0.010* | -0.023*** | 0.110*** |
|  | (0.002) | (0.004) | (0.004) | **(0.007)** | (0.004) | (0.007) | (0.005) | (0.006) | (0.003) |
| F&V | 0.012*** | 0.010*** | -0.070*** | 0.039*** | **-0.632\*\*\*** | -0.134*** | -0.015*** | 0.006 | -0.002 |
|  | (0.002) | (0.003) | (0.002) | (0.004) | **(0.004)** | (0.005) | (0.004) | (0.004) | (0.003) |
| Animal meats, fresh | -0.001 | 0.127*** | 0.163*** | 0.100*** | -0.253*** | **-0.625\*\*\*** | 0.286*** | -0.010 | 0.017** |
|  | (0.004) | (0.009) | (0.008) | (0.013) | (0.009) | **(0.026)** | (0.011) | (0.011) | (0.008) |
| Packaged proc. foods | -0.032*** | 0.125*** | -0.084*** | -0.011 | -0.065*** | 0.460*** | **-0.966\*\*\*** | 0.032*** | -0.021*** |
|  | (0.004) | (0.012) | (0.007) | (0.015) | (0.011) | (0.018) | **(0.009)** | (0.007) | (0.005) |
| Sweets | -0.004 | 0.123*** | -0.057*** | -0.125*** | -0.005 | -0.025 | 0.038*** | **-0.388\*\*\*** | -0.084*** |
|  | (0.006) | (0.016) | (0.009) | (0.019) | (0.014) | (0.022) | (0.009) | **(0.015)** | (0.008) |
| SSBs | 0.017*** | 0.108*** | 0.013*** | 0.162*** | -0.020*** | 0.018** | -0.011*** | -0.041*** | **-1.506\*\*\*** |
|  | (0.003) | (0.008) | (0.003) | (0.006) | (0.005) | (0.008) | (0.003) | (0.004) | **(0.007)** |

Notes: Sample size: 247,368 households in 7,146 clusters. Estimated based on NSS Household Consumption Expenditure survey 2022-23 [7] and using Deaton (1988)'s quality-adjusted Almost Ideal Demand System model [1]. Own-price elasticities and their standard errors are denoted in bold. Bootstrapped standard errors after 300 replications. Pack.: Packaged; proc.: processed; F&V: fruits & vegetables; SSBs: sugar-sweetened beverages. \* p<0.1, \*\* p<0.05, \*\*\* p<0.01.

**Table C3.** Robustness: Estimated average price elasticities using non-food CPI as unit value for numeraire, full sample.

|  | Cereals | Dairy | Pulses | Oils & spices | F&V | Animal meats, fresh | Packaged proc. foods | Sweets | SSBs | Non-SSBs |
| --- | --- | --- | --- | --- | --- | --- | --- | --- | --- | --- |
| Cereals | **-0.797\*\*\*** | -0.088*** | 0.007*** | -0.026*** | 0.049*** | 0.023*** | -0.028*** | 0.022*** | 0.015** | 0.004 |
|  | **(0.004)** | (0.007) | (0.003) | (0.005) | (0.004) | (0.007) | (0.004) | (0.005) | (0.006) | (0.004) |
| Dairy | -0.038*** | **-1.124\*\*\*** | -0.010*** | -0.003 | -0.024*** | 0.072*** | 0.050*** | 0.057*** | 0.014** | 0.071*** |
|  | (0.002) | **(0.006)** | (0.002) | (0.003) | (0.003) | (0.006) | (0.004) | (0.005) | (0.007) | (0.004) |
| Pulses | 0.013*** | -0.017* | **-0.748\*\*\*** | -0.143*** | -0.170*** | 0.284*** | -0.071*** | -0.032*** | 0.001 | 0.036*** |
|  | (0.004) | (0.009) | **(0.013)** | (0.016) | (0.011) | (0.019) | (0.009) | (0.010) | (0.009) | (0.004) |
| Oils & spices | -0.006*** | 0.023*** | -0.042*** | **-0.367\*\*\*** | 0.061*** | 0.070*** | 0.005 | -0.031*** | 0.016** | 0.075*** |
|  | (0.002) | (0.004) | (0.005) | **(0.009)** | (0.004) | (0.008) | (0.006) | (0.006) | (0.006) | (0.003) |
| F&V | 0.014*** | -0.017*** | -0.051*** | 0.037*** | **-0.642\*\*\*** | -0.127*** | -0.017*** | 0.003 | 0.007 | 0.006** |
|  | (0.002) | (0.003) | (0.003) | (0.004) | **(0.005)** | (0.006) | (0.004) | (0.005) | (0.005) | (0.003) |
| Animal meats, fresh | 0.006 | 0.131*** | 0.128*** | 0.085*** | -0.236*** | **-0.630\*\*\*** | 0.265*** | -0.013 | -0.024** | 0.025*** |
|  | (0.005) | (0.011) | (0.009) | (0.012) | (0.010) | **(0.030)** | (0.013) | (0.014) | (0.012) | (0.007) |
| Packaged proc. foods | -0.042*** | 0.138*** | -0.063*** | -0.024 | -0.067*** | 0.414*** | **-0.922\*\*\*** | 0.002 | 0.035*** | -0.011** |
|  | (0.005) | (0.013) | (0.007) | (0.016) | (0.011) | (0.020) | **(0.011)** | (0.008) | (0.008) | (0.005) |
| Sweets | 0.013** | 0.192*** | -0.040*** | -0.141*** | -0.017 | -0.031 | 0.001 | **-0.439\*\*\*** | 0.030*** | -0.073*** |
|  | (0.006) | (0.019) | (0.010) | (0.019) | (0.016) | (0.028) | (0.010) | **(0.019)** | (0.010) | (0.008) |
| SSBs | 0.020 | 0.080 | -0.014 | 0.046 | 0.002 | -0.124** | 0.087*** | 0.059*** | **-1.203\*\*\*** | 0.090*** |
|  | (0.018) | (0.058) | (0.018) | (0.043) | (0.035) | (0.052) | (0.021) | (0.022) | **(0.017)** | (0.020) |
| Non-SSBs | -0.001 | 0.195*** | 0.020*** | 0.151*** | 0.016** | 0.042*** | -0.002 | -0.045*** | 0.036*** | **-1.474\*\*\*** |
|  | (0.004) | (0.011) | (0.003) | (0.007) | (0.007) | (0.010) | (0.005) | (0.005) | (0.006) | **(0.009)** |

Notes: Estimated based on NSS Household Consumption Expenditure survey 2022-23 [7] and using Deaton (1988)'s quality-adjusted Almost Ideal Demand System model [1]. Own-price elasticities and their standard errors are denoted in bold. Bootstrapped standard errors after 300 replications. Pack.: Packaged; proc.: processed; F&V: fruits & vegetables; SSBs: sugar-sweetened beverages. \* p<0.1, \*\* p<0.05, \*\*\* p<0.01.

### Supporting information file 4

#### Appendix D. Health-GPS microsimulation model

Appendix D forms part of the revised submission.

Supplement to: Roche M, Zhu J, Olney J, Laydon DJ, Joe W, Sharma M, Steele L, Sassi F. Taxation of foods high in saturated fat, sugar, and sodium in India: A modelling study of health and economic impacts. Submitted after revision on 19 September 2025.

##### D1. Overview

Health-GPS is a microsimulation model that simulates individuals over time, estimating their risks of developing a variety of NCDs (e.g., stroke) given their exposure to particular risk factors (e.g., excessive sodium intake). Health-GPS then models possible health policies (e.g., changes in taxation on foods high in sodium), that might affect these risk factors, and therefore may impact the incidence and outcomes of NCDs at the population level.

The model creates a synthetic population of individuals that broadly reproduces demographic and socioeconomic characteristics of the population of a given country, or sub-national jurisdiction, and simulates individual life histories from birth to death. Because it simulates a population at the individual level, Health-GPS can capture heterogeneity in risk exposures, diseases, and multi-morbidity patterns. Correlations between risk factor exposures and between diseases are reflected in simulated populations. Health-GPS is calibrated to real-world data to capture relationships between variables while matching to estimated and projected demographic and epidemiological metrics from international databases.

In Health-GPS, a baseline scenario is simulated reflecting expected demographic and epidemiological changes over a time horizon in a given population, to match current demographic projections over that same time horizon. Intervention scenarios are then simulated, reflecting the introduction of health policies applied to targeted individuals and risk factors. Changes in risk exposures will, in turn, impact other risk factors, diseases, and mortality through the model equations. Outcomes simulated through the model typically include, but are not limited to, the prevalence of risk factors, the incidence of, and mortality from, NCDs, years of life lost (YLLs), years lived with disability (YLDs), disability-adjusted life years (DALYs), and life expectancy. Policy impacts are estimated as the difference in these outcomes between the baseline and intervention scenarios. Given the random variation in model outputs due to Health-GPS’ stochastic nature, the model repeats these steps multiple times and averages the differences to obtain the effect of a given health policy. Another way of limiting the impact of stochastic variation on simulation results is to model large population samples.

This appendix is divided into the following sections describing **four key Health-GPS component modules**, illustrated in **Figure D1**:

**1. Demographic module:** births, deaths, immigration, population and socioeconomic status.
**2. Risk factor modules:** including energy balance and high dietary sodium intake in the version used for the study described in this paper.
**3. Disease modules:** we explicitly model the following diseases: asthma; chronic kidney disease; diabetes; ischemic heart disease; and stroke (including intracerebral haemorrhage, ischemic stroke, and subarachnoid haemorrhage). Other diseases are accounted for in a residual mortality aggregate.
**4. Disease burden module:** including metrics used to estimate population-level outcomes of interventions, specifically YLLs, YLDs, and DALYs.

In the last section, we additionally include sources and methods used to estimate the health expenditure impact of interventions.

**Figure D1.**
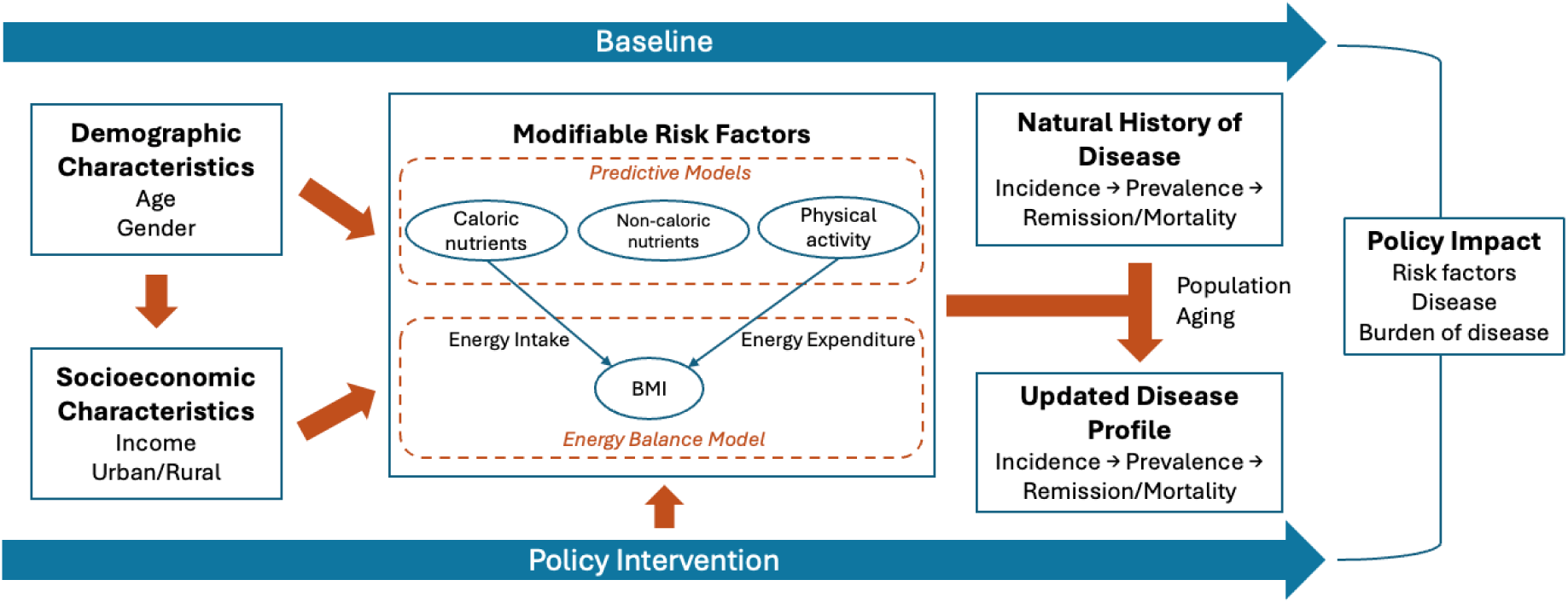
Health-GPS model diagram. Notes: Disease is modelled through three types of events: onset, remission, and fatality, with data on prevalence, incidence, remission, and mortality rates for non-cancerous diseases collected from the IHME Global Burden of Disease. Cancers are modelled using data from the International Agency for Research on Cancer (IARC) on prevalence at 1, 3, and 5 years after diagnosis, incidence, and mortality rates. Cancer survivors beyond five years post-diagnosis are assumed to be in remission.

##### D2. Demographic module

The demographic module manages the yearly numbers of births, deaths, and migrations, as well as socioeconomic status. This module aims to produce a baseline scenario consistent with projected demographic data.

###### D2.1. Data

Health-GPS uses the United Nations (UN)’s World Population Prospects projections for yearly births, deaths, total population, and net migration, for all countries, by age and sex [1]. In the next sections, we explain how these data are calibrated in the demographic module.

###### D2.2. Births and deaths

The demographic module reproduces historical and projected births and deaths in a baseline scenario. The UN estimates and projects birth and death rates for all countries from 1950 until 2100. These estimates and projections are available in five-year intervals, and the module interpolates the data linearly to account for missing years.

These data determine the number of births at the beginning of each year of the simulation, assigning sex based on a sex ratio. Newborns are disease-free at the beginning of the simulation, and their initial risk factor exposures are generated using the initialisation equations of the risk factor modules. For all individuals in Health-GPS, risk and disease profiles are updated yearly until they either die or migrate.

Similarly, UN data provide estimates of the number of deaths in each year of the simulation. Health-GPS calculates an individual’s probability of death each year based on their sex, age and disease profile. The model simulates multiple diseases but allows for death from other non-modelled causes, i.e. residual mortality.

###### D2.3. Residual mortality

Let the *i*^th^ person (who has neither died nor emigrated) have age *a*_*i*_ and sex *g*_*i*_. Let *m*_*dag*_(*t*) be the annual ‘excess mortality’ associated with disease *d*, for age *a*, and sex *g*, during calendar year *t*. We use the Institute for Health Metrics and Evaluation (IHME) definition of excess mortality [2], that is disease-specific mortality divided by disease prevalence. Then let 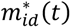 denote

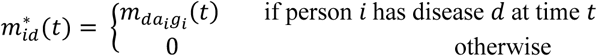

where 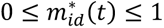. Then the probability *s*_*i*_(*t*) that the *i*^th^ person survives diseases 1, …, *N*_*D*_ in year *t* is given by

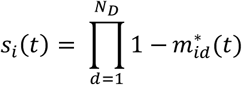

Then the number of people *N*_*sag*_(*t*) of age *a*, and sex *g* who survive diseases 1, …, *N*_*D*_ in year *t* is given by

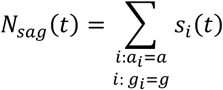

and so the proportion of people *S*_*ag*_(*t*) of age *a*, and sex *g* who survive diseases 1, …, *N*_*D*_ in year *t* is given by

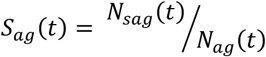

where *N* is the number of people in the population, and *N*_*ag*_(*t*) denotes the number of people of age *a* and sex *g* at time *t*.

If *M*_*ag*_ (*t*) and *RM*_*ag*_(*t*) respectively denote the population-level mortality rate and residual mortality rate (of diseases *not* modelled in Health-GPS), for people of age *a* and sex *g*, then we have

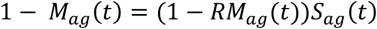

and so rearranging gives

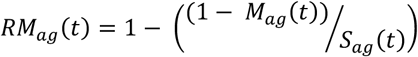

This equation is used throughout the model. It is calculated in the baseline scenario, and the corresponding values are then used in the simulation of intervention scenarios. This assumes that mortality from non-modelled causes of death is constant across scenarios. However, diseases are not always assumed to be independent in Health-GPS, which means that variations in morbidity from modelled diseases that are produced by an intervention may impact non-modelled causes of death, and therefore residual mortality. Therefore, the assumption that residual mortality is constant across scenarios is an approximation and may lead to a (small) underestimation of the effects of an intervention when such an intervention produces a reduction in morbidity from modelled diseases.

At any given time *t*, each individual *i* is exposed to a specific risk of death in connection with the diseases they have, plus a residual risk in connection with causes of death that are not explicitly modelled in the simulation. Individuals who reach age 110 (i.e., the maximum attainable age in Health-GPS) are assumed to die at the next time step.

In the simulation, each year individuals die with probability 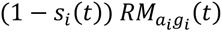.

###### D2.4. Population

The demographic module uses historical and projected population changes by year, sex, and age. UN population projections are available in 5-year age groups, and we linearly interpolate the population for each age group between two years. Similarly to deaths, population numbers are smoothed. This conserves the total annual population by sex.

###### D2.5. Net migration

While data on births and deaths are publicly available in several databases, finding accurate data on migration is more challenging. We model migration using the following formula:

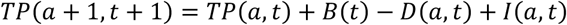

where *TP*(*a*, t), *B*(*t*), *D*(*a*, t) and *I*(*a*, t) respectively denote the total population, number of births, number of deaths, and net migration for age *a* in year *t*.Therefore, migration is simulated indirectly to replicate expected numbers from the UN World Population Prospects database [1]. Consequently, depending on the sign of net migration, individuals will be added or removed from the population. In particular, the attributes of migrants are bootstrapped (i.e., sampled with replacement) from the distributions of the existing individuals of the same sex and age.

##### D3. Risk factor modules

Health-GPS models risk factor exposures accounting for hierarchical dependencies. For instance, characteristics of the living environment (e.g., neighbourhood deprivation) may influence individual behaviours (e.g., dietary intake), and this in turn, may influence the likelihood of conditions (e.g., obesity, hypertension) typically linked with the incidence of NCDs. Diseases themselves can be risk factors for other diseases. Diabetes, for example, is associated with a higher incidence of cardiovascular disease or chronic kidney disease [3,4]. The associations between risk factors are complex and dynamic. They can change due to exogenous shocks such as changes in food prices, or the implementation of new interventions and policies. As individuals age in the simulation, their risk exposures are updated in line with the latest observed age- and sex-specific prevalences (those used to initialise the synthetic population at the start of the simulation). Such age- and sex-specific prevalences are typically held constant throughout the projection period unless specific underlying trends are assumed (in some of the analyses in this project, for instance, a future trend in ultra-processed food consumption has been assumed). Future trends over time in risk factor exposures may apply uniformly for the entire population, or differently for different individuals or population sub-groups.

In the version of the Health-GPS model used for the evaluation of food tax policy scenarios in India, the most upstream risk exposure is food prices, which are assumed to influence food purchases and eventually nutrient intakes. All of the above risk exposures are modelled at the household level outside Health-GPS, using a separate demand system model, described in the paper. Individual nutrient intakes are calculated based on household food purchases and are used as inputs into Health-GPS microsimulations. Nutrient intakes are assumed to generate health impacts through two main pathways: energy and sodium. These two pathways are described in the following sections, 3.1 and 3.2.

**Figure D2.**
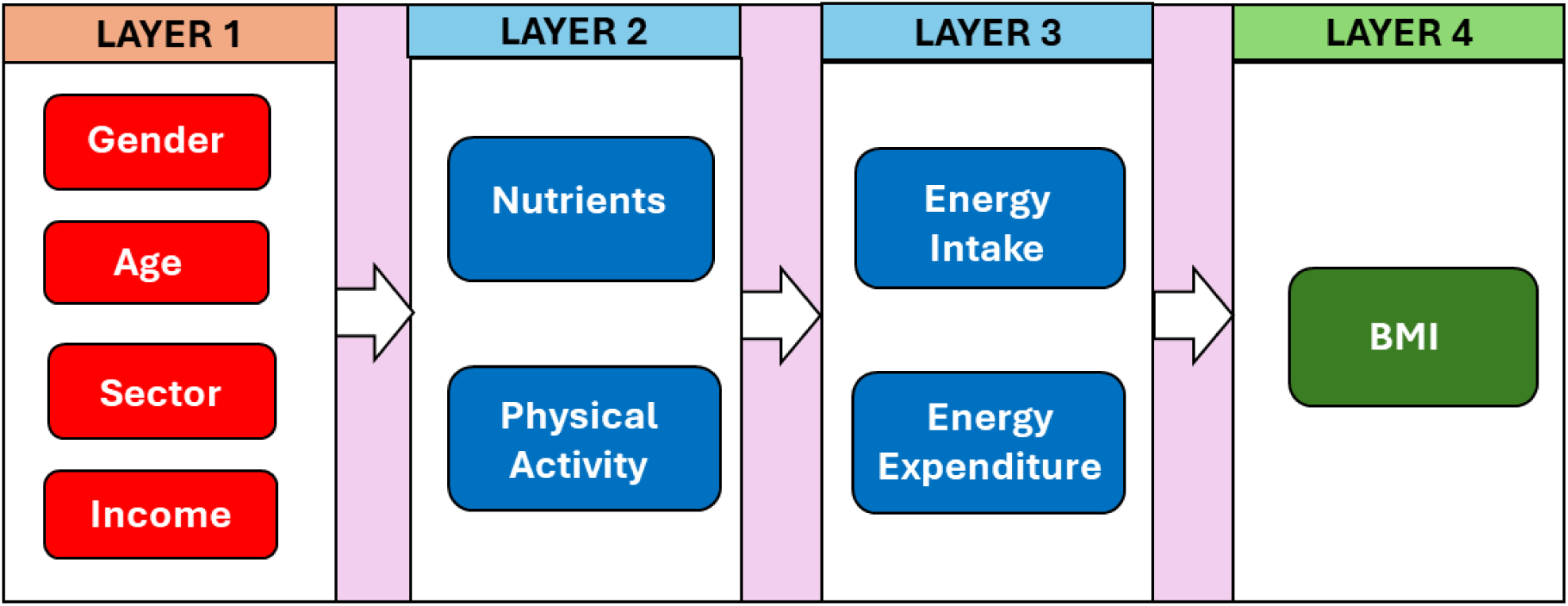
Energy balance model layers and factors, used within Health-GPS. Notes: BMI: body mass index.

###### D3.1. Energy balance model

Health-GPS models body weight dynamics through an energy balance model (EBM) following the mathematical framework developed by Hall et al (2022) [5]. The purpose of this model is to understand and predict how various factors, such as diet and physical activity, influence an individual’s body weight over time. In this section, we provide an overview of how Health-GPS incorporates the Hall et al (2022) model [5]. Full details can be found in Hall et al (2022) [5] and all Health-GPS model codes (including the implementation of the Hall et al (2022) model) can be found at https://github.com/imperialCHEPI/healthgps.

The Hall et al (2022) model takes into account the principles of energy balance, which is the relationship between the calories consumed through food and beverages and the calories expended through metabolism and physical activity [5]. The model incorporates complex physiological processes and feedback mechanisms to provide a more realistic representation of weight dynamics than simpler, linear models.

In this framework, energy intake and expenditure are the result of diet, physical activity and metabolic patterns. Body weight changes in the microsimulation are associated with dynamic imbalances between energy intake (*EI*, the calories consumed from food and beverages), and energy expenditure (*EE*, the energy expended throughout the day). When an individual is in a steady state energy balance (i.e., “energy in” equals “energy out”), body weight is assumed to remain stable. A surplus of energy is stored as fat, leading to weight gain. Conversely, an energy deficit leads to weight loss.

Energy intake and expenditure are determined by nutrient intakes (fat, carbohydrates, protein), physical activity, and prior body mass index (BMI) as a proxy for metabolic energy requirements, which in turn are determined by individual characteristics including age, sex and socioeconomic status (**Figure D2**). The output of the energy balance model is a (time-dependent) change in BMI.

Each layer in **Figure D2** influences subsequent layers. Risk factors are simulated in two steps: 1) initialisation, using static risk factor equations; 2) projection, which updates values for each risk factor at every time cycle during the simulation using dynamic risk factor equations.

###### D3.2. Sodium intake

Sodium intake is modelled alongside nutrient intake that contribute to the EBM. Sodium intake affects disease incidence directly, through relative risks derived from existing studies. Most of the sodium-attributable risk of disease is generated through an increase in blood pressure caused by excessive sodium intakes, however, the individual steps of this causal chain are not modelled in Health-GPS. The relative risks used to model increases in disease risk linked with sodium intake account for the increased blood pressure that mediates the effect of sodium intake on diseases. On the other hand, changes in hypertension prevalence are calculated in parallel as a function of sodium intake.

###### D3.3. Risk factor initialization and propagation

We use a two-stage approach to initialize risk factors within each person in Health-GPS that combines regression modelling, data transformation, and stochastic variation.

###### Nutrient initialisation

First, we use a regression model to determine average values of nutrient intake by age, sex, income (i.e., high, middle and low) and sector (i.e., rural or urban). Second, for each person and each nutrient, we add a stochastic variation term to these average values to generate the required population heterogeneity. This stochastic variation term is sampled from the residuals of the regression performed in the first step.

To ensure normality of the nutrient intake variables, a Box-Cox transformation is applied:

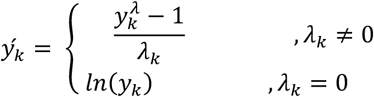

where *ý*_*k*_ is the transformed variable, *y*_*k*_ is the dependent variable (nutrient intake) and *λ*_*k*_ is the transformation variable for nutrient *k* = 1, …, 4.

After transforming the variables, average daily nutrient intake *ý*_k_ is initialized using the equation below for each of the following four nutrients: fat, protein and carbohydrates (measure in grams per day); and sodium (measured in milligrams per day). For nutrient *k*, we have

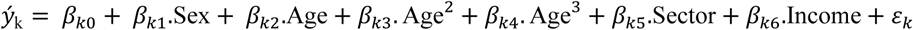

where *β*_*k*0_ is the intercept and *ɛ*_*k*_ is the error term for nutrient *k*. Data input for socio-economic variables and nutrient intake are based on NSS 68^th^ round 2011-2012 and adjusted to December 2022 using the methods detailed in the Methods section of the paper.

To capture realistic variation and maintain correlations between nutrients, stochastic residuals (*ε*) are generated using Cholesky decomposition.

###### Energy intake and energy expenditure initialisation

After daily nutrient intake values have been assigned for each person, they are inputted into the Hall et al (2022) model,^5^ together with each person’s Physical Activity Level (*PAL*), to determine each person’s energy intake (*EI*), and energy expenditure (*EE*). Physical Activity Level (*PAL*) is the ratio between energy expended through physical activity and the person’s baseline metabolic rate. Individual *PAL* values are initialized from a normal distribution

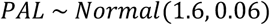

and subsequently adjusted to maintain population-level expected values. Energy expenditure (*EE*) is calculated as a function of body composition and several metabolic parameters, integrating multiple physiological processes, including resting metabolic rate, thermic effect of food, adaptive thermogenesis, and energy partitioning. Further details can be found in Hall et al (2022) [5].

Energy intake (*EI*) quantifies the total caloric content consumed and is a function of the daily nutrient intakes calculated above.

###### BMI calculation

Body Mass Index (BMI) is calculated using the standard definition

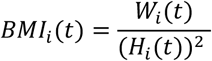

where *W*_*i*_(*t*) and *H*_*i*_(*t*) respectively denote individual *i*’s weight and height at time *t*.

*W*_*i*_(*t*) is initialized as a function of age, sex, energy intake and physical activity. *H*_*i*_ is calculated based on population-level expected values given a person’s age and sex, their weight, as well as randomly sampled individual variation. Height is assumed to remain constant after age 19. Individual BMI values are calibrated to the BMI distribution for India in the NCD-RisC database through an adjustment of weight and nutrient intake values [6].

Using the initialized values for weight, height, energy intake and physical activity, the Hall et al (2022) model is used to initialize values of body fat, lean tissue, glycogen, extracellular fluid and energy expenditure for each person [5]. The changes in these values (governed by changes in nutrient intake and energy intake) are then used to update each individual’s weight, and thus their BMI.

##### D4. Disease modules

Health-GPS is calibrated to match empirical disease distributions. Changes in the distributions of risk factors and diseases are then transferred to other diseases. Although we assume the associations of risk factors to diseases to be constant throughout the simulation, any deviation of a risk factor from its initial distribution will translate into increased or decreased disease incidence through relative risk equations (the epidemiological literature does not always distinguish clearly between relative risks and hazard ratios; when it is not possible to identify values for both, available values are used interchangeably in Health-GPS). Therefore, any change in the prevalence of a disease is solely caused by changes in risk factor distributions, alongside the ageing of the population.

The disease module uses multiple data sources, including IHME for prevalence, incidence, mortality and remission rates for non-cancerous diseases [2]. When cancers are modelled (which is not the case in this study), Health-GPS uses data from the International Agency for Research on Cancer (IARC) for prevalence at 1, 3 and 5 years (from diagnosis), incidence, and mortality rates [7]. Relative risks from BMI to diseases are derived from IHME attributable risk data, while those for sodium to ischaemic heart disease, stroke, and chronic kidney disease are derived from specific studies [8,9].

Probabilities of disease for each individual are updated throughout the simulation to reflect changes in risk exposures and disease profiles. Prevalence in the initial population is used to calibrate Health-GPS parameters. However, as time progresses, risk factor distributions and demographics are the sole drivers of prevalence.

The excess mortality rate is the proportion of excess deaths within a designated population of “cases” (people with a medical condition) over the course of the disease. These rates are kept constant over time in the simulation. Remission is the rate at which people with a disease revert to a state with the same survival prospect and disability level as those without the disease. Remission rates are kept constant over time in Health-GPS. In this study, remission rates are taken from IHME [2]. When not available, they are assumed zero.

Disease is modelled through 3 types of events: onset, remission and fatality. An individual’s probability of acquiring a disease is calculated based on their risk exposures, existing conditions, and the population-level incidence of each disease. Therefore, this incidence is adjusted for each individual to account for their sex, age, risk and disease profiles. For each individual in the population and for each disease, we compute an Individual Relative Risk (IRR), which reflects the aggregate change in disease incidence associated with the individual’s risk factors and diseases. After disease onset, the individual may either go into remission or die, over time.

Let *RR*_*dfag*_ be the relative risk of disease *d* associated with risk factor *f* = 1, …, *N*_*f*_ for age *a* and sex *g*. Let *DR*_*dagj*_ be the relative risk of disease *d* associated with the presence of another disease *j* (where *j* = 1, …, *N*_*D*_). for age *a* and sex *g*. If

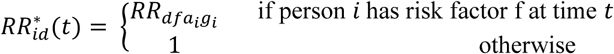

and

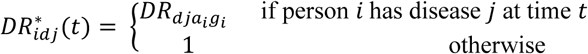

then individual *i*’s overall relative risk *IRR*_*id*_ (t) of disease *d* is given by

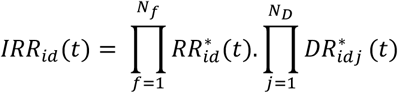

At the beginning of the simulation, each individual’s disease status is assigned based on: (a) their *IRR*_*id*_ (*t*_0_); (b) the mean value of this quantity among people of the same age and sex; and (c) the prevalence of disease *d* in the same age and sex group. Therefore, individual *i* is assigned disease *d* with probability equal to

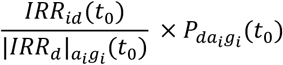

where *P*_*dag*_ (*t*_0_) and |*IRR*_*d*_ |_*ag*_ respectively denote the population level prevalence, and the average relative risk of disease *d* for age *a* and sex *g*, at the beginning of the simulation. Note that this formulation assumes relative risks combine multiplicatively and are independent.

Similarly, if *I*_*dag*_(*t*_0_) denotes the population level incidence of disease *d* for age *a* and sex *g*, as currently observed empirically (at *t0*), then individual *i* develops disease *d* at time *t* with probability equal to

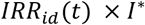

and

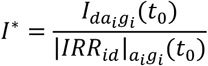

*I*^∗^is calculated in the baseline scenario and is held constant throughout the simulation. It is also used throughout the simulation of intervention scenarios.

Finally, each person of age *a* and sex *g* with disease *d* has probability *ρ*_*dag*_ of going into remission each year, and dies with probability 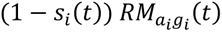 each year (see above section on Residual Mortality).

##### D5. Burden of disease module

If *DW*_*d*_ represents the disability weight associated with disease *d*, let

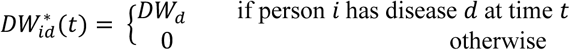

Now define the product *X*_*i*_(*t*) of the compliment of disability weights *X*_*i*_(*t*) by

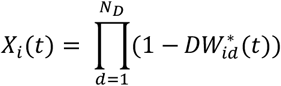

and the average of this product for age *a* and sex *g* is

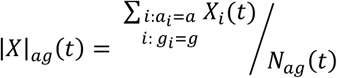

where *N*_*ag*_ (*t*) denotes the number of people of age *a* and sex *g* at time *t*. If *YLD*_*ag*_ (0) is the empirically observed years lived with disability for age *a* and sex *g* over all diseases at the time of initialization, and if *RDW*_*ag*_ is the residual disability weight for diseases *not* included in Health-GPS for age *a* and sex *g*, then at time *t* = 0, we have

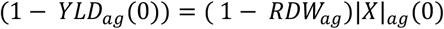

Rearranging gives

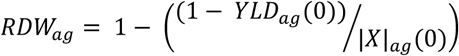

*RDW*_*ag*_ is initialized as above at the beginning of the simulation and unchanged after. More generally, across diseases (modelled in Health-GPS or otherwise) at time *t*, each person *i* has disability weight *DW*_*i*_ (*t*) given by

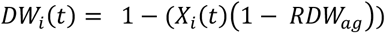

Disability-adjusted life years (DALYs_*ag*_(*t*)) are calculated as the sum of the years lost due to premature mortality (*YLL*_*ag*_ (*t*)) and the years lived with a disability *YLD*_*ag*_ (*t*) over the time horizon of the simulation, for age *a* and sex *g*. *YLD*(*t*) at time *t* is given by

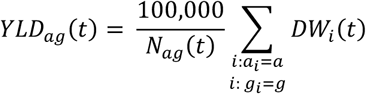

The contribution of person *i* to years of life lost at time *t* is given by

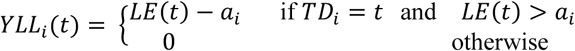

where *a*_*i*_ is their age, *TD*_*i*_ gives their time of death, and *LE*(*t*) = max {*LE*_Male_(*t*), *LE*_Fenale_ (*t*)} is the life expectancy at time *t*. Then

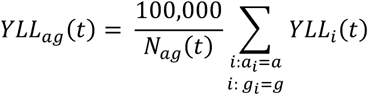

The DALYs at time *t* are then given by

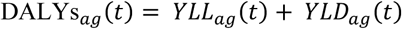

##### D6. Health expenditure

Total health expenditure (THE) consists of current health expenditure (CHE) and capital expenditure. According to National Health Accounts Estimates for India 2021-22, CHE is 87.32% of THE, and of the CHE, governments’ share is 38.79% and the households’ share is 50.55% (including out-of-pocket expenditure and insurance contributions). Out-of-pocket expenditure (OOPE) specifically account for 45.11% of THE [10]. Therefore, households’ expenditure and households’ OOPE on health accounts for 44.14% and 39.39% of THE, respectively.

For each disease in asthma, chronic kidney disease, diabetes, ischemic heart disease, and stroke (including ischemic stroke, intracerebral haemorrhage, and subarachnoid haemorrhage), either households’ expenditure, or OOPE on health, per person per year in India is derived from previous studies [11–13], depending on availability, and is then converted into THE per case following the method adopted by WHO in NCD investment cases [14]. Finally, the averted annual THE of each disease under intervention is calculated as the product of reduced cases under intervention and THE per case.

##### D7. Hypertension

Changes in hypertension prevalence are estimated independently with data on hypertension prevalence in India from the National Family Health Survey (NFHS-5), 2019-21 [15], estimates on changes in sodium and BMI under intervention from Health-GPS, and the association between sodium, BMI and the risk of hypertension from literature [16,17].

A linear association is assumed between BMI and the risk of hypertension, and between sodium and the risk of hypertension. One unit change in sodium (mg) and BMI (kg/m^2^) is associated with a change of 0.00003 and 0.186 in the risk of hypertension, respectively [16,17]. Reduction in prevalence number of hypertension by age and sex is calculated as follows.

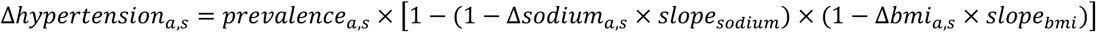

Where *a* denotes age and *s* denotes sex. *prevalence*_*a*,*s*_ is the baseline prevalence number of hypertension from NFHS-5. Δ*sodium*_*a*,*s*_ and Δ*bmi*_*a*,*s*_ are policy effect estimates from Health-GPS. *slope*_*sodium*_ = 0.00003. *slope*_*bmi*_ = 0.186.

**Table D1.** Model parameters and variables.

| Symbol | Definition | Source/Assumption |
| --- | --- | --- |
| Indices |  |  |
| $i$ | Index of person | |
| $a$ | age | 0-100 |
| $g$ | Index of sex | Male/Female |
| $d$ | Index of disease | |
| $k$ | Index of nutrient | |
| $j$ | Index of disease other than disease $d$ | |
| $l$ | Index of covariate | |
| Demographic module |  |  |
| $t$ | Calendar time (year) | 2022-2053 (policy introduced in 2024) |
| $m_{dag}(t)$ | Annual ‘excess mortality’ associated with disease $d$, for age $a$, and sex $g$, during calendar year $t$ | IHME [2] |
| $N_D$ | Number of diseases simulated | 7 in this study including asthma, chronic kidney disease, diabetes, ischemic heart disease, and stroke (including ischemic stroke, intracerebral haemorrhage, and subarachnoid haemorrhage) |
| $s_i(t)$ | Probability that the $i^{\text{th}}$ person survives all simulated diseases in year $t$ | Calculated in Health-GPS |
| $N_{sag}(t)$ | Number of people of age $a$, and sex $g$ who survive all simulated diseases in year $t$ | Calculated in Health-GPS |
| $S_{ag}(t)$ | Proportion of people of age $a$, and sex $g$ who survive all simulated diseases in year $t$ | Calculated in Health-GPS |
| $N_{ag}(t)$ | Number of people of age $a$ and sex $g$ at time $t$. | UN World Population Prospects database [1] |
| $M_{ag}(t)$ | Age- and sex-specific population-level mortality rate in year $t$ | UN World Population Prospects database [1] |
| $RM_{ag}(t)$ | Age- and sex-specific population-level residual mortality rate (of diseases not modelled in Health-GPS) in year $t$ | Calculated in Health-GPS |
| $TP(a, t)$ | Total population of age $a$ in year $t$ | UN World Population Prospects database [1] |
| $B(t)$ | Births in year $t$ | UN World Population Prospects database [1] |
| $D(a, t)$ | Deaths of age $a$ in year $t$ | UN World Population Prospects database [1] |
| $I(a, t)$ | Net migration for age $a$ in year $t$ | UN World Population Prospects database [1] |
| $income_i(t)$ | Income tertile of individual $i$ in year $t$ | NSS Household Consumption Expenditure survey 2022-23 |
| $sector_i(t)$ | Sector of residence (urban or rural) of individual $i$ in year $t$ | NSS Household Consumption Expenditure survey 2022-23 |
| Risk factor module |  |  |
| $y_1$ | Value of nutrient 1: Carbohydrate (untransformed) | NSS Household Consumption Expenditure survey 2022-23 |
| $y_2$ | Value of nutrient 2: Protein (untransformed) | NSS Household Consumption Expenditure survey 2022-23 |
| $y_3$ | Value of nutrient 3: Fat (untransformed) | NSS Household Consumption Expenditure survey 2022-23 |
| $y_4$ | Value of nutrient 4: Sodium (untransformed) | NSS Household Consumption Expenditure survey 2022-23 |
| $y'_k$ | Box-Cox transformed value of nutrient $k$ | Calculated in Health-GPS |
| $\lambda_k$ | Box-Cox transformation parameter for nutrient $k$ | Calculated with $y_k$ |
| $\beta_{kl}$ | Linear regression coefficient for nutrient $k$ and covariate $l$ | Calculated with $y'_k$, $a$, $g$, $income_i(t)$ and $sector_i(t)$ |
| $\varepsilon_k$ | Error term for nutrient $k$ | Calculated with $y'_k$, $a$, $g$, $income_i(t)$, $sector_i(t)$ and $\beta_{kl}$ |
| $\sigma_k$ | Standard deviation of $\varepsilon_k$ | Calculated with $\varepsilon_k$ |
| $PAL$ | Physical Activity Level | A normal distribution with mean as 1.6 (i.e., 1.6 times basic metabolic requirement) and standard deviation as 0.06 |
| $W_i(t)$ | Weight of individual $i$ at time $t$ | Distribution from NCD-RisC [6] |
| $H_i(t)$ | Height of individual $i$ at time $t$ | Distribution by age and sex from NCD-RisC [6] |
| $slope_g$ | Slope of height over weight for sex $g$ | Female: 0.3564; Male: 0.3718; estimated based on NCD-RisC data [6] |
| $std_g$ | Standard deviation of height for sex $g$ | Female: 0.096; Male: 0.085; estimated based on NCD-RisC data [6] |
| $BMI_i(t)$ | Body Mass Index of individual $i$ at time $t$ | Calculated with $W_i(t)$ and $H_i(t)$ |
| Disease module |  |  |
| $RR_{dfag}$ | Age- and sex-specific relative risk of disease $d$ associated with risk factor $f$ | IHME [2] and literature [8-9] |
| $N_f$ | Number of dietary risk factors simulated | Given by authors, 4 in this study (carbohydrates, protein, fat, and sodium) |
| $DR_{dagj}$ | Age- and sex-specific relative risk of disease $d$ associated with presence of another disease $j$ | IHME [2] and literature [18-19] |
| $IRR_{id}(t)$ | Individual $i$'s overall relative risk $IRR_{id}(t)$ of disease $d$ at time $t$ | Calculated in Health-GPS |
| $P_{dag}(t_0)$ | Age- and sex-specific population level prevalence of disease $d$ at the beginning of the simulation | IHME [2] |
| $\rho_{dag}$ | Age- and sex-specific probability of going into remission from disease $d$ each year | IHME [2] |
| Burden of disease module |  |  |
| $DW_d$ | Disability weight associated with disease $d$ | IHME [2] |
| $RDW_{ag}$ | Age- and sex-specific residual disability weight for diseases *not* included in Health-GPS | Calculated in Health-GPS |
| $YLL_{ag}(t)$ | Age- and sex-specific years lost due to premature mortality over the time horizon of the simulation | Calculated in Health-GPS |
| $YLD_{ag}(t)$ | Age- and sex-specific years lived with a disability over the time horizon of the simulation | Calculated in Health-GPS |
| $YLL_i(t)$ | Contribution of person $i$ to years of life lost at time $t$ | Calculated in Health-GPS |
| $TD_i$ | Time of death of person $i$ | Calculated in Health-GPS |
| $LE(t)$ | Life expectancy at time $t$ | Calculated in Health-GPS |
| $DALY_{s_{ag}}(t)$ | Age- and sex-specific disability-adjusted life years at time $t$ | Calculated in Health-GPS |

### Supporting information file 5

#### Appendix E. Income trend assumption

Appendix E forms part of the revised submission.

Supplement to: Roche M, Zhu J, Olney J, Laydon DJ, Joe W, Sharma M, Steele L, Sassi F. Taxation of foods high in saturated fat, sugar, and sodium in India: A modelling study of health and economic impacts. Submitted after revision on 19 September 2025.

##### Description

The income trend factor *f*_*i*,*j*,0_ for income group *i* and food group *j* for period *t* = 0 is estimated as the product of the income elasticity estimate (*η*_*i*,*j*_) (**Table C1**) and an income growth assumption. We assume a constant 6% income growth for India based on IMF projections [2].

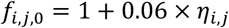

Food consumption for period *t* = 1 is then estimated at the food item level by multiplying *f*_*i*,*j*,0_ with the household level total daily consumption at *t* = 0. We derive household level daily intake of energy and nutrient by multiplying the estimated daily consumption for each item at *t* = 1 with each item’s energy and nutrient content derived from food composition tables. We sum this up at household-level to obtain the estimated household daily total intake of energy and nutrient. We then individualise daily intake based on household members’ age and sex using the daily NIN Dietary Guidelines for Indians 2024 to obtain the estimated daily individual nutrient intake (*q̂*_*i*,*n*,1_) for individual *i*, nutrient *n*, and period *t* = 1. The baseline individual-level nutrient-specific income trend factor *f*_*i*,*n*,0_ for individual *i* and nutrient *n* is recovered as below:

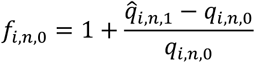

We then run the below cross-sectional OLS regression:

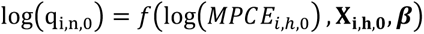

Where *MPCE*_*i*,*h*_ represents the per capita monthly total expenditure for household *h* and **X**_**i,h**_ a vector of household and individual characteristics including the household sector, size, religion, and the individual sex, age, and education. We obtain the vector of coefficients ***β̂***.

We recover the predicted individual level nutrient intake for future periods as:

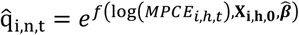

Where *MPCE*_*i*,*h*,*t*_ = *MPCE*_*i*,*h*,0_ × 1.06^*t*^ based on the constant IMF projection assumption for real GDP per capita.

The nutrient-specific decay rate *b̂*_*n*_ is finally recovered as the coefficient from the below OLS regression of the logarithm of the predicted growth rate in nutrient intake on a time trend:

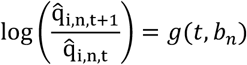

As expected from Engel’s Law, which posits that, as household income increases, the proportion spent on food decreases, *b*_*n*_ < 0. To align with Engel’s Law [2], the individual-level nutrient-specific income trend factor for period *t* is scaled by an exponential decay over time and is recovered using the below formulae:

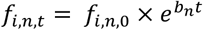

**Figure C1** shows that our income elasticities are well-behaved, i.e., higher for low-income households across food groups, in line with Engel’s Law [2]. **Figure E1** cross-validates this approach by comparing the estimated yearly growth rate in the consumption of packaged processed foods and sweets - two food groups with a high proportion of HFSS foods (**Table A6**) - with historical trends based on Euromonitor International Passport data and Tak et al (2022) for selected highly processed food items [3,4].

**Figure E1.**
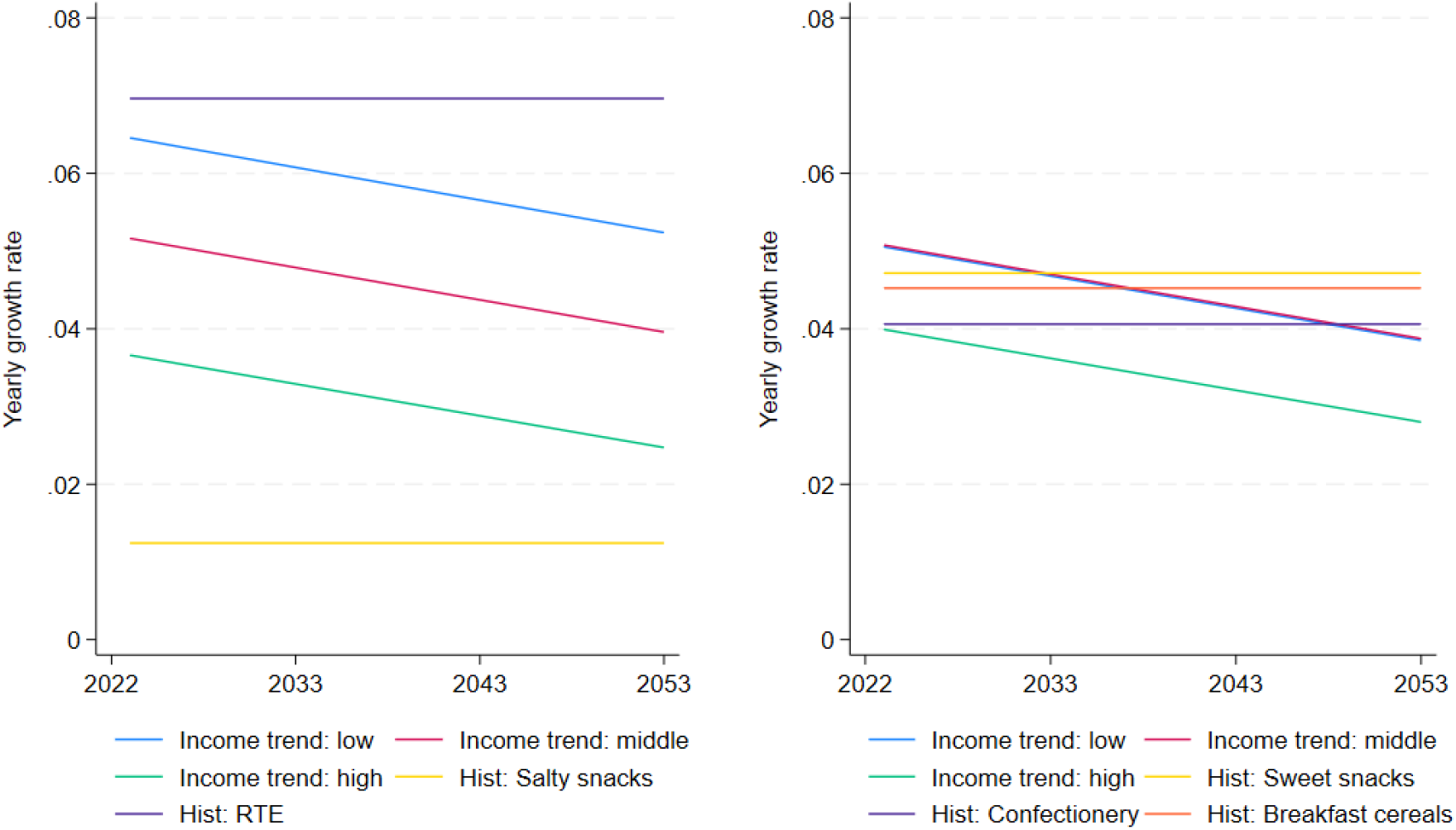
Comparing projection assumptions for highly processed food items. Notes: Historical trends based on Euromonitor International Passport data and Tak et al (2022) for selected highly processed food items [3,4]. Income group categorized using NSS Household Consumption Expenditure Survey 2022-23 monthly total household expenditure per capita. Hist: historial, RTE: ready-to-eat.

